# Spatial Point Pattern Analysis of Environmental Effects on Valley Fever Intensity in Phoenix, Arizona

**DOI:** 10.1101/2025.07.14.25331482

**Authors:** John Ginos, Beckett Sterner, Petar Jevtić

**Affiliations:** School of Mathematical and Statistical Sciences, Arizona State University, 901 Palm Walk, Tempe, 85281, Arizona, USA; School of Life Sciences, Arizona State University, 427 East Tyler Mall, Tempe, 85281, Arizona, USA

**Keywords:** Valley fever, Spatial point pattern, *Coccidioides*, Coccidioidomycosis, Phoenix, Medicaid, NDVI, Soil

## Abstract

Using a novel Arizona Medicaid data set, we model Valley fever (VF) cases in the Phoenix Metropolitan Area as a spatial point process during six month intervals between 2013-2023. We estimate the intensity function of VF cases, observed at residential locations of patients, as a function of environmental covariates available at high spatial resolutions, including land cover type, change in land cover type, Normalized Difference Vegetation Index (NDVI), change in NDVI, and a Habitat Suitability Index (HSI) for the *Coccidioides* fungus. Models with all covariates included and quadratic terms for NDVI, NDVI difference, and HSI tend to perform the best. We show that land cover change and HSI measured at the residential (point) level are useful for understanding VF risk. Furthermore, we find that NDVI and its semesterly changes appear to have a nonlinear relationship with the intensity of VF cases. Despite more pronounced peaks in predicted intensities for our most complex model compared to our simplest model, we show that even our best, most complex model underestimates the intensity in portions of the study region. Thus, our results provide initial evidence that spatially localized environmental information is useful for examining VF incidence among Medicaid patients in the study area.

## 1. Introduction

Coccidioidomycosis, commonly known as Valley fever (VF), is a disease caused by the fungal pathogens *Coccidioides posadasii* and *C. immitis*, which are endemic to North America. The geographic range of Valley fever and number of human cases are growing, especially in the western United States [20, 18]. Identifying environmental factors that influence VF incidence is important to public health authorities and communities. Among the environmental factors identified in recent work on VF incidence and seasonal variability are temperature, precipitation, air quality, and soil properties (see [22], [15], and [36]; see also [27] for a succinct summary of variables and methods used in the analysis of VF data). Often, these studies aggregate VF cases to administrative geographical units like census tracts, counties, or cities (e.g. see [19], [22], and [27], respectively). Aggregation of spatially referenced location data to larger geographical units may present problems for data analysis including the modifiable areal unit problem [12], in which results from spatial data analysis are greatly impacted by the scale of aggregation. A related example of this when analyzing VF data specifically is discussed in [11], where the authors report that percentages of land cover type in geographical units showed drastic variation depending on the spatial scale of units used (e.g. census tract, block group, and zip code). We seek to avoid this problem to the greatest extent possible by considering the VF case locations (by residence) at the point level and using higher resolution covariate data. However, since exposure to *Coccidioides* arthroconidia is not limited to one’s residential location, we simultaneously explore the utility of spatially local covariate information (i.e. information around one’s residence) in understanding VF risk.

To these ends, we employ the first spatial point process model^1^ of residential addresses of human VF cases based on dataset of Arizona Medicaid patients^2^ . The dataset incldues VF cases of Medicaid patients in and around the Phoenix Metropolitan Area during 2013 to 2023 (hereafter referred to as the study period). In 2023, Maricopa County, Arizona, which includes the greater part of the Phoenix Metropolitan Area accounted for roughly 38% of VF cases according to provisional data from the Centers for Disease Control and Prevention (CDC) as well as figures reported by Maricopa county; see [13] and [24].

While one-dimensional temporal point process models have been employed to understand VF incidence over time [34], we are not aware of any formal treatment of VF cases as a spatial point pattern. The most granular analysis of VF data in the literature is the logistic regression approach in [11], where the authors used presence/absence data derived from residential addresses of those with VF and pseudo-absence data randomly generated based on the population of census block groups in Arizona. A comparison of spatial point pattern analytic methods with the logistic regression approach (using pseudo-absences) similar to the one used in [11] and the limitations of this latter approach are discussed in [35] in the case of ecological data analysis. We consider environmental factors at all locations where individuals could live in the study window at the finer spatial scale. To accomplish this, we treat the residential addresses of VF cases as a spatial point pattern and employ well-established techniques^3^ to analyze factors that influence the spatially inhomogeneous intensity of VF cases. This approach also allows us to obtain fitted intensities for the whole study region (and at the locations of VF cases themselves), which can be used to determine where the model fit breaks down.

We find evidence supporting spatially localized effects of environmental covariates, including land cover category (including annual changes in land cover category), vegetation level, change in vegetation, and soil habitat suitability for *Coccidioides* fungal pathogens. The support we find for biotic spatial variables such as vegetation level and change is consistent with the enzoonotic hypothesis that rodents play an important role in spreading the disease, although we are not able to directly investigate the importance of wind versus animal-borne dissemination mechanisms as discussed in [27].

While the Medicaid dataset we analyze has several limitations, our results suggest that VF exposure is associated with environmental covariates at a fine spatial scale and that additional environmental variation and/or nonlinear model structure may explain residual spatial clustering we observe in the data based on our best model.

## 2. Data and Methods

### 2.1. Valley fever Data

Locations of residence for individuals diagnosed, treated, and/or tested for VF were obtained from the Arizona Health Care Containment System (AHCCCS; the Arizona state Medicaid agency) during the study period 2013 through 2023. The Arizona Medicaid (AHCCCS)^4^ data used in this study were provided by the Arizona State University, Center for Health Information Research (CHiR). We received approval from the Arizona State University Internal Review Board (Study 20039) prior to the beginning of research and followed all HIPAA protocols to protect human subjects data. The data were aggregated by semester^5^ and year with the first semester of each year defined as January 1st through June 30th; the remaining days of the year constitute the second semester. All individuals represented in the data set (hereafter simply patients) were identified with a member identification code associated with unique patients who were continuously enrolled during a given semester.

Demographic information for each patient including age and sex were included along with whether the individual was tested, treated, and/or diagnosed with VF in a given semester. Only the earliest diagnosis of VF was considered for patients with multiple (non-consecutive) diagnoses to avoid double counting unique diagnoses for individuals who may have unenrolled and re-enrolled in Medicaid during the study period. The demographic information was used to categorize the VF cases into four groups based on age and sex (see 2.3 for more details). Splitting the locations of VF cases into sub-patterns of spatial points allows us to consider how each pattern’s spatial distribution is influenced by the same set of covariates and account for known heterogeneity in VF prevalence among such groups.

Due to the extreme contrast in the spatial density of VF cases between major Arizona metropolitan areas (like Phoenix and Tucson) and rural regions of the state and due to covariate data availability, we decided to limit our analysis to the Central Arizona-Phoenix Long-Term Ecological Research study area within metropolitan Phoenix^6^ (hereafter study window; see Figure 1).

**Figure 1:**
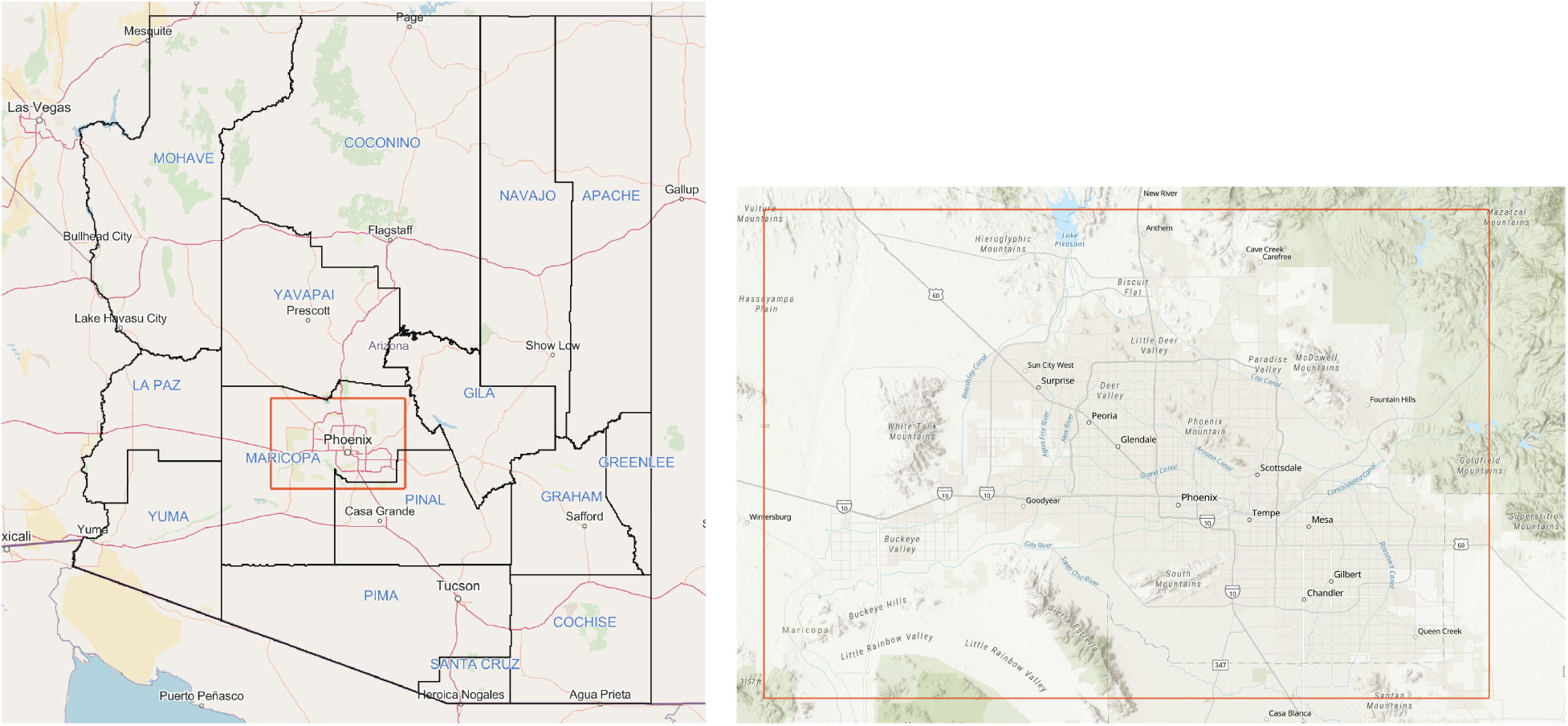
Left: CAP LTER study window (red box) as situated within Arizona. Arizona county borders (black) are shown along with county names in blue. This image was taken as a screenshot of the openstreetmap basemap available in ArcGIS Online [26]. County boundaries obtained from [5] and [2]. Right: CAP LTER study window (taken as a screenshot of the topographic basemap available in ArcGIS Online [17])

### 2.2. Spatial Covariate Data

As mentioned above, we are interested in how the intensity of VF cases is influenced by environmental covariates including the soil Habitat Suitability Index (HSI) of the *Coccidioides* fungus (see [15]), NDVI, land cover category, and changes in land cover category that occur from one year to the next (we call this land cover change). Certain land cover changes are considered relevant for our purposes (see 2.2.5). Aside from environmental covariates, we also use the estimated Medicaid population density to account for the spatially varying population at risk within the study window.

All covariate data were rasterized^7^ (if not already in raster format) and converted to a pixel image object for compatibility with the R package spatstat [8].

#### 2.2.1. Medicaid Population Density (MPD) Estimates

We expect that the spatial distribution of VF cases varies with the Medicaid enrollment population density (MPD) which varies differently than the overall population density due, in part, to socioeconomic factors. Since we did not have access to the true MPD, we used the U.S. Census Bureau’s American Community Survey (ACS) 5-year estimates for Medicaid enrollment for the study period^8^, respectively. Estimates are available at the census block group level; however, when aggregated to the county level, they underestimate the true Medicaid enrollment figures available from AHCCCS during the study period. To address this, we normalized all block group estimates (for all of Arizona) to sum to their respective county-level estimates^9^ with data obtained for AHCCCS (for monthly AHCCCS enrollment data see [6]).

Areas where the Medicaid enrollment population is estimated to be zero present a numerical problem when modeling spatial point data using an offset to account for spatially varying exposure. To avoid zero-MPD areas, we set these block group estimates to one-fourth of the margin of error pertaining to the estimate^10^. The resulting Medicaid enrollment block group estimates are divided by block group area (in square miles) to obtain MPD.

To further reduce potential numerical issues related to outliers and to deal with the uncertainty in the estimates themselves, we rasterize (to 1,000-meter-resolution) and then smooth the MPD block group estimates using a Gaussian kernel. The standard deviation of the Gaussian kernel, often called the bandwidth for data smoothing purposes, is chosen to be 2-kilometers to preserve local features in the MPD estimates but avoid large outliers in the population estimates. Finally, we normalize the smoothed MPD estimates (SMPDE) for use as an offset term in modeling VF cases. This is done by dividing all resulting pixel values by the sum of all pixel values in the study window. Such normalization reduces the remaining large-scale spatial variation in the offset term to avoid numerical problems when model fitting while maintaining positive values at each pixel location.

#### 2.2.2. Normalized Difference Vegetation Index (NDVI)

Raster data for the study region were obtained at the 30-meter resolution from the Environmental Data Initiative [29]. Seasonal NDVI rasters for the study period and the year 2012 were used for the analysis. The average NDVI (by raster cell/pixel value) for winter and spring^11^ of a given year was used for semester 1 covariates while the average NDVI for the summer and fall were used as semester 2 covariates during the study period.

NDVI is known have a lagged positive effect on VF cases for up to and slightly beyond a year (see [28]), though this lagged relationship has not been explored at the finer spatial scale of interest here. Unfortunately, our VF incidence data does not align with the seasonal NDVI data; it is hard to link our case data with seasonal variations in NDVI since individuals in the data set could have been diagnosed or treated at any time in a six month period. For simplicity, we explore the NDVI and lagged NDVI effects up to one semester prior. Once the NDVI averages were obtained for each semester, a *differenced* NDVI covariate was obtained by differencing the NDVI for each semester with the NDVI from the prior semester. For example, for 2013 semester 1, the differenced NDVI covariate is calculated by computing the pixel-wise difference between the winter-spring average NDVI for 2013 and the fall-summer average NDVI for 2012^12^ .Figure 2 shows the normalized difference of fall NDVI rasters from 2012 and 2023. The normalized difference, given by 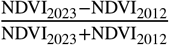, provides a usful pixel-wise calculation for inspecting changes in from the semester prior to and near the end of the study period. Note the significant areas of change in the west and southeastern portions of the study window. The increase in population and urban development in these areas may explain the apparent decreases in NDVI.

**Figure 2:**
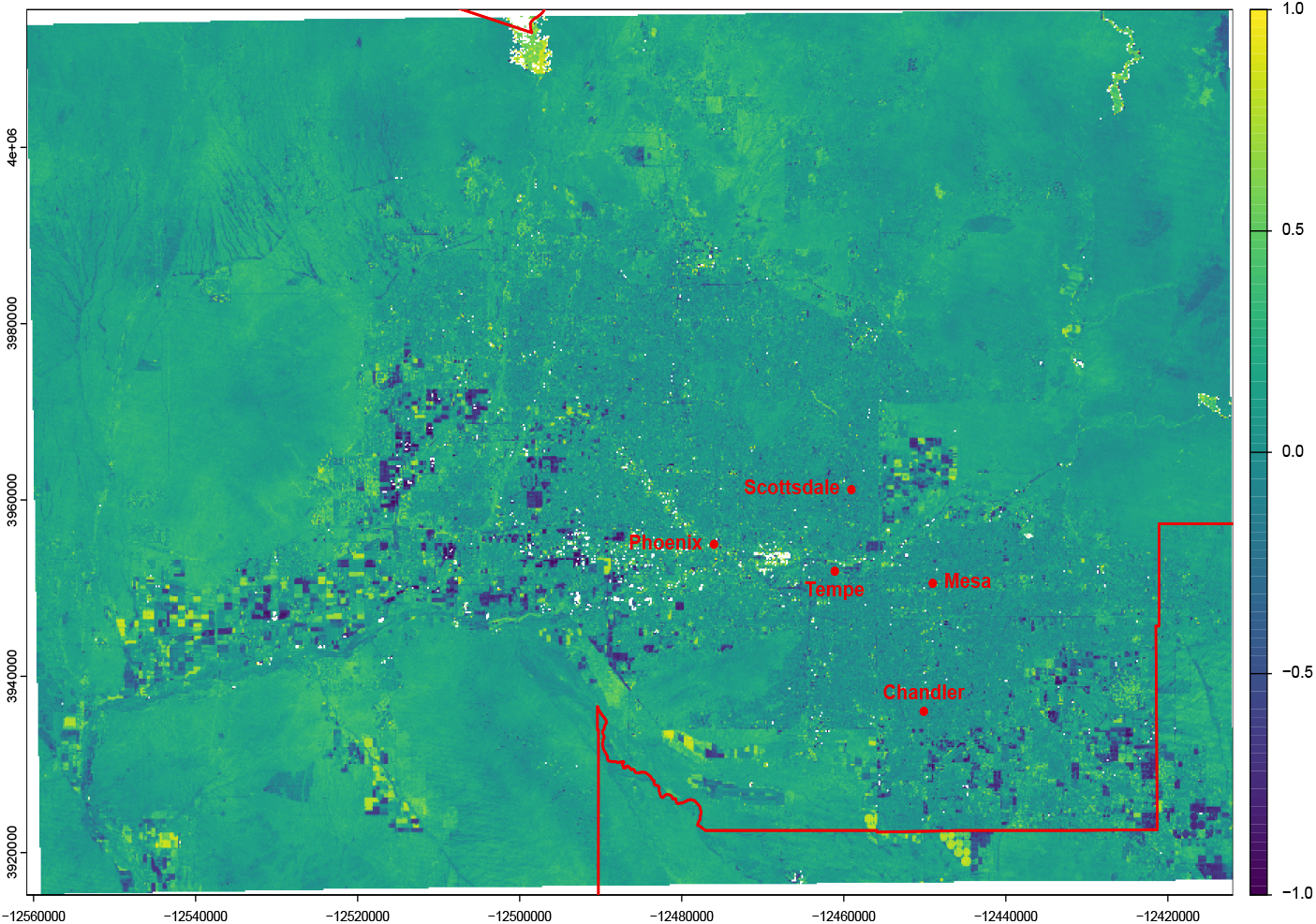
Normalized difference (by pixel) between fall NDVI of 2023 and the fall NDVI of 2012 (the year prior to the start of the study period; 2012 NDVI data were used for creating a differenced NDVI covariate). Bright yellow areas denote an increase in NDVI over this period while dark purple areas represent a decrease in NDVI. Blank spaces are NA. Phoenix and a few nearby city centroids labeled in red along with county borders. **Source: The NDVI data used to create this figure was obtained from [29]**

#### 2.2.3. Coccidioides Habitat Suitability Index (HSI)

Recent work in [15] showed that certain soil characteristics provide suitable conditions for the *Coccidioides* fungus. The authors^13^ in [15] created the Habitat Suitability Index to provide a spatially granular method of measuring where exposure to VF is a greater risk. The index takes values between 0 and 1 with values closer to 1 indicating increased suitability. The index is, in part, derived from soil survey data available and climatic data. A resulting10-meter resolution raster is obtained from the authors’ predicted habitat suitability index within the study region^14^ . Figure 3 shows the HSI raster for the study window; the large block of NA pixels in the northeast corner is due to unavailable soil survey data for the Tonto National Forest. We note that we use a single HSI covariate raster for the entire study period (the raster does not vary by year and semester).

**Figure 3:**
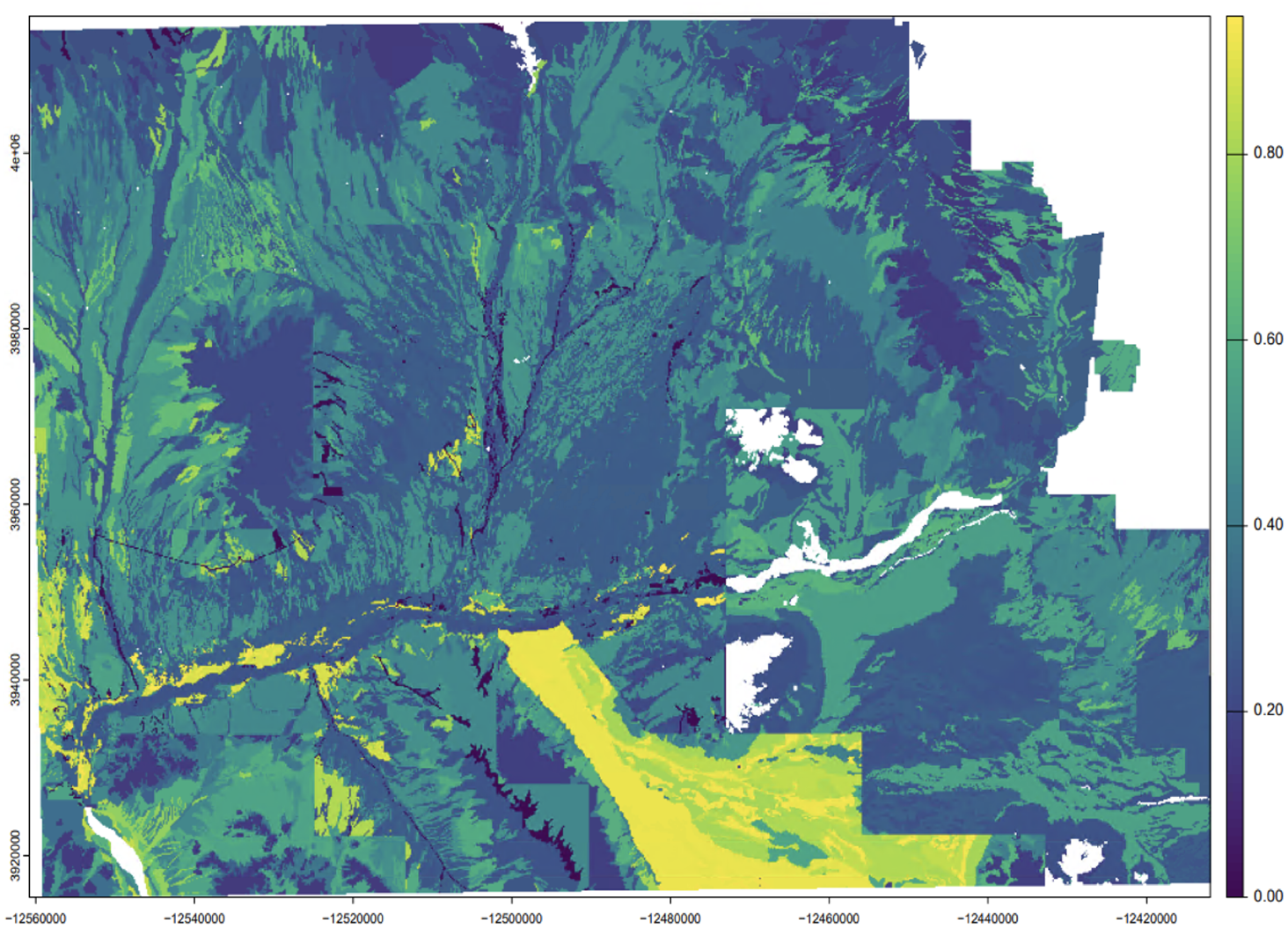
Coccidioides habitat suitability (HSI) index shown at the 10-meter resolution within the Phoenix metropolitan area. **Source:** Data obtained with the help of Robert R. Dobos and links to the original data sources provided in [15]. Blank spaces are NA^16^.

#### 2.2.4. Land Cover Category

The Multi-Resolution Land Characteristics (MRLC) Consortium provides data from the National Land Cover Database (NLCD) for use by the public (see [33]). From the available data, we have obtained annual land cover characteristics at the 30-meter resolution for the study window for 2013 through 2023. We are interested in whether four land cover categories (LCCs), namely open-area development^17^ (LCC1), low intensity development areas (LCC2), medium intensity development (LCC3) areas, and high intensity development (LCC4) areas are associated with an increase in the spatial intensity of VF cases compared with the base category “Other” which includes undeveloped areas of all remaining classifications found in the study window.

As we mentioned above, the Medicaid population density data contains inherent uncertainty. Thus, using land cover categories as a covariate is helpful as another measure to account for the spatially varying population density of the study window. While it is hypothesized that animal hosts of *Coccidioides* are more likely to live in areas with less development, we recall that our data consists of the location of residence for associated VF infections. Thus, we hypothesize a positive association between the spatial intensity of VF cases and areas of higher intensity development due to population concentration in these areas. It is also possible that those who live on less developed land (low-intensity development; LCC1) may be at a greater risk of VF exposure (especially on land with a high HSI) where they live.

#### 2.2.5. Land Cover Change

Construction, land development and renovation of existing structures often involve soil-disturbing activities like excavation. Displacing soil may cause increased risk for VF among both construction workers and those who live near the development site [23] [37]. To consider the effect that such development sites may have on the intensity of VF cases within the target population, we use land cover change data from the NLCD made available by the MRLC. The raster data classify 30-meter pixels in the study window based on whether a change from one land cover category to another occurred from one product year to another (product years are when imagery is produced for public use).

Recent work in [27] investigated the effects of land cover change (mostly increased urbanization), among other ors, on the presence of *Coccidioides* arthroconidia in air filters placed throughout the Phoenix Metro area. Their approach was to consider the area within a 2.4km radius of air filter sites and calculate the proportion of area that underwent any land cover change. Note that the authors included all possible changes of land cover types in the NLCD raster data. The authors found no major relationship between air prevalence of *Coccidioides* arthroconidia and land cover change. Similarly, the relationship between human clinical cases and air prevalence of *Coccidioides* arthroconidia was also insignificant in that study.

We are interested in the relationship between our subset of disaggregated human cases and certain classes of lander change. Since many individuals can live near multiple sources of land use change, we create a 0 − 1 binary^18^ raster indicating pixels with a 1 if a site experienced relevant land cover change. We then smooth the result using a two-dimensional Gaussian kernel. We expect that such distance effects decay rapidly with distance, so we choose a 1-km bandwidth. The pixels indicated with a 1 for relevant land cover change include those pixels that reflect a change from a non-development category to a development category (i.e. the land cover development categories described in 2.2.4).

### 2.3. Spatial Point Pattern Analysis

We seek to estimate the intensity function of the VF cases over the study window as a function of high-resolution environmental covariate data. Loosely speaking, the intensity function of a spatial point process can be used as a measure of the expected number of cases in any bounded subregion of the study window *W* (including the entirety of *W* itself; see [7], chapter 6, section 6.3 for specifics on the intensity function of spatial point processes). For individuals infected with VF, it may be difficult or impossible to identify the exact spatial location of the exposure that caused the infection. However, the locations of residence of patients infected with VF may offer valuable insight into the spatio-temporal dynamics of the disease. For example, in locations where soil and other environmental conditions are right, nearby soil disturbance due to new construction, dust storms, or other activity may infect multiple individuals in close proximity. Analyzing residential locations as a spatial point pattern allows us to understand the local conditions that increase the likelihood of VF exposure. Among the benefits of this approach is being able to extract covariate effects on disease intensity at a much smaller scale than if data were aggregated, while avoiding the so-called modifiable areal unit problem inherent in spatial data analysis. Treating VF cases as a spatial point pattern allows us to leverage environmental data collected at much finer spatial resolutions, which is especially useful when the spatial distribution of VF cases exhibits high levels of inhomogeneity over relatively small spatial scales (e.g. kilometers).

Our approach involves a 250 × 250 grid of quadrature points over the entire study window, which corresponds to extracting covariate and VF case information in a grid with horizontal and vertical spacing of approximately 0.6 × 0.4 kilometers (i.e. this is the spacing of the quadrature points in the horizontal and vertical directions, respectively). This differs from the approach in [11], where the authors construct the number and location of pseudo-absences based on census block group information in their study window (the entire state of Arizona)^19^ .

By analyzing the VF cases at the point level on the scale of kilometers, we may construct a smoothly varying surface of the intensity of VF cases. Specifically, we treat the VF cases as a spatially inhomogeneous, ‘multitype’ Poisson process. A ‘multitype’ process in our case refers to the categorization of VF cases based on qualitative attributes (or types) of the points called marks; we analyze the pattern as a multivariate spatial point process. In our case, each VF case may have one of four mark categories based on the age and sex of the person infected. The number of Arizona Medicaid patients during each semester of the study period are shown in Figure 4 and Table 1.

**Table 1.** The total number of cases by age and sex category including residences with multiple reported cases. While subpopulations for semester 1, 2013 through semester 1, 2015 are not shown here to protect data privacy, each of these subpopulations include at least 16 people. In parenthesis: the total number of cases used in this analysis (which involves the removal of duplicates, or points with identical attributes and locations; see 2.3). Recall that we define a case as anyone diagnosed with and/or treated for VF. **Source:** Arizona Medicaid (AHCCCS) 2023. CHiR created patient record files from Arizona Medicaid (AHCCCS) data that were used for this analysis.

| Year Sem. | Males Under 45 Years | Females 45 Years or Older | Males 45 Years or Older | Females Under 45 Years | Year/Sem. Total |
| --- | --- | --- | --- | --- | --- |
| 2013 1 | - | - | - | - | 140 (139) |
| 2013 2 | - | - | - | - | 151 (150) |
| 2014 1 | - | - | - | - | 243 (234) |
| 2014 2 | - | - | - | - | 201 (190) |
| 2015 1 | - | - | - | - | 197 (191) |
| 2015 2 | 64 (63) | 216 (212) | 232 (224) | 53 (53) | 565 (552) |
| 2016 1 | 86 (84) | 230 (228) | 196 (186) | 72 (70) | 584 (568) |
| 2016 2 | 74 (73) | 197 (192) | 215 (203) | 80 (80) | 566 (548) |
| 2017 1 | 76 (74) | 214 (207) | 232 (222) | 54 (54) | 576 (557) |
| 2017 2 | 89 (88) | 201 (199) | 225 (212) | 60 (60) | 575 (559) |
| 2018 1 | 87 (83) | 234 (230) | 207 (202) | 92 (92) | 620 (607) |
| 2018 2 | 85 (85) | 183 (181) | 204 (195) | 75 (75) | 547 (536) |
| 2019 1 | 83 (81) | 189 (187) | 195 (187) | 83 (82) | 550 (537) |
| 2019 2 | 124 (119) | 251 (244) | 219 (213) | 114 (112) | 708 (688) |
| 2020 1 | 114 (106) | 206 (200) | 215 (206) | 124 (122) | 659 (634) |
| 2020 2 | 165 (157) | 240 (239) | 250 (244) | 129 (126) | 784 (766) |
| 2021 1 | 164 (152) | 283 (280) | 281 (269) | 129 (128) | 857 (829) |
| 2021 2 | 133 (127) | 256 (254) | 277 (263) | 134 (131) | 800 (775) |
| 2022 1 | 137 (132) | 246 (243) | 214 (199) | 113 (113) | 710 (687) |
| 2022 2 | 133 (126) | 240 (235) | 234 (220) | 120 (120) | 727 (701) |
| 2023 1 | 132 (124) | 244 (241) | 215 (206) | 132 (131) | 723 (702) |
| 2023 2 | 165 (164) | 229 (221) | 289 (279) | 140 (136) | 823 (800) |
| Category Total | 2,030 (1,954) | 4,175 (4106) | 4,279 (4.088) | 1,822 (1802) | 12,306 (11,950) |

**Figure 4:**
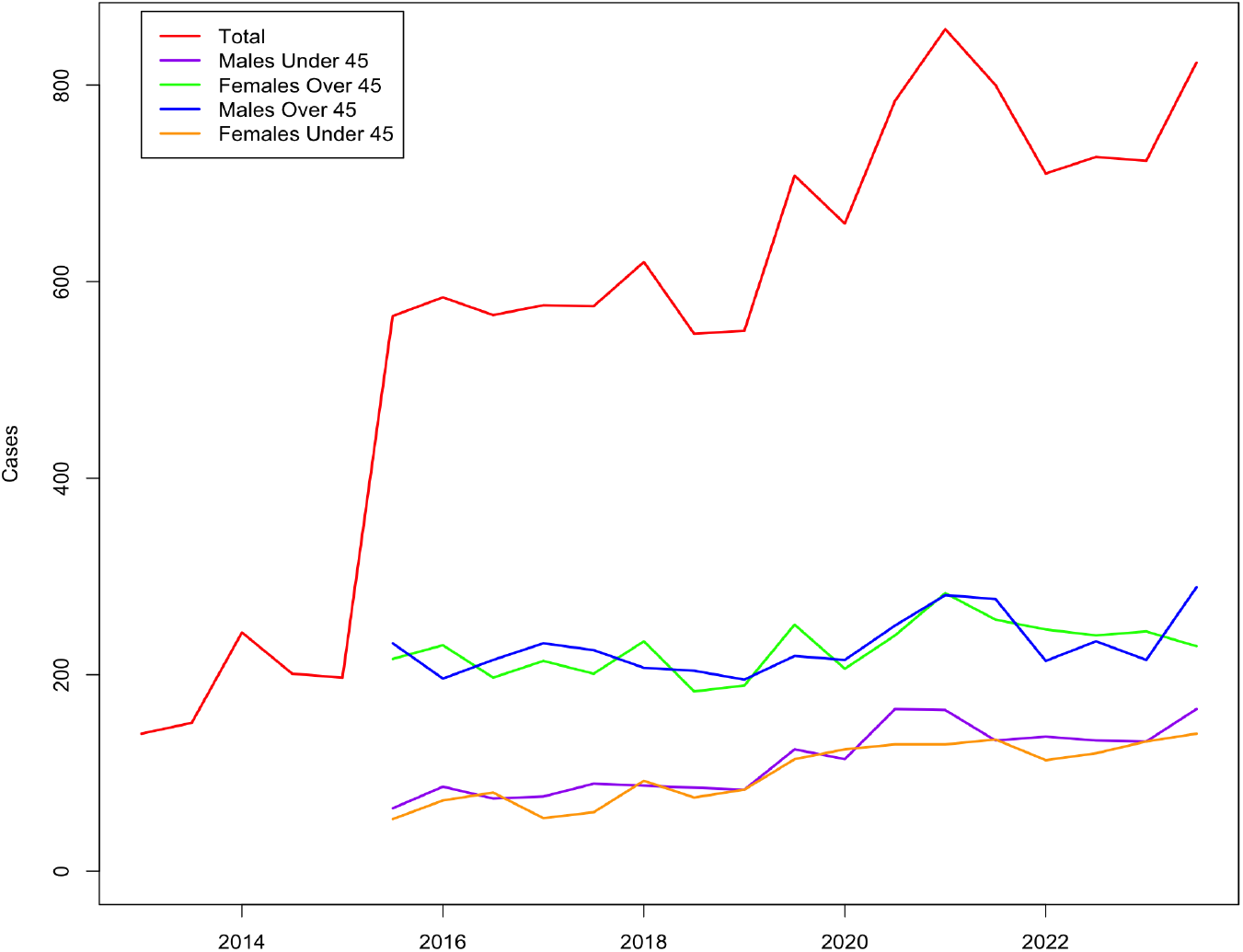
Semesterly VF cases (allowing duplicate residences) among Arizona Medicaid patients during the study period. For age-sex categories, we have removed the first few semesters for privacy. **Source:** AHCCCS 2023. CHiR created patient record files from Arizona Medicaid (AHCCCS) data that were used for this analysis.

The age categories were determined by considering the distribution of VF cases by age as given in the Arizona Department of Health Services ‘Valley fever Annual Report’ [3] over multiple years. It should also be noted that Medicare eligibility, which begins at age 65, likely affects the Medicaid enrollment populations among older individuals.

Duplicate points, where both the mark category and location coincide (e.g. in the case of multiple people infected with VF in the same home), cause a technical violation of the assumption of a spatial Poisson process. Thus, to deal duplicated points, we randomly shuffle the indices of entire data set and take only the first mark category appearing in the shuffled data set at each location.

The MPD estimates vary dramatically within the study window and it is well known that environmental factors influence the abundance of VF cases in Maricopa County [22]. Thus, we model the spatially inhomogeneous intensity of VF cases as a function of environmental and non-environmental covariates. Furthermore, since VF is not communicable among humans, the independence assumption that follows from modeling the VF case as a spatial Poisson process (accounting for the spatially inhomogeneous intensity) is a reasonable one.

For ease of comparison among fitted model coefficients, we standardize (pixel-wise) each non-categorical covariate subtracting the mean pixel value and dividing by the standard deviation of pixel values within the study window. Additionally, to account for uncertainty in the underlying MPD estimates and as a method of facilitating model fitting, we smooth the MPD estimates (SMPD) over the study window using a Gaussian kernel with standard deviation equal to two kilometers. After smoothing MPD, we normalize the SMPD by dividing each pixel by the sum over all pixels in the study window.

For year *i* = 2013, …, 2023 and semester *j* = 1, 2, we specify log-intensity of VF cases for age-sex category *k* = 1, …, 4, denoted by log *λ*_*i,j*_ (*u*, ***θ***, *k*), at spatial location *u* ∈ *W* as a linear function of environmental covariates. Here we let ***θ***≔ (*γ*_*i,j*_, *α*_*i,j*_(*k*), ***β***_*i,j*_) where ***β***_*i,j*_ is a vector of model coefficients with length depending on the number of covariates included in the model and *α*_*i,j*_(*k*) is the age-sex specific intercept term that depends on *k*. An age-sex specific intercept term embeds the assumption that baseline intensities of VF cases for the various age-sex categories differ from one another. This is in contrast to the assumption that the age and sex related effects are also covariate specific, an assumption we do not adopt for sake of parsimony.

We first consider a log-linear base model with only an offset term and age-sex-specific intercept terms based on the marks of the VF cases. This model takes the form

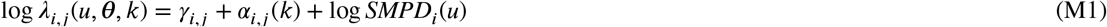

where we once again emphasize that *u* represents a spatial location in the study window *W* .

We build upon the base model by including land cover and land cover change covariates in the linear predictor of log-intensity. The resulting intensity function is given by

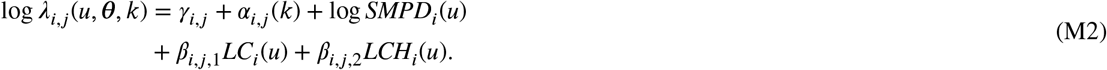

To understand the effect of HSI and NDVI on the intensity of VF cases, we add linear terms for these covariates to the structure of model M2 to get

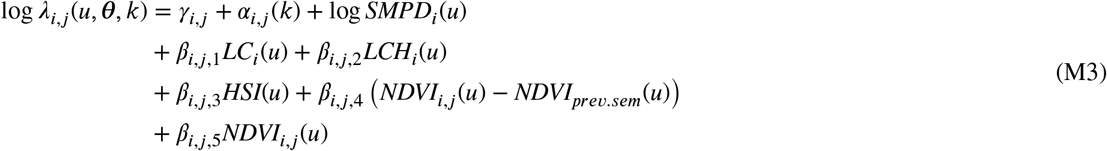

where *NDVI*_*prev.sem*_(*u*) ≔ *NDVI*_*i*−1,*j*+1_(*u*) if *j* = 1 and *NDVI*_*prev.sem*_(*u*) ≔ *NDVI*_*i,j*−1_(*u*) if *j* = 2. Note that for the NDVI difference covariate in 2013, semester 1, we also use the summer-fall average NDVI for 2012, one year before the start of the study period.

Preliminary nonparametric analysis of the relationship between NDVI, HSI, and NDVI difference revealed a nonlinear relationship between these covariates and the intensity of VF cases. Thus, we consider lastly, a log-quadratic (quadratic in HSI and NDVI covariates) model of intensity given by

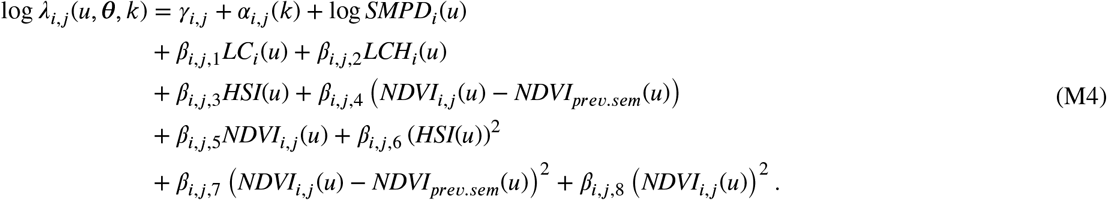

Note that SMPD estimates are used as an offset term to ensure that the dependence of VF cases on covariates is considered after accounting for the spatially varying at-risk population for VF.

The R package spatstat provides a robust toolkit for the analysis of spatial point patterns [8]. Exploratory analysis, statistical inference on the covariates, and model validation were conducted mainly using tools from the spatstat package. We have used the spatstat function ppm to fit the models. We note that we have increased the resolution of the quadrature scheme used in model fitting from its default setting to a 250 × 250^20^ grid of dummy points. Furthermore, our model results involve randomization as we have used the Huang and Ogata model improvement procedure (HOI procedure; see [21]) to obtain more accurate parameter estimates^21^ .

## 3. Model Results

Table 2 displays AIC scores for models M1-M4 for all twenty-two semesters, and estimated coefficients for all four models appear in Tables 6, 5, 4, and 3 located in the Appendix. As expected, the most complex model (M4) outperforms the remaining models in more than half (twelve of twenty-two) of the semesters in terms of Akaike Information Criterion (AIC). Notably, all semesters show improved model fit compared to M1 when incorporating covariates in the model. This is confirmed by likelihood ratio test (LRT) results as shown in Table 7 in the Appendix, which compares models M1 and M4 for all semesters.

**Table 2.** AIC scores for models M1, M2, M3, and M4. Bold values indicate the model with lowest AIC in a given year and semester.

|  | AIC: (M4) | AIC: (M3) | AIC: (M2) | AIC: (M1) |
| --- | --- | --- | --- | --- |
| 2013 1 | **1490.571** | 1501.069 | 1515.979 | 1530.526 |
| 2013 2 | 1614.612 | 1614.078 | **1596.012** | 1651.457 |
| 2014 1 | 2360.409 | 2321.056 | **2320.504** | 2343.830 |
| 2014 2 | **1903.285** | 1936.436 | 1919.132 | 1962.517 |
| 2015 1 | 2034.357 | 1993.157 | **1989.947** | 2010.536 |
| 2015 2 | 4556.189 | 4619.267 | **4545.423** | 4621.829 |
| 2016 1 | **4666.974** | 4751.056 | 4682.002 | 4790.093 |
| 2016 2 | **4691.444** | 4752.454 | 4786.152 | 4830.226 |
| 2017 1 | 4655.598 | 4637.935 | **4635.077** | 4698.978 |
| 2017 2 | 4740.715 | 4748.753 | **4695.288** | 4787.417 |
| 2018 1 | **5081.113** | 5111.509 | 5101.664 | 5217.765 |
| 2018 2 | **4526.751** | 4593.108 | 4579.467 | 4671.321 |
| 2019 1 | **4501.442** | 4571.224 | 4533.488 | 4642.235 |
| 2019 2 | **5384.942** | 5542.344 | 5498.665 | 5610.138 |
| 2020 1 | **5246.709** | 5297.711 | 5283.889 | 5386.455 |
| 2020 2 | **6063.660** | 6099.828 | 6089.066 | 6222.545 |
| 2021 1 | **6584.133** | 6621.179 | 6665.665 | 6727.698 |
| 2021 2 | 6171.841 | **6154.894** | 6220.150 | 6313.311 |
| 2022 1 | 5639.890 | **5633.005** | 5654.852 | 5761.771 |
| 2022 2 | 5684.374 | 5662.000 | **5637.197** | 5790.466 |
| 2023 1 | **5669.598** | 5709.332 | 5706.816 | 5840.440 |
| 2023 2 | 6294.313 | 6359.979 | **6270.750** | 6392.168 |

**Table 3.** M4 results.

|  | Intercept | Fem>=45 | Male>=45 | Fem<45 | Dev Open | Dev LI | Dev MI | Dev HI |
| --- | --- | --- | --- | --- | --- | --- | --- | --- |
| 2013 1 | 0.602 | 1.398*** | 1.1427** | 0.2801 | 1.6334** | 2.3681*** | 3.5824*** | 1.7065** |
| 2013 2 | -0.25 | 1.0248*** | 0.8222** | -0.1181 | -11.843*** | 3.6217*** | 4.8472*** | 4.034*** |
| 2014 1 | 1.903*** | 1.4213*** | 1.5162*** | 0.0139 | 0.1987 | 0.8495* | 2.2616*** | 0.0254 |
| 2014 2 | 1.088* | 0.5604* | 1.075*** | -0.16 | 1.9331*** | 2.643*** | 3.4497*** | 1.5589** |
| 2015 1 | 0.919* | 1.221*** | 1.2808*** | 0.2102 | 1.744*** | 1.7438*** | 3.3956*** | 1.0641* |
| 2015 2 | 2.113*** | 1.2998*** | 1.4654*** | -0.2601 | 2.097*** | 1.4182*** | 2.8135*** | 0.5385 |
| 2016 1 | 2.299*** | 1.0789*** | 0.9596*** | 0.0874 | 1.5533*** | 2.2433*** | 3.2416*** | 0.7513** |
| 2016 2 | 1.358*** | 1.1638*** | 1.1993*** | 0.0615 | 2.5412*** | 3.1337*** | 3.8323*** | 2.122*** |
| 2017 1 | 2.084*** | 1.0124*** | 1.1046*** | -0.4055 | 1.6293*** | 2.3855*** | 3.1276*** | 1.3996*** |
| 2017 2 | 2.956*** | 0.5765*** | 0.763*** | -0.5095** | 1.2105*** | 1.5539*** | 2.4789*** | 0.2897 |
| 2018 1 | 1.93*** | 1.2105*** | 0.8984*** | -0.0667 | 1.8386*** | 2.4908*** | 3.1298*** | 0.8675*** |
| 2018 2 | 2.492*** | 0.5343*** | 0.6155*** | -0.3238* | 1.8884*** | 2.2033*** | 2.6149*** | 0.5903* |
| 2019 1 | 1.882*** | 0.8628*** | 0.9172*** | -0.0078 | 1.2781*** | 1.873*** | 2.6499*** | -0.5019 |
| 2019 2 | 2.637*** | 0.6239*** | 0.5949*** | -0.3257* | 0.8358** | 1.8621*** | 2.6324*** | 0.5177* |
| 2020 1 | 3.442*** | 0.7389*** | 0.7211*** | 0.2495 | -0.3811 | 0.729*** | 1.6245*** | -0.1567 |
| 2020 2 | 2.385*** | 0.471*** | 0.6116*** | 0.0638 | 2.3353*** | 2.1223*** | 2.7974*** | 1.0828*** |
| 2021 1 | 3.255*** | 0.5339*** | 0.5062*** | -0.2114 | 1.3537*** | 2.0718*** | 2.5558*** | 0.4322 |
| 2021 2 | 2.95*** | 0.6906*** | 0.6075*** | -0.1635 | 1.5052*** | 2.0223*** | 2.7231*** | -0.0415 |
| 2022 1 | 1.631*** | 0.8558*** | 0.5676*** | -0.2883 | 3.0608*** | 3.749*** | 3.9127*** | 2.0063*** |
| 2022 2 | 1.606*** | 0.9093*** | 0.7859*** | 0.2326 | 2.6119*** | 3.2225*** | 4.2006*** | 2.6983*** |
| 2023 1 | 1.839*** | 0.5147*** | 0.2824* | -0.0181 | 2.4427*** | 3.5093*** | 3.9838*** | 2.3257*** |
| 2023 2 | 3.733*** | 0.1273 | 0.4203*** | -0.4754*** | 0.3282 | 1.1656*** | 1.9714*** | -0.0289 |
|  | Land Ch | NDVI Cur | (NDVI Cur)^2 | HIS | (HSI)^2 | NDVI Diff | (NDVI Diff)^2 |  |
| 2013 1 | 56.292 | 0.1555 | -0.8503*** | 0.6243*** | -0.2739 | -0.0269 | -0.8561*** |  |
| 2013 2 | 80.561*** | 0.4972** | -0.913*** | 0.1721 | -0.1287 | -0.4098* | -0.6177*** |  |
| 2014 1 | 31.626 | -0.3821** | -0.4637*** | 0.1818 | 0.0409 | -0.2337 | -0.4966*** |  |
| 2014 2 | 38.847* | 0.3887** | -0.6762*** | 0.1912 | 0.002 | 0.0736 | -0.872*** |  |
| 2015 1 | 25.78 | 0.0381 | -0.0831 | 0.2274* | -0.4845*** | 0.0771 | -0.0949*** |  |
| 2015 2 | 22.458* | 0.346*** | -0.4485*** | 0.1286* | 0.0354 | 0.0689 | -0.079*** |  |
| 2016 1 | 23.062* | 0.0788 | -0.3779*** | 0.3303*** | -0.2754*** | 0.1758* | -0.4224*** |  |
| 2016 2 | 45.537*** | 0.257** | -0.5367*** | 0.2279*** | -0.1086 | -0.3587*** | -0.0128 |  |
| 2017 1 | 51.058*** | -0.227** | -0.1699*** | 0.4047*** | -0.2396*** | 0.1376 | 0.0061 |  |
| 2017 2 | 34.564*** | 0.0205 | -0.4824*** | 0.0986 | 0.0517 | 0.5827*** | -0.394*** |  |
| 2018 1 | 46.073*** | -0.0437 | -0.5553*** | 0.2815*** | 0.1667** | 0.4694*** | -0.0074 |  |
| 2018 2 | 30.41*** | -0.1074 | -0.3278*** | 0.276*** | -0.2213** | -1.1068*** | -0.277*** |  |
| 2019 1 | 23.795*** | -0.6186*** | -0.2567*** | 0.3506*** | -0.0181 | -1.044*** | -0.632*** |  |
| 2019 2 | 19.877*** | -0.0795 | -0.4817*** | 0.3606*** | -0.0479 | 0.6988*** | -0.1086*** |  |
| 2020 1 | 32.728*** | -0.2719*** | -0.4455*** | 0.2238*** | -0.1088 | -1.0807*** | -0.6443*** |  |
| 2020 2 | 39.136*** | -0.1062 | -0.4686*** | 0.3457*** | -0.0149 | 1.0133*** | -0.3224*** |  |
| 2021 1 | 31.842*** | 0.0968 | -0.4792*** | 0.2003*** | 0.0979* | 0.3192*** | -0.0634*** |  |
| 2021 2 | 20.34*** | -0.1869** | -0.27*** | 0.1814*** | -0.1271* | -0.1875* | -0.2914*** |  |
| 2022 1 | 58.477*** | 0.0369 | -0.4304*** | 0.1506** | 0.0363 | 0.8011*** | -0.3773*** |  |
| 2022 2 | 65.558*** | 0.2166** | -0.5071*** | 0.0907 | -0.0951 | -0.6878*** | -0.3*** |  |
| 2023 1 | 25.755*** | -0.2309*** | -0.4066*** | 0.2442*** | 0.0123 | 0.1983* | -0.1418*** |  |
| 2023 2 | 11.763* | 0.1562* | -0.4872*** | 0.1474** | -0.0398 | 1.1641*** | -0.4785*** |  |

**Table 4.** M3 results.

|  | Intercept | Fem>=45 | Male>=45 | Fem<45 | LCC1: Dev Open | LCC2: Dev LI |
| --- | --- | --- | --- | --- | --- | --- |
| 2013 1 | 0.0992 | 1.0816*** | 0.9343*** | -0.063 | 1.5506** | 2.4641*** |
| 2013 2 | -1.6574*** | 1.2834*** | 1.2962*** | 0.2982 | -11.4666*** | 3.9071*** |
| 2014 1 | 1.5113*** | 1.0415*** | 1.1018*** | -0.1624 | 1.0299* | 1.5917*** |
| 2014 2 | -0.6544 | 0.631* | 1.1942*** | -0.3673 | 2.5838*** | 3.5621*** |
| 2015 1 | 0.6805 | 0.9618*** | 1.1584*** | -0.1091 | 1.8519*** | 1.9468*** |
| 2015 2 | 2.1062*** | 1.4148*** | 1.1607*** | -0.1419 | 1.2713*** | 1.6265*** |
| 2016 1 | 1.7555*** | 1.1626*** | 1.0642*** | 0.0743 | 1.6316*** | 2.3682*** |
| 2016 2 | 1.4618*** | 0.7962*** | 0.8589*** | -0.0798 | 2.4394*** | 3.0696*** |
| 2017 1 | 1.7595*** | 0.9107*** | 1.0972*** | -0.2232 | 1.6034*** | 2.314*** |
| 2017 2 | 1.8964*** | 0.9353*** | 0.9751*** | -0.3439* | 1.3994*** | 2.2327*** |
| 2018 1 | 2.0173*** | 0.8252*** | 0.6817*** | -0.0079 | 1.3328*** | 2.3376*** |
| 2018 2 | 1.9117*** | 0.7345*** | 0.778*** | -0.1788 | 1.8704*** | 2.1945*** |
| 2019 1 | 1.0771*** | 0.9242*** | 0.6606*** | -0.2542 | 1.9263*** | 3.1992*** |
| 2019 2 | 1.975*** | 0.7452*** | 0.5702*** | -0.1504 | 1.1045*** | 2.4038*** |
| 2020 1 | 2.5624*** | 0.5864*** | 0.5859*** | 0.0541 | 0.493 | 1.7925*** |
| 2020 2 | 1.8196*** | 0.3932*** | 0.448*** | -0.1381 | 2.2812*** | 2.7281*** |
| 2021 1 | 2.6587*** | 0.6073*** | 0.6507*** | -0.2748* | 1.5693*** | 2.0875*** |
| 2021 2 | 2.5532*** | 0.6355*** | 0.6274*** | -0.0468 | 1.4593*** | 2.3363*** |
| 2022 1 | 1.2879*** | 0.5931*** | 0.3023* | -0.2457 | 3.0827*** | 3.7845*** |
| 2022 2 | 1.1601*** | 0.6984*** | 0.602*** | -0.0449 | 2.404*** | 3.5418*** |
| 2023 1 | 1.2171*** | 0.5658*** | 0.388** | -0.1121 | 2.28*** | 3.8005*** |
| 2023 2 | 2.5143*** | 0.2635* | 0.4249*** | -0.1573 | 0.3404 | 2.2005*** |
|  | LCC3: Dev MI | LCC4: Dev HI | Land Ch | NDVI Cur | HSI | NDVI Diff |
| 2013 1 | 3.6874*** | 1.2542* | 61.2137 | -0.0156 | 0.3468** | 0.334* |
| 2013 2 | 5.2736*** | 3.5621*** | 71.5796*** | -0.0354 | 0.1007 | -0.3547* |
| 2014 1 | 2.6966*** | 0.1032 | 20.1026 | -0.3505*** | 0.2971** | 0.3274** |
| 2014 2 | 4.6541*** | 1.4326** | 55.6534** | 0.0024 | 0.199 | -0.1935 |
| 2015 1 | 3.3045*** | 1.1716** | 18.1605 | 0.0057 | 0.1429 | 0.0815 |
| 2015 2 | 2.8391*** | 0.0665 | -5.322 | -0.1754** | 0.2202*** | -0.0424 |
| 2016 1 | 3.388*** | 0.3693 | 22.2503* | -0.212*** | 0.3361*** | 0.3038*** |
| 2016 2 | 3.8619*** | 1.8207*** | 42.2528*** | -0.1098 | 0.2261** | -0.5349*** |
| 2017 1 | 3.3065*** | 1.6896*** | 37.8675*** | -0.1452* | 0.33*** | 0.2121* |
| 2017 2 | 3.1628*** | 0.736** | 30.9184*** | -0.3107*** | 0.2035*** | 0.0369 |
| 2018 1 | 3.1707*** | 0.6035* | 38.7866*** | -0.2458*** | 0.2099** | 0.2688*** |
| 2018 2 | 2.9144*** | 0.482* | 38.6607*** | -0.2338*** | 0.2999*** | -0.5054*** |
| 2019 1 | 3.8359*** | 0.3035 | 22.0072*** | -0.5197*** | 0.4278*** | 0.0991 |
| 2019 2 | 3.4376*** | 0.457 | 14.0296*** | -0.2775*** | 0.4097*** | 0.2377** |
| 2020 1 | 2.6435*** | 0.4169 | 24.1469*** | -0.2575*** | 0.1953*** | 0.0321 |
| 2020 2 | 3.5354*** | 1.5059*** | 33.9831*** | -0.21*** | 0.3348*** | 0.3084*** |
| 2021 1 | 2.8703*** | 0.3807 | 27.7633*** | -0.1954*** | 0.2095*** | -0.0128 |
| 2021 2 | 2.754*** | 0.2797 | 24.377*** | -0.3296*** | 0.0987 | 0.0137 |
| 2022 1 | 4.1128*** | 2.1503*** | 49.742*** | -0.1213* | 0.1562** | 0.4512*** |
| 2022 2 | 4.3937*** | 2.3734*** | 33.2426** | -0.0171 | 0.1818** | -0.3827*** |
| 2023 1 | 4.391*** | 2.0144*** | 20.721*** | -0.0273 | 0.1997*** | 0.1897* |
| 2023 2 | 3.1163*** | 0.5515* | 17.2684** | -0.0431 | 0.0756 | 0.2525*** |

**Table 5.** M2 results.

|  | Intercept | Fem>=45 | Male>=45 | Fem<45 | LCC1: Dev Open | LCC2: Dev LI |
| --- | --- | --- | --- | --- | --- | --- |
| 2013 1 | -0.1044 | 1.1903*** | 1.0555*** | 0.2676 | 0.9464 | 2.2443*** |
| 2013 2 | -0.9713* | 0.9203** | 1.0018** | -0.1111 | -12.4111*** | 3.5004*** |
| 2014 1 | 0.812* | 1.2865*** | 1.3672*** | 0.1211 | 0.7769* | 1.7048*** |
| 2014 2 | -0.327 | 0.9236*** | 1.2674*** | 0.1193 | 1.768*** | 3.2271*** |
| 2015 1 | 0.6952 | 0.8613*** | 1.0083*** | -0.126 | 1.7804*** | 1.9472*** |
| 2015 2 | 1.8373*** | 1.1858*** | 1.1779*** | -0.174 | 1.6895*** | 1.8642*** |
| 2016 1 | 1.6978*** | 0.9712*** | 0.7145*** | -0.1843 | 1.3724*** | 2.232*** |
| 2016 2 | 1.1448*** | 1.0719*** | 1.245*** | 0.4429* | 1.7814*** | 2.588*** |
| 2017 1 | 1.9113*** | 0.8207*** | 0.9869*** | -0.2375 | 1.1861*** | 2.145*** |
| 2017 2 | 1.8505*** | 0.7713*** | 0.8489*** | -0.4208** | 1.6499*** | 2.4166*** |
| 2018 1 | 1.6741*** | 1.2391*** | 1.0221*** | 0.1988 | 1.2935*** | 2.3346*** |
| 2018 2 | 1.9223*** | 0.7351*** | 0.8123*** | -0.1473 | 1.6922*** | 2.4239*** |
| 2019 1 | 1.0902*** | 0.6929*** | 0.7793*** | -0.1122 | 1.7567*** | 3.1404*** |
| 2019 2 | 2.1279*** | 0.6151*** | 0.4563*** | -0.1041 | 0.9007*** | 2.4089*** |
| 2020 1 | 2.0395*** | 0.7623*** | 0.8725*** | 0.3018* | 0.5163* | 2.1615*** |
| 2020 2 | 2.2345*** | 0.422*** | 0.5374*** | -0.4078*** | 1.7866*** | 2.3368*** |
| 2021 1 | 2.1434*** | 0.7875*** | 0.5772*** | -0.0613 | 1.8187*** | 2.6809*** |
| 2021 2 | 2.1914*** | 0.5468*** | 0.5288*** | -0.113 | 1.2509*** | 2.68*** |
| 2022 1 | 1.3401*** | 0.6176*** | 0.4274** | -0.1945 | 2.3677*** | 3.3809*** |
| 2022 2 | 2.2525*** | 0.6183*** | 0.581*** | -0.1889 | 1.1947*** | 2.199*** |
| 2023 1 | 1.0382*** | 0.5721*** | 0.3747** | -0.0498 | 2.3458*** | 3.6507*** |
| 2023 2 | 2.8011*** | 0.0975 | 0.4828*** | -0.2985* | 0.5425* | 2.1918*** |
|  | LCC3: Dev MI | LCC4: Dev HI | Land Ch |  |  |  |
| 2013 1 | 3.6991*** | 1.2126* | 63.8399 |  |  |  |
| 2013 2 | 4.6899*** | 3.0909*** | 64.0815*** |  |  |  |
| 2014 1 | 3.1092*** | 0.6862 | 26.0356 |  |  |  |
| 2014 2 | 4.1012*** | 1.4211** | 46.1408** |  |  |  |
| 2015 1 | 3.3359*** | 0.8796* | 14.3497 |  |  |  |
| 2015 2 | 3.0031*** | 0.7468** | 24.1879** |  |  |  |
| 2016 1 | 3.4325*** | 1.1208*** | 37.5759*** |  |  |  |
| 2016 2 | 3.6904*** | 1.6164*** | 36.3559*** |  |  |  |
| 2017 1 | 3.0607*** | 1.2883*** | 48.9081*** |  |  |  |
| 2017 2 | 3.2608*** | 1.1848*** | 25.8847** |  |  |  |
| 2018 1 | 3.1883*** | 0.897*** | 41.1538*** |  |  |  |
| 2018 2 | 3.1182*** | 0.8994*** | 42.631*** |  |  |  |
| 2019 1 | 4.0873*** | 1.2144*** | 21.7399*** |  |  |  |
| 2019 2 | 3.457*** | 0.889*** | 19.115*** |  |  |  |
| 2020 1 | 3.0348*** | 1.0112*** | 32.5382*** |  |  |  |
| 2020 2 | 3.2983*** | 1.4803*** | 34.5728*** |  |  |  |
| 2021 1 | 3.3915*** | 1.0976*** | 38.7192*** |  |  |  |
| 2021 2 | 3.3944*** | 0.5352** | 24.9*** |  |  |  |
| 2022 1 | 4.0184*** | 1.9804*** | 52.7634*** |  |  |  |
| 2022 2 | 3.2173*** | 1.22*** | 27.4793* |  |  |  |
| 2023 1 | 4.3949*** | 2.2371*** | 19.5177*** |  |  |  |
| 2023 2 | 2.8946*** | 0.8094*** | 20.3694*** |  |  |  |

**Table 6.** M1 results.

|  | Intercept | Fem>=45 | Male>=45 | Fem<45 |
| --- | --- | --- | --- | --- |
| 2013 1 | 2.8168*** | 1.226*** | 1.0328** | 0.0747 |
| 2013 2 | 2.9566*** | 1.0101*** | 1.0948*** | 0.0481 |
| 2014 1 | 3.3989*** | 1.0485*** | 1.0692*** | -0.1088 |
| 2014 2 | 3.21*** | 0.7543** | 1.2392*** | 0.0382 |
| 2015 1 | 3.3383*** | 0.8** | 0.9966*** | -0.0423 |
| 2015 2 | 4.1346*** | 1.2182*** | 1.2785*** | -0.1636 |
| 2016 1 | 4.4283*** | 1.0044*** | 0.7954*** | -0.1459 |
| 2016 2 | 4.2864*** | 0.9707*** | 1.0245*** | 0.1003 |
| 2017 1 | 4.3203*** | 1.0149*** | 1.0809*** | -0.3334 |
| 2017 2 | 4.4799*** | 0.7936*** | 0.8849*** | -0.3922* |
| 2018 1 | 4.3667*** | 1.0598*** | 0.9411*** | 0.1361 |
| 2018 2 | 4.4296*** | 0.765*** | 0.8418*** | -0.1238 |
| 2019 1 | 4.3995*** | 0.8222*** | 0.8433*** | 0.0038 |
| 2019 2 | 4.7801*** | 0.7122*** | 0.5954*** | -0.0705 |
| 2020 1 | 4.6507*** | 0.6284*** | 0.6653*** | 0.1519 |
| 2020 2 | 5.0635*** | 0.4185*** | 0.4416*** | -0.2212 |
| 2021 1 | 5.0083*** | 0.6115*** | 0.5708*** | -0.165 |
| 2021 2 | 4.8429*** | 0.7032*** | 0.7268*** | 0.0254 |
| 2022 1 | 4.8872*** | 0.5939*** | 0.4147*** | -0.1614 |
| 2022 2 | 4.8158*** | 0.6492*** | 0.5726*** | -0.0343 |
| 2023 1 | 4.8215*** | 0.6723*** | 0.5095*** | 0.0552 |
| 2023 2 | 5.1024*** | 0.2949* | 0.5291*** | -0.192 |

**Table 7.** Likelihood ratio test results comparing models M4 and M1 for data in all semesters.

|  | Deviance | Pr.Chi.grt | Significance |
| --- | --- | --- | --- |
| 2013 1 | -91.92962 | 6.97E-15 | *** |
| 2013 2 | -106.11369 | 1.09E-17 | *** |
| 2014 1 | -102.32279 | 6.19E-17 | *** |
| 2014 2 | -107.07977 | 7.01E-18 | *** |
| 2015 1 | -64.69665 | 1.23E-09 | *** |
| 2015 2 | -196.17208 | 4.63E-36 | *** |
| 2016 1 | -255.7314 | 1.76E-48 | *** |
| 2016 2 | -223.75343 | 8.53E-42 | *** |
| 2017 1 | -166.22895 | 7.04E-30 | *** |
| 2017 2 | -213.64009 | 1.09E-39 | *** |
| 2018 1 | -267.02563 | 7.53E-51 | *** |
| 2018 2 | -232.45647 | 1.30E-43 | *** |
| 2019 1 | -268.34043 | 3.99E-51 | *** |
| 2019 2 | -324.50412 | 5.94E-63 | *** |
| 2020 1 | -244.08938 | 4.82E-46 | *** |
| 2020 2 | -309.22889 | 9.94E-60 | *** |
| 2021 1 | -268.66618 | 3.41E-51 | *** |
| 2021 2 | -268.97535 | 2.94E-51 | *** |
| 2022 1 | -260.21749 | 2.02E-49 | *** |
| 2022 2 | -289.93618 | 1.15E-55 | *** |
| 2023 1 | -256.785 | 1.06E-48 | *** |
| 2023 2 | -282.72423 | 3.79E-54 | *** |

**Table 8.** Likelihood ratio test results comparing models M4 and M3 for data in all semesters.

|  | Deviance | Pr.Chi.grt | Significance |
| --- | --- | --- | --- |
| 2013 1 | -29.93174 | 1.43E-06 | *** |
| 2013 2 | -27.10015 | 5.61E-06 | *** |
| 2014 1 | -21.33266 | 8.98E-05 | *** |
| 2014 2 | -32.21236 | 4.72E-07 | *** |
| 2015 1 | -5.16872 | 1.60E-01 |  |
| 2015 2 | -38.2708 | 2.48E-08 | *** |
| 2016 1 | -53.19729 | 1.66E-11 | *** |
| 2016 2 | -48.10531 | 2.02E-10 | *** |
| 2017 1 | -13.86293 | 3.10E-03 | ** |
| 2017 2 | -50.67361 | 5.74E-11 | *** |
| 2018 1 | -55.17654 | 6.30E-12 | *** |
| 2018 2 | -54.3936 | 9.25E-12 | *** |
| 2019 1 | -31.73858 | 5.94E-07 | *** |
| 2019 2 | -58.61812 | 1.16E-12 | *** |
| 2020 1 | -59.47836 | 7.60E-13 | *** |
| 2020 2 | -72.30227 | 1.37E-15 | *** |
| 2021 1 | -56.57793 | 3.16E-12 | *** |
| 2021 2 | -43.03796 | 2.42E-09 | *** |
| 2022 1 | -57.79082 | 1.74E-12 | *** |
| 2022 2 | -70.662 | 3.08E-15 | *** |
| 2023 1 | -36.71785 | 5.28E-08 | *** |
| 2023 2 | -78.63098 | 6.03E-17 | *** |

### 3.1. HSI Results

Since we treat the residential address of VF cases as a marked point pattern, it is useful to interpret the results of each model as providing four separate spatial point patterns based on age and sex category. By incorporating age-sex specific intercept terms, we are estimating an increase or decrease in the baseline intensity for that specific age-sex category with respect to the base category of males under 45 years of age.

Among individuals over 45 years of age (for both sexes), estimated VF intensity is greater relative to that of males under 45 years. No clear pattern exists in sign or statistical significance for the estimated intercept term of the intensity of VF cases among females under 45 years relative to males under 45. Our results suggest that, while there is a greater intensity of VF cases among individuals over 45 years of age of both sexes relative to the intensity of males under 45 years, no such pattern exists regarding the intensities of VF cases males and females. It is worth highlighting that the intercept terms for males and females over 45 years seem to be larger for earlier semesters when compared to later semesters; this holds for all models considered. This may be due to changing Medicaid enrollment standards and demographic shifts during the study period.

To understand how all environmental factors together affect the intensity of VF cases, we focus on the interpretation results for models M3 and M4. Recall that we have standardized the continuous covariates for ease of model implementation. Thus, for model M3, an increase by one standard deviation of the coefficient for a standardized, continuous covariate (e.g. HSI, NDVI, or NDVI difference) is associated with a change in the intensity of VF cases by a factor of 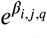 per square kilometer, where *β*_*i,j,q*_ is the associated parameter estimate and *q* = 1, …, 5. For model M4, a unit change in the standard deviation of the continuous covariate depends on the value of the estimated coefficients and a particular value of the covariate^22^. For example, in semester 1 of 2013, one standard deviation increase in HIS is estimated to increase the intensity of VF cases by a factor of 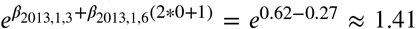 or by 41% per square kilometer, when calculated at the mean value of HSI and holding all other covariates constant. In 2023, using the same procedure, the resulting factor of increase on VF incidence is *e*^0.147−0.04^ ≈ 1.11 (an 11% increase), all other factors held constant. Figure 6 shows the marginal effects (ME) from model M4 for the continuous covariates for semester 1 of 2020 and semester 1 of 2021 when the values of the other covariates (and population offset) are fixed at specified values or categories. Note that the different shape of the HSI ME curves (in the rightmost column of Figure 6) for the two semesters is due to differing signs for the quadratic terms in those semesters. For most semesters, there is greater uncertainty around the predicted intensity for the values of HSI above about 0.7. This is clear in the predicted intensity curves shown in Figure 6. This may be due to limited Medicaid population and observed VF cases in locations with the highest HSI despite our using an estimated Medicaid population as a model offset. Marginal effect curves for all semesters/years are available in the supplementary file.

Estimated coefficients for HSI, NDVI, NDVI difference, and their quadratic terms over time are given in Figure 5. Our results suggest inconsistent coefficient sign and statistical significance patterns for the quadratic HSI terms in model M4 (see Figure 5). For models M4 and M3 the linear coefficients for HSI were positive and significant at the 95% level or greater in seventeen of the twenty-two semesters, revealing consistent evidence for a log-linear relationship between HSI and VF incidence. Only in seven semesters do we find significant evidence (at the 95% level of significance or greater) of a log-quadratic effect between HSI and VF incidence. In six of those seven semesters where we observed significant log-quadratic effects, the quadratic terms were negative, suggesting a diminishing positive effect on VF incidence for higher values of HSI. The magnitude of the linear coefficients associated with HSI in model M4 appear to be relatively stable over the study period compared to the coefficients of NDVI and NDVI difference (see the rightmost column of Figure 5). Results from models M4 and M3 suggest that, among Medicaid patients, soil properties associated with higher HSI near a residence can increase VF risk.

**Figure 5:**
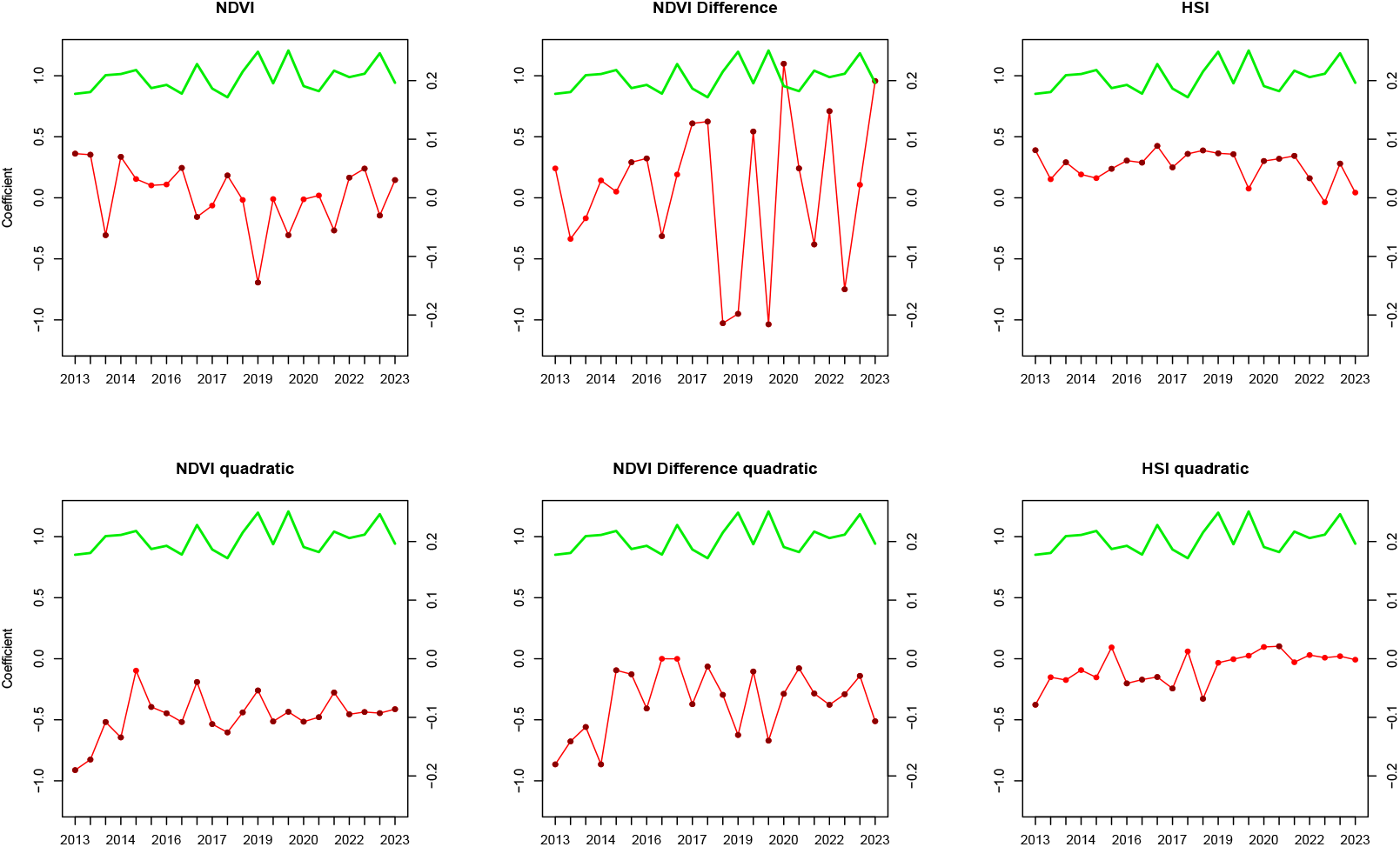
Time series of coefficient estimates from model M4 for all semesters (red lines). The secondary vertical axis pertains to the pixel-wise average NDVI (green curve). Dark red points indicate semesters where the coefficient was significant at the 95% level or greater (see Table 3). Dark red points indicate semesters where the coefficient was not significant at this level.

### 3.2. NDVI Results

Figure 5 reveals an apparent relationship between the average NDVI calculated pixel-wise for each semester and the trajectory in the estimated coefficients for NDVI and differenced NDVI over the study period. The pattern suggests that when the average NDVI is higher, the estimated log-linear effect of NDVI on VF incidence is decreased. This may be due to patterns of bimodal seasonality in all observed VF cases and how our Medicaid VF data are temporally aggregated (see [22]). The changing signs for the NDVI and NDVI difference coefficients are also likely related to the dynamic effect of NDVI on VF incidence as discussed in [28]; the authors note that higher NDVI measured up to (even slightly over a year) prior can have a positive effect on VF incidence. Despite the dynamic nature of the relationship between NDVI and VF incidence, the negative and consistently significant quadratic terms in the model suggest that a nonlinear relationship is more appropriate with lower VF incidence at the highest values of NDVI and NDVI difference. The most pronounced effect on predicted VF intensity occurs at values of NDVI within 0.1 − 0.3, as can be seen in the left panel of Figure 6. This pattern holds for the ME curves in other semesters as well.

**Figure 6:**
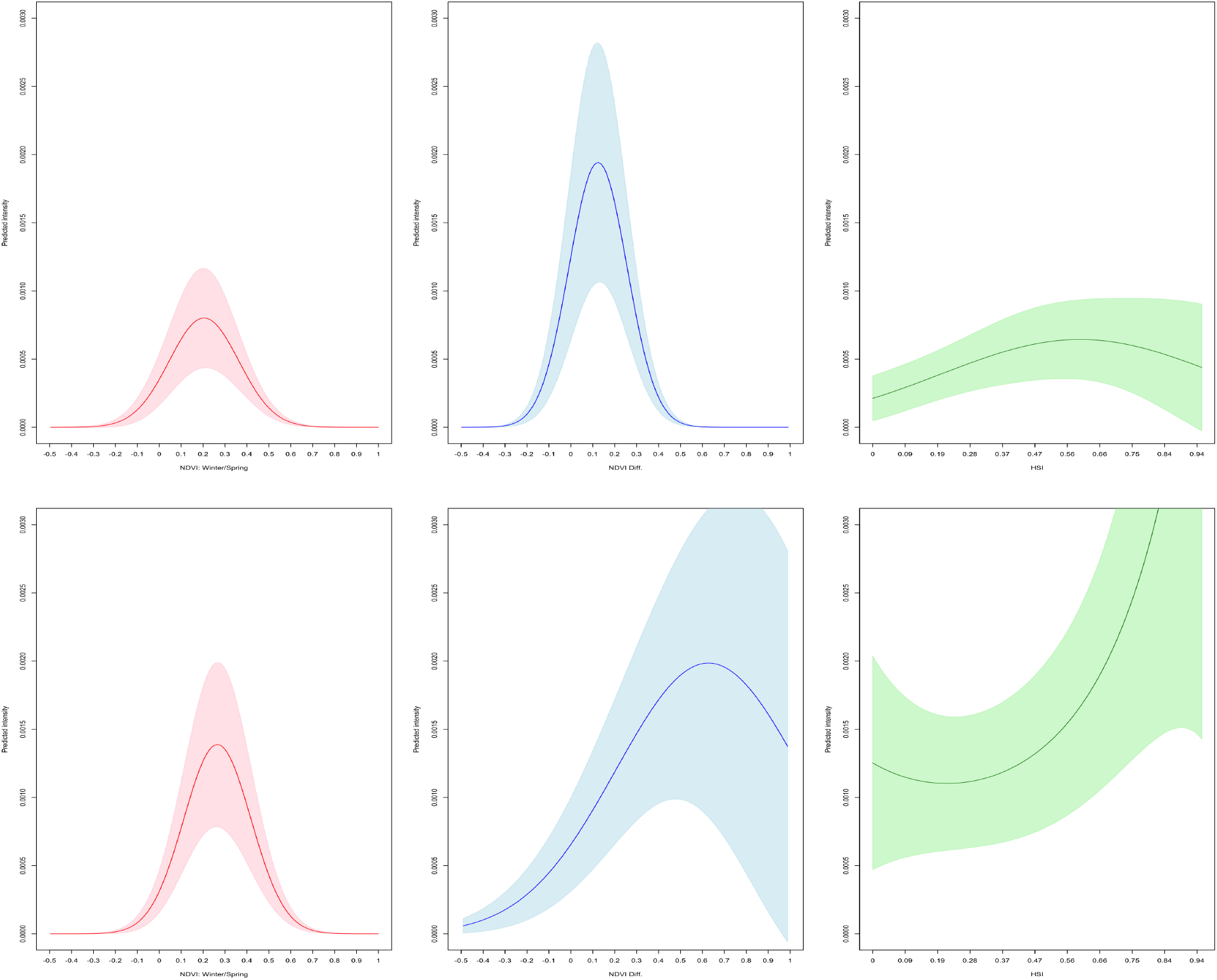
Marginal effect of NDVI (left column), NDVI difference (middle column) and HSI (right column) on intensity of VF cases based on model M4: 2020 semester 1 (top row) and 2021 semester 1 (bottom row). Effects are calculated by setting other continuous covariates to their 0.75 quantile. Land cover is fixed at the base category “Other”. Shaded regions are 95% confidence bands.

Another interesting pattern in Figure 5 is the increase in the volatility of estimates for the coefficient of NDVI difference following the second semester of 2017 for model M4. During the same period, it can be observed that the coefficients are statistically significant at the 95% level or greater for all semesters except the first semester of 2023. While a notable increase in all VF cases (i.e. not restricted to those diagnosed with VF who are enrolled in Medicaid) was observed in Maricopa county during this same period [9], no such spike appears within the number of cases used in this study (see Table 4). An alternative explanation is that volatility in environmental factors that affect NDVI (e.g. precipitation and temperature) are interacting in ways not accounted for here.

### 3.3. Land Cover and Land Cover Change Results

Recall that for the factor-valued land cover covariate, the coefficient *β* reflects the intensity of VF cases per square kilometer for each category relative to the base category “Other”, which is treated as having coefficient *β* = 0.

We now consider model M4 results specifically. In most semesters development of all types had positive and statistically significant (at the 95% level or greater) effects on VF incidence when compared with the base land cover category “Other”. In particular, relatively large positive estimated effects are observed in every semester for LCC2 and LCC3 compared to the base category “Other”. These comparatively large estimated effects for all LCCs may be due to Medicaid population-related factors associated with areas of low and medium intensity development. For example, based on LCC2 estimates, areas with LCC2 had VF incidence ranging from *e*^0.729^ = 2.073 (in 2020 semester 1) to *e*^3.749^ = 42.479 (2022 semester 1) times the VF incidence in areas classified as the base land cover category “Other”.

An even more pronounced variation holds for LCC3 estimates while for LCC4, even when considering only semesters where the estimated effect was statistically significant at the 95% level or greater, the estimated effect on VF incidence varied from *e*^0.517^ = 1.678 (2019 semester 2) to *e*^2.698^ = 14.855 (2022 semester 2) times that of VF incidence in the base land cover category “Other”. The variation in estimates for the different LCCs in this time period in particular could be due to a changing Medicaid population during the COVID-19 pandemic that is not accounted for in Census estimates.

Treating land cover change as a continuous covariate allows us to estimate the diffusive effect of exposure to arthroconidia arising from soil disruption due to new development. Our approach captures the effect of distance to land cover change as well as the quantity of new development in the vicinity of VF cases. The relatively large coefficients reported in Tables 3, 4, 5, and 6 are due to the fact that the land cover change variable can theoretically take values from 0 (no new development effects) to 1 (new development effects recorded in a 30-meter pixel). However, actual pixel values after applying Gaussian smoothing to the binary rasters much smaller (i.e. the maximum pixel value for the study window was smaller than 0.23 in all semesters. For example, a given residence close to a high-development region in semester 1 of 2023 might have an associated pixel value of 0.11. In this case, the estimated intensity of VF cases at a residence where the land cover change covariate is 0.11 is increased by a factor of *e*^25.755∗0.11^ = 17 compared to places where there is no nearby development. This effect decays rapidly over space. If one were to say, shift the location described above by 5km to a location with less nearby development, the factor of increase drops to just 1.076 or a 7.6% increase in VF intensity. Thus, those who are closer to development, tend to see higher risk of VF but the risk diminishes rapidly over space.

### 3.4. Relative and Predicted Intensities

An advantage of using spatial point pattern techniques is that we obtain a prediction of the VF intensity in all pixels from which we obtained quadrature points to fit the inhomogeneous Poisson process model. Thus, we obtain a full predicted intensity surface to estimate VF incidence in areas where there are no observed cases for a given semester based on the covariates of interest. In Figure 8 we display the predicted intensities for models M4 (the quadratic, full mode) and M1 (the offset-only model) for semester 2 of 2016, 2018, and 2020 to give insight into the temporal evolution of the predicted intensities. Due to the strong effect of the land cover types on the predictions at each spatial location, we smoothed the resulting predicted intensity of model M4 using a Gaussian kernel with a bandwidth of 1 km to highlight the general trend in the predicted intensity. Model M4 tends to predicted sharper peaks in intensities throughout the study window than model M1. This, along with the better fit of model M4 in terms of AIC suggests that the covariates provide greater insight into the intensity of VF cases for Medicaid patients beyond the general trend of cases that follows the Medicaid population density. Furthermore, predicted intensities also provide the basis for spatial point pattern model validation (see [7], Chapter 11).

Relative intensity^23^ estimates are a simple way to show congruence between fitted spatial point process models and a smooth, nonparametric estimate of intensity of VF cases for a given semester. Let *X*_*i,j*_ denote the unmarked spatial point process of VF cases (i.e. the superposition of all four types of VF cases, one for each age-sex category) in year *i*, semester *j* and let *λ*_*i,j*_ (*u*) denote the true intensity of *X*_*i,j*_ at *u* ∈ *W* . Note that *X*_*i,j*_ would be the unobserved process generating VF cases among the Medicaid population and the process for which we are interested in learning the intensity function. The relative intensity of *X*_*i,j*_ and a fitted model for the intensity of *X*_*i,j*_ is given by 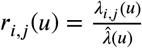 where 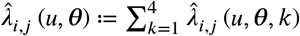 denotes the estimated intensity function of *X*_*i,j*_. We estimate the relative intensity based on an observation of *X*_*i,j*_ by

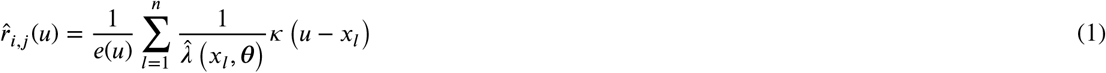

where *x*_*l*_, *l* = 1, …, *n* denotes the locations of *n* VF cases in an observation of *X*_*i,j*_, *κ* is a smoothing kernel, and *e*(*u*) is an edge correction^24^ of the estimate to deal with the bias arising from fact that we only observe the pattern inside the window *W* of finite area^25^ [8]. The relative intensity is simply a weighted estimate of the intensity of VF cases with the weight at each observed VF case equal to the inverse of the model-predicted intensity at that location (in our case we use predicted intensities of the parametric models M1 through M4). Despite performing better than the other models in most semesters, model M4 produced small predicted intensities at certain observed VF cases creating large outliers in the weights used for computing (1). We limit the visual effect of outliers to emphasize the general trend in model performance over the study window as shown in Figure 7. To achieve this, when computing the weights in (1), we replaced^26^ predicted intensities less than 0.001, with the smallest predicted intensity greater than or equal to 0.001. Using an adaptive-bandwidth kernel^27^ allows us to tailor our relative intensity estimate to protect privacy while highlighting regions within the study window where model M4 fails to explain the observed VF intensity in terms of the covariates^28^ . Figure 7 shows the estimated relative intensity for model M4 in semester 1 of 2023 which is the latest semester where model M4 performed better than other models in terms of AIC. Relative intensity plots for all years/semesters and models are available in the supplementary material.

**Figure 7:**
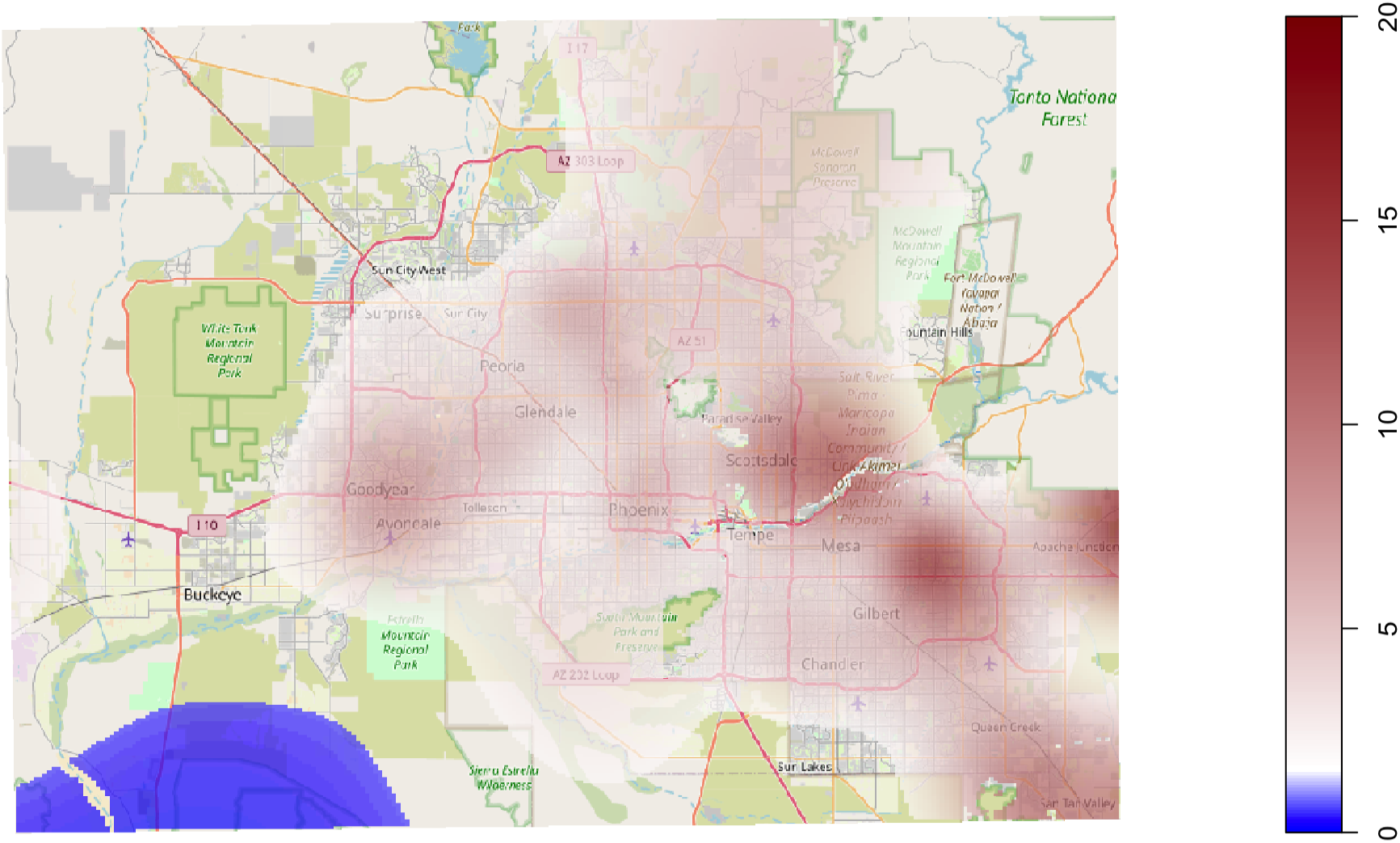
Estimated relative intensity for M4 in 2023, semester 1. Red indicates areas where the model underestimates the intensity of VF cases with darker red showing more severe underestimation, while blue indicates areas where the intensity is overestimated. Relative intensities that were NA, below 2 and above 0.5 are not shown in color. To highlight general trends in model performance, pixels with predicted intensity values (which are used to calculate the weights in (1)) less than 0.001 are replaced with the minimum predicted intensity greater than or equal to 0.001. **Source:** AHCCCS, 2023. CHiR created patient record files from Arizona Medicaid (AHCCCS) data that were used for this analysis.

**Figure 8:**
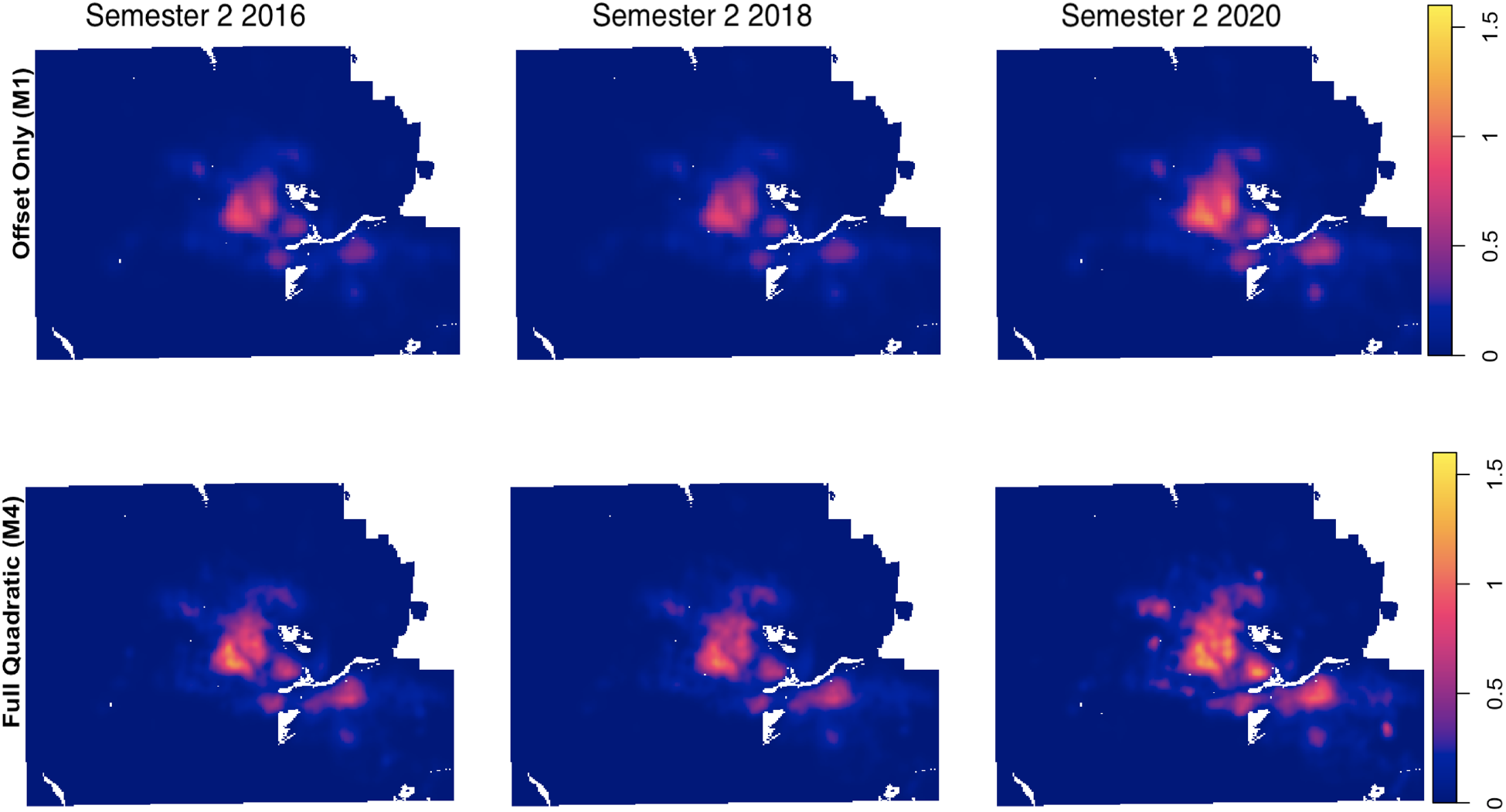
Predicted intensities for three separate years in which model M4 outperforms the other models in terms of AIC (see Table 2). Predicted intensities for model M4 are smoothed with a Gaussian kernel using a bandwidth of 1 km to highlight general patterns. **Source:** AHCCCS, 2023. CHiR created patient record files from Arizona Medicaid (AHCCCS) data that were used for this analysis.

Clearly, in the first semester of 2023, model M4 tends to perform worse in the southeastern part of the Phoenix Metro as can be seen in Figure 7. This pattern of poorer model fit in the southeast was present for many, but not all semesters. North Phoenix tended to be another area where model M4 underestimated the intensity as well. Relative intensities for semesters after 2015 tended to have pockets are more extreme over estimation, a pattern that evidently holds for all models (see the supplementary material). No other clear trends emerged in the relative intensity plots for all models and semesters. Overall, Figure 7 suggests that the model fit can still be improved systematically. We suspect that the large underestimation in the east valley shown in 7 is due to the large spatial volatility in predicted intensity in relatively small regions likely due to the nature of the strong estimated effects of land cover category on VF incidence, a pattern which holds for all semesters. This is not evident in 8 since these predicted intensities have been smoothed to highlight general trends in predicted intensity.

## 4. Discussion and Conclusion

In Maricopa county where VF is endemic, exposure to *Coccidioides* arthocronida can occur in a variety of places, yet our results suggest that for Medicaid enrollees in Arizona, local environmental factors around one’s residence do influence risk of VF. Indeed, we have shown that such factors that are known to influence VF incidence based on spatially aggregated data can inform estimates of VF risk at smaller scales. This allows a more precise understanding how local environmental factors like soil properties, NDVI, land cover type and land cover changes affect the risk of VF based on residential location. Our analysis reveals nonlinearities and confirms lagged effects in the relationship between NDVI and observed VF cases at a local spatial scale. Lastly, our approach suggests that land cover change, when measured at small spatial scales, has an effect on VF incidence, a result that differs from that of other recent work (see e.g. [27] for an example where similar land cover change data are used and [10] for a specific example regarding employees near a construction/new development site).

While our approach provides evidence at the spatial point level for environmental covariate effects on VF incidence, even our best model substantially under/overestimates VF case intensity in portions of the study region (see Figure 7) and especially in areas at the boundary of the more densely populated region of the study area. There are multiple possible reasons for this, among which the most likely is that we are using only estimates of the spatially varying Medicaid population. Indeed, some of the census block groups used for the estimates have relatively large margins of error.

Our results demonstrate that Geocoded residential addresses of VF patients can be useful in estimating VF risk at the residential level. However, more sophisticated spatial point process methods could help identify localized outbreaks. For example, a model that incorporates unobserved spatial ‘random effects’ could formally identify the regions in the study area where the covariates fail to effectively capture the spatial dynamics of VF intensity^29^ .

Our exploratory analysis in this paper has highlighted the utility of spatial point pattern analysis techniques in understanding VF incidence; however, since our data is temporally aggregated to semesters, this poses similar challenges to the spatial aggregation of VF data. If the VF cases were aggregated at, say, monthly intervals, we would observe different spatial point patterns which may reveal effect sizes that differ from those estimated at the semester-based temporal scale. Temporal aggregation of covariate data also plays a critical role in analyzing VF case intensity. It has been shown, for example, that one-month lagged precipitation is useful for predicting VF incidence at the county level [22]. Indeed, covariates that may be used to identify short-term spatially localized effects that can influence VF incidence (e.g. weather events) are not included here due to the aggregation of the case data. To address this, higher frequency environmental covariate data could be analyzed in a spatiotemporal point pattern model for VF cases that are identified by location as well as date of diagnosis or first symptoms.

## Data Availability

The empirical patient data are restricted by a data-sharing agreement with the Arizona Health Care Cost Containment System and cannot be shared. We have created a simulated patient dataset that is available online as supplementary information to this study. Environmental datasets used in the study are available at https://github.com/jginos/VF-SPP-Analysis.

https://github.com/jginos/VF-SPP-Analysis

## Acknowledgements

This work was supported under the National Institutes of Health, United States grant DMS-1615879.

This study was made possible by an ongoing partnership between Arizona State University’s Center for Health Information Research (CHiR) and the Arizona Healthcare Cost Containment System (AHCCCS). Special thanks to CHiR for providing valuable technical support to the study.

## A. Appendix

In all of the tables below, we use *,**,and *** to denote the 95%, 99%, and 99 9% significance level for the Wald for model coefficients (see [7] page 374).

## Footnotes

1 All code, covariate data and simulated VF data are available at https://github.com/jginos/VF-SPP-Analysis.

2 We gratefully acknowledge Arizona State University’s Center for Health Information Research (CHiR) and the Arizona Healthcare Cost Containment System (AHCCCS) for access to this data.

3 See chapter 9 of [7] and section 2.3.

4 Hereafter, we refer to this data as Arizona Medicaid.

5 The semesterly time series of VF cases for the Medicaid population in Maricopa County had a 79% correlation with the series of all VF cases for Maricopa County as reported by the Arizona Department of Health Services (with 2023 data being considered provisional at the time of the analysis; see [4] for annual reports on Valley fever Cases in Arizona Counties).

6 While the CAP LTER study region is roughly rectangular in latitude/longitude coordinates, as will be seen below, the plots of covariate rasters and the study window used for the analysis contained herein are based on a bounding box constructed from areas where the NDVI rasters were defined (i.e. not NA) inside the study region, which, after projecting (using the WGS 84/Pseudo-Mercator projection) deviated from the original rectangular shape.

7 All rasters, including those for land cover, NDVI, land cover change, and HSI are projected using the WGS84/Pseudo Mercator projection and rescaled to reflect measurements in kilometers. NDVI was projected using the cubic spline method available as an option when using the R package Terra. Land cover data are projected using the modal pixel resampling scheme while land cover change was projected using the nearest neighbor scheme.

8 Estimates regarding healthcare coverage by demographic group and at the block group level can be found ACS tables with ID B27010. This was used in conjunction with 2019 and 2022 TIGER/Line block group shapefiles provided by the U.S. Census Bureau to represent the 2010 and 2020 block group geometries [30], [31], [32]

9 This was done for all census block groups in Arizona but only the subset of block groups contained in the study window were included in the analysis (this includes fractional portions of block groups near the boundaries of the study window)

10 This fraction does not significantly impact the analysis as block groups with an estimated Medicaid enrollment population of zero not likely locations for VF cases and did not have large margins of error

11 Winter rasters include landsat imagery from December 21st the prior year through March 19th of the current year. Spring rasters include imagery from March 20th through June 20th and Fall NDVI rasters include imagery starting on June 21st.

12 Differenced NDVI covariates were obtained after conversion from a raster to a pixel image for compatibility with the R package spatstat

13 We would like to thank Robert R. Dobos for help with obtaining the HSI data for this study.

14 Using gridded Soil Survey Geographic Database (SSURGO) data and a table with HSI values containing a map unit key, the HSI raster layer was created using ArcGIS Pro 3.3.2 [16].

17 According to the NLCD class legend and description “…These areas most commonly include large-lot single-family housing units, parks, golf courses, and vegetation planted in developed settings for recreation, erosion control, or aesthetic purposes.” [25]

18 When projecting land cover change rasters to the WGS 84 coordinate system, a nearest neighbor scheme was used for sampling pixel values

19 However, it should be noted that the logistic regression approach used in [11] would yield similar results to the spatial point process approach as the number of pseudo-absences, either placed on a regular grid or in a uniformly random way over the study window, goes to infinity (see [35] for details on this).

20 While the entire grid is 250 × 250 dummy points, those points located where any covariate was labeled NA were removed from the quadrature scheme used to estimate the intensity over the window of study.

21 See documentation of the ‘ppm’ function in spatstat for details.

22 The pixel-wise standard deviation of the HSI raster used for analysis is 0.187 over *W* . The pixel-wise mean HSI is about 0.4 over *W* .

23 See [7], chapter 11, section 11.2.1 for a description of this approach.

24 We used the spatstat default adaptive bandwidth edge correction given by 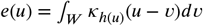 for *u* ∈ *W*. Here *h*(*u*) denotes a spatially varying bandwidth; see section 2.3 in [14] for more details on edge correction factors in adaptive bandwidth kernel density estimation.

25 see chapter 6, section 6.5.1 and chapter 11, section 11.2.1 in [8] for more details on edge correction factors and relative intensity estimates for model validation, respectively.

26 In semester 1 of 2023, predicted intensities from model M4 were replaced at 14 out of the 702 observed VF cases while for model M1, predicted intensities were replaced at 11 such locations.

27 See [8] and [7], especially chapter 6, for details on nonparametric intensity estimation.

28 A global bandwidth of 10 km was chosen for use as a pilot bandwidth which is passed to the spatstat function bw.abram.ppp. See also [1] and [14] for details on how this method works.

29 See [7], chapter 12 for a discussion of relevant methods (e.g. Cox processes).

## Notes

### Competing Interest Statement

The authors have declared no competing interest.

### Author Declarations

The Institutional Review Board of Arizona State University gave ethical approval for this work.

### Summary of Updates

We Have changed the nature of the land cover change covariate which has not affected overall model results significantly but the interpretation is different and the model results (i.e. effect size and statistical significance) for this particular covariate are new. We have also added predicted intensity plots, accompanying discussion, and updated various small errors and typos.

