## Supplementary file for "Spatial Point Pattern Analysis of Environmental Effects on Valley Fever Intensity in Phoenix, Arizona"

#### Relative intensities for all models and semesters

Below we show the estimated relative intensities for the Medicaid population offset only model (M1) , land cover and land change covariate model (M2), the linear model with all environmental covariates (M3), and the quadratic model with all environmental covariates (M4) in all semesters and years. Brighter colors indicate areas where the model underestimates the intensity of VF cases. Darker colors indicate areas where the intensity is overestimated. Relative intensities that were NA are blank white as are areas that have estimated relative intensities beyond twenty, the upper boundary of values for the color ribbon. To highlight general trends in model performance, pixels with predicted intensity values (which are used to calculate the weights in (1) in the main text of the paper) less than 0.001 are replaced with the minimum predicted intensity greater than or equal to 0.001. **Source:** AHCCCS, 2023. CHiR created patient record files from Arizona Medicaid (AHCCCS) data that were used for this analysis (including the plots below).

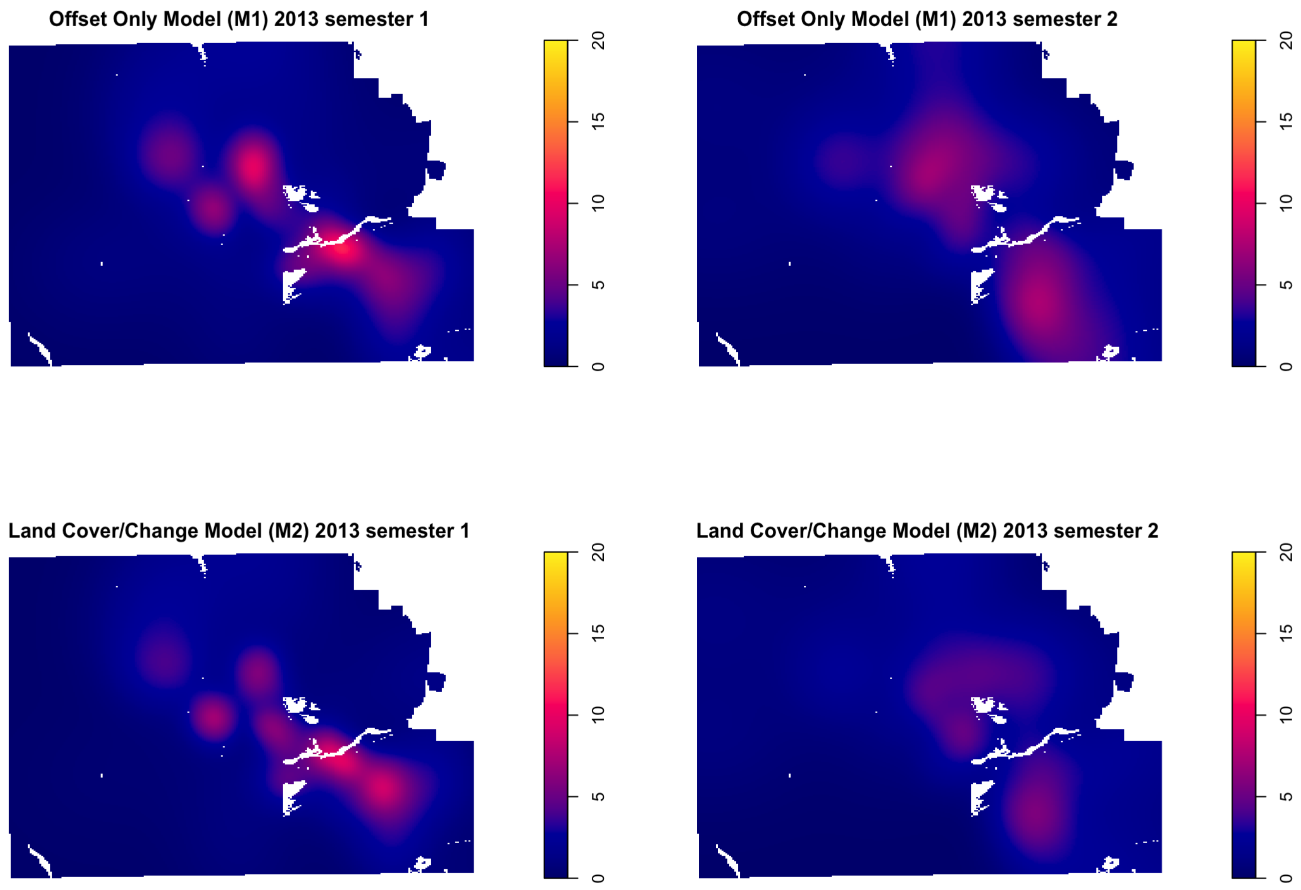
Offset Only Model (M1) 2013 semester 1
Offset Only Model (M1) 2013 semester 2
Land Cover/Change Model (M2) 2013 semester 1
Land Cover/Change Model (M2) 2013 semester 2
0
5
10
15
20

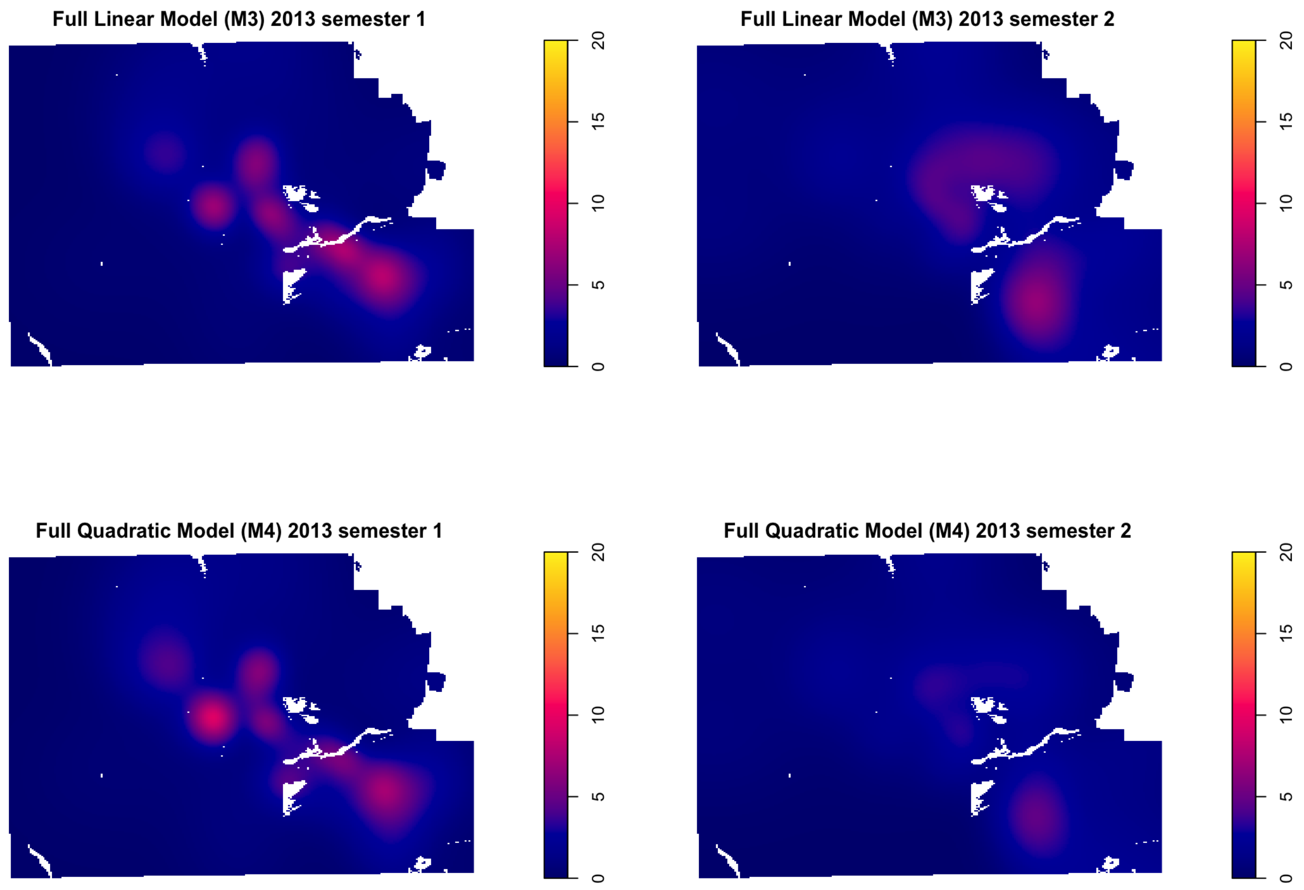
Full Linear Model (M3) 2013 semester 1
Full Linear Model (M3) 2013 semester 2
Full Quadratic Model (M4) 2013 semester 1
Full Quadratic Model (M4) 2013 semester 2
0
5
10
15
20

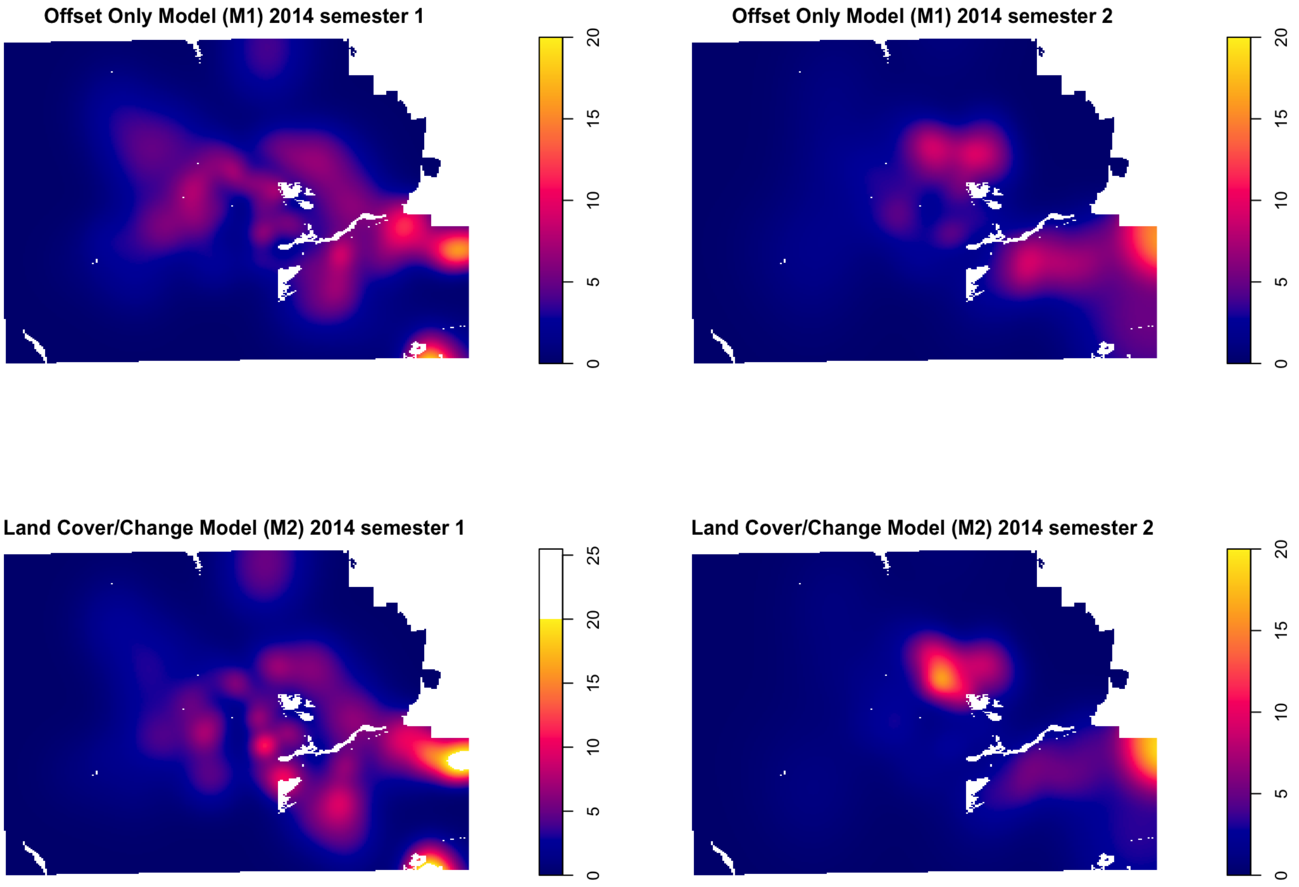

Offset Only Model (M1) 2014 semester 1
0
5
10
15
20
Offset Only Model (M1) 2014 semester 2
0
5
10
15
20
Land Cover/Change Model (M2) 2014 semester 1
0
5
10
15
20
25
Land Cover/Change Model (M2) 2014 semester 2
0
5
10
15
20

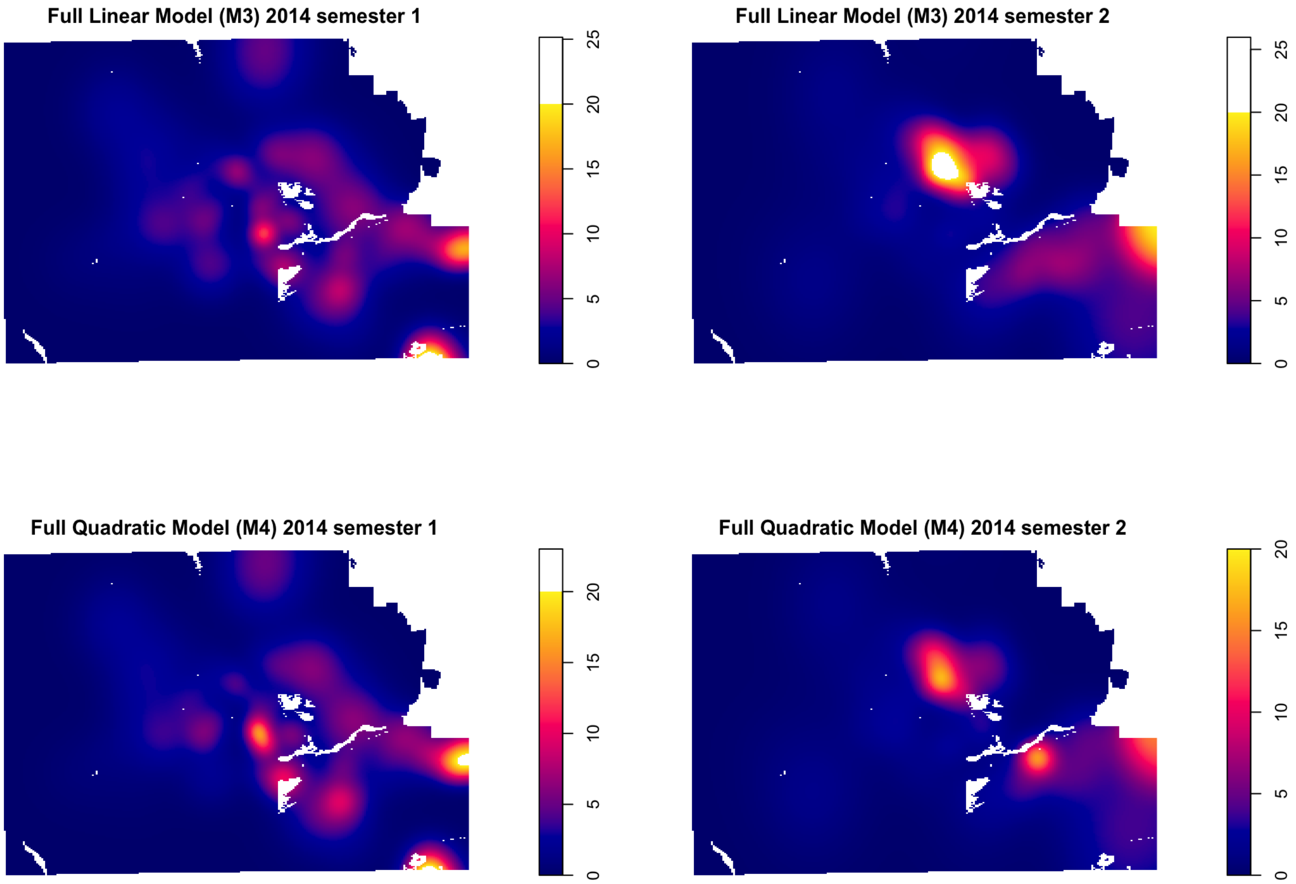
Full Linear Model (M3) 2014 semester 1
0
5
10
15
20
25
Full Linear Model (M3) 2014 semester 2
0
5
10
15
20
25
Full Quadratic Model (M4) 2014 semester 1
0
5
10
15
20
Full Quadratic Model (M4) 2014 semester 2
0
5
10
15
20

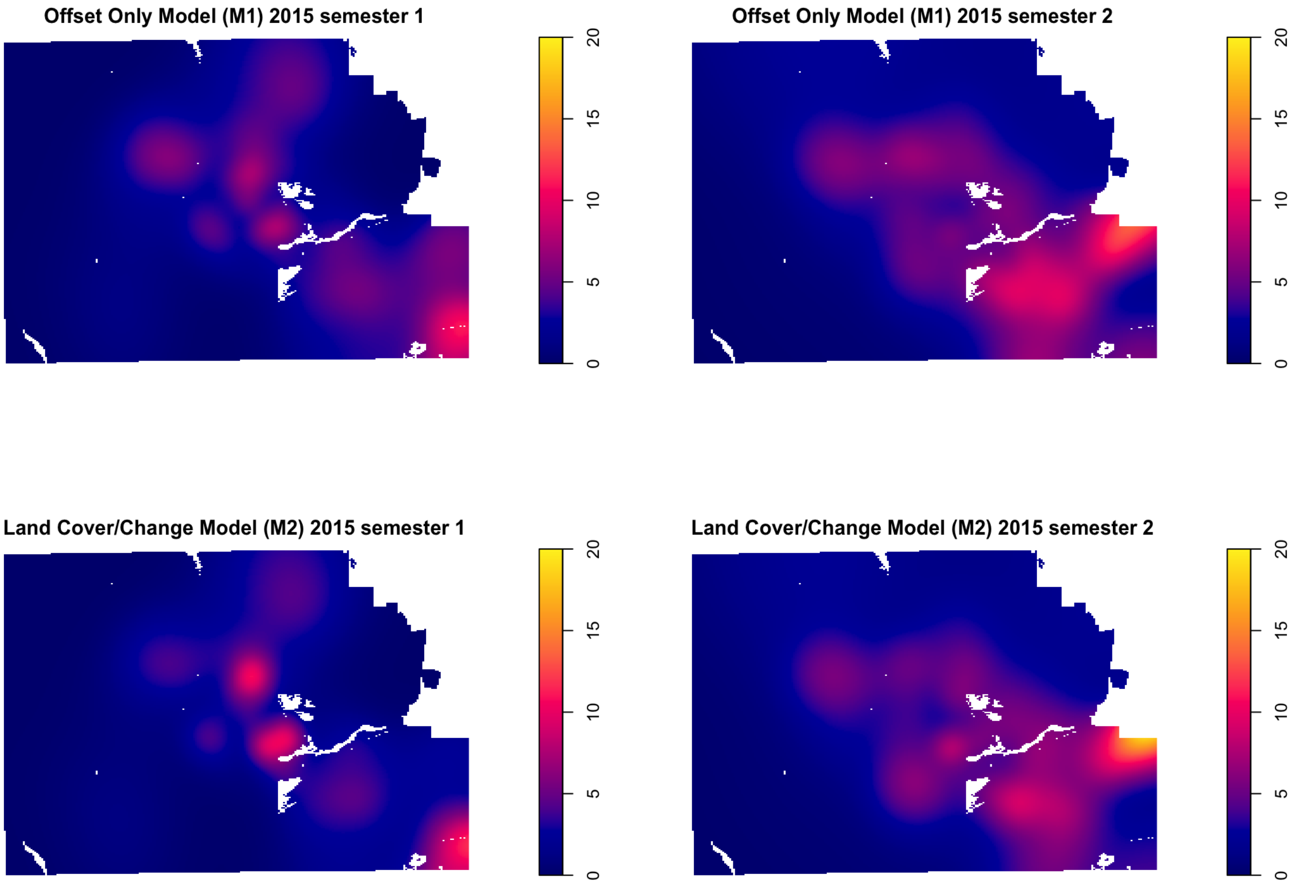

Offset Only Model (M1) 2015 semester 1
Offset Only Model (M1) 2015 semester 2
Land Cover/Change Model (M2) 2015 semester 1
Land Cover/Change Model (M2) 2015 semester 2
0
5
10
15
20

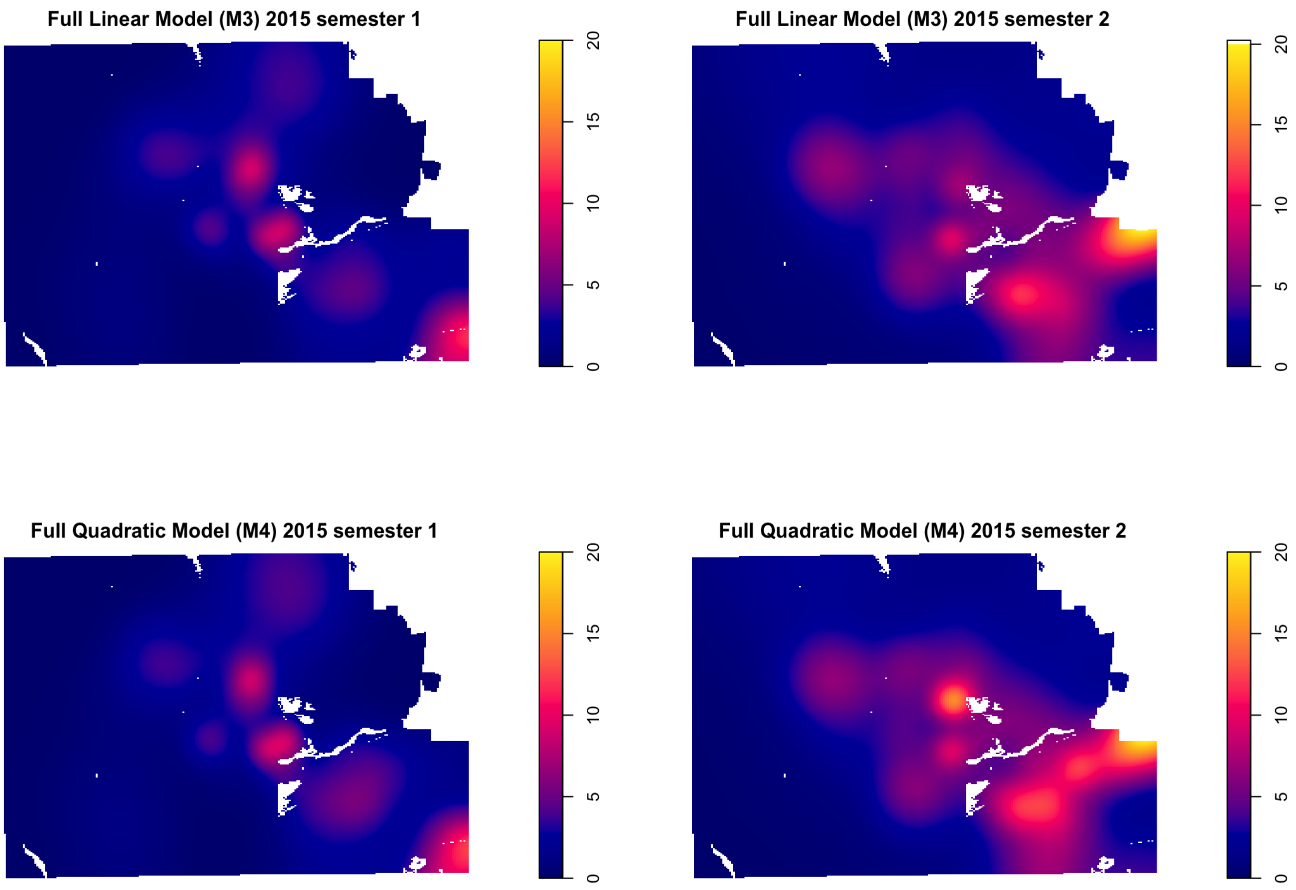
Full Linear Model (M3) 2015 semester 1
Full Linear Model (M3) 2015 semester 2
Full Quadratic Model (M4) 2015 semester 1
Full Quadratic Model (M4) 2015 semester 2
0
5
10
15
20

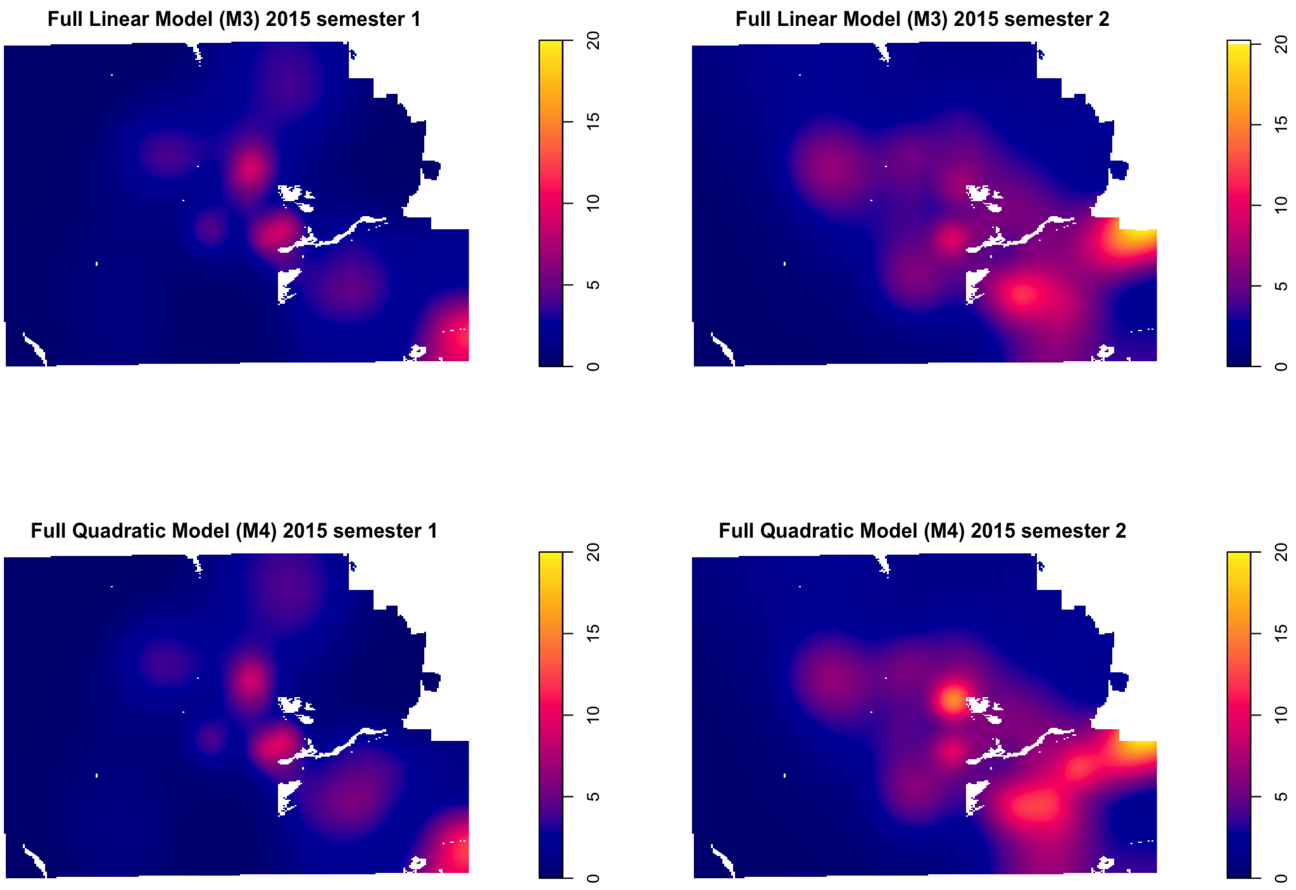
Full Linear Model (M3) 2015 semester 1
Full Linear Model (M3) 2015 semester 2
Full Quadratic Model (M4) 2015 semester 1
Full Quadratic Model (M4) 2015 semester 2
0
5
10
15
20

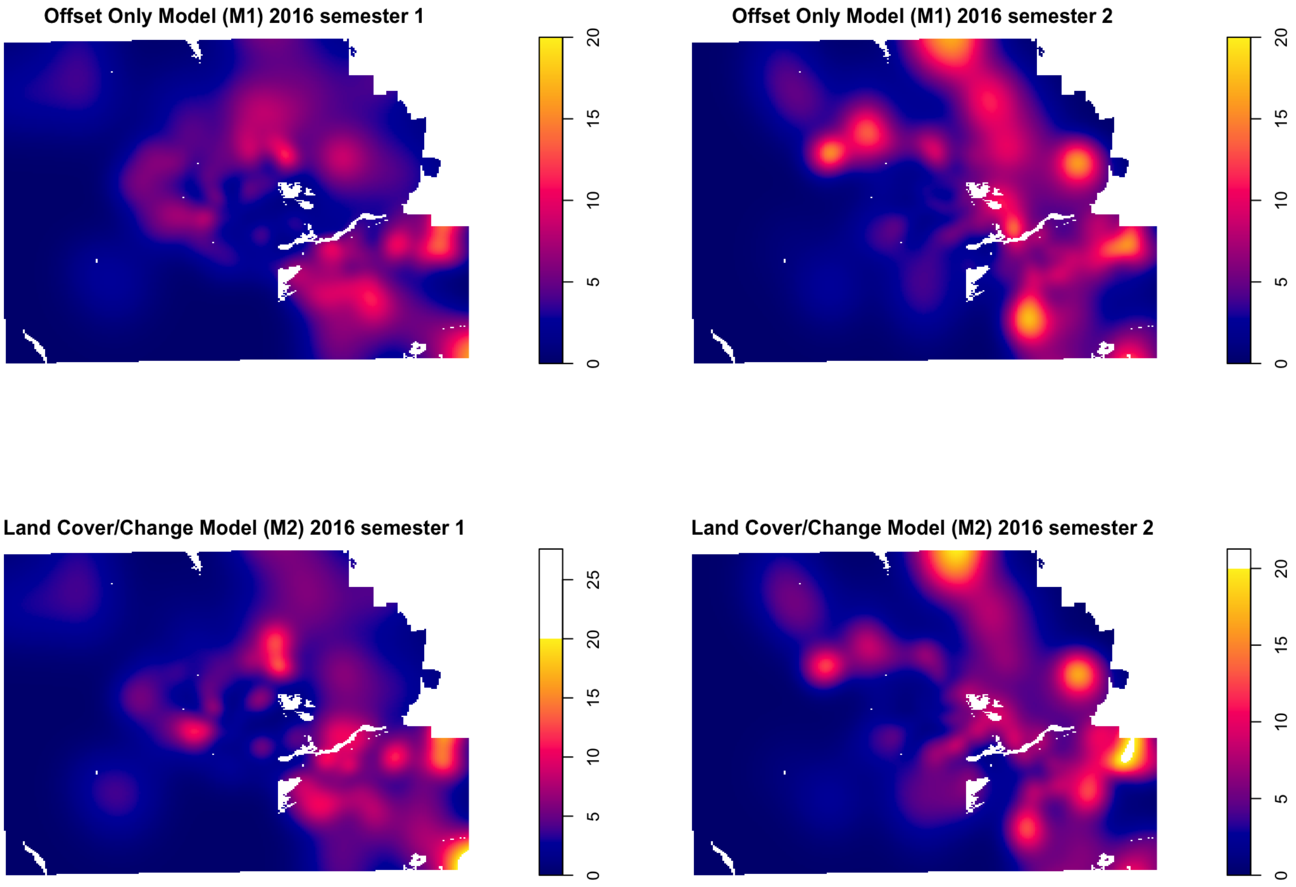
Offset Only Model (M1) 2016 semester 1
0
5
10
15
20
Offset Only Model (M1) 2016 semester 2
0
5
10
15
20
Land Cover/Change Model (M2) 2016 semester 1
0
5
10
15
20
25
Land Cover/Change Model (M2) 2016 semester 2
0
5
10
15
20

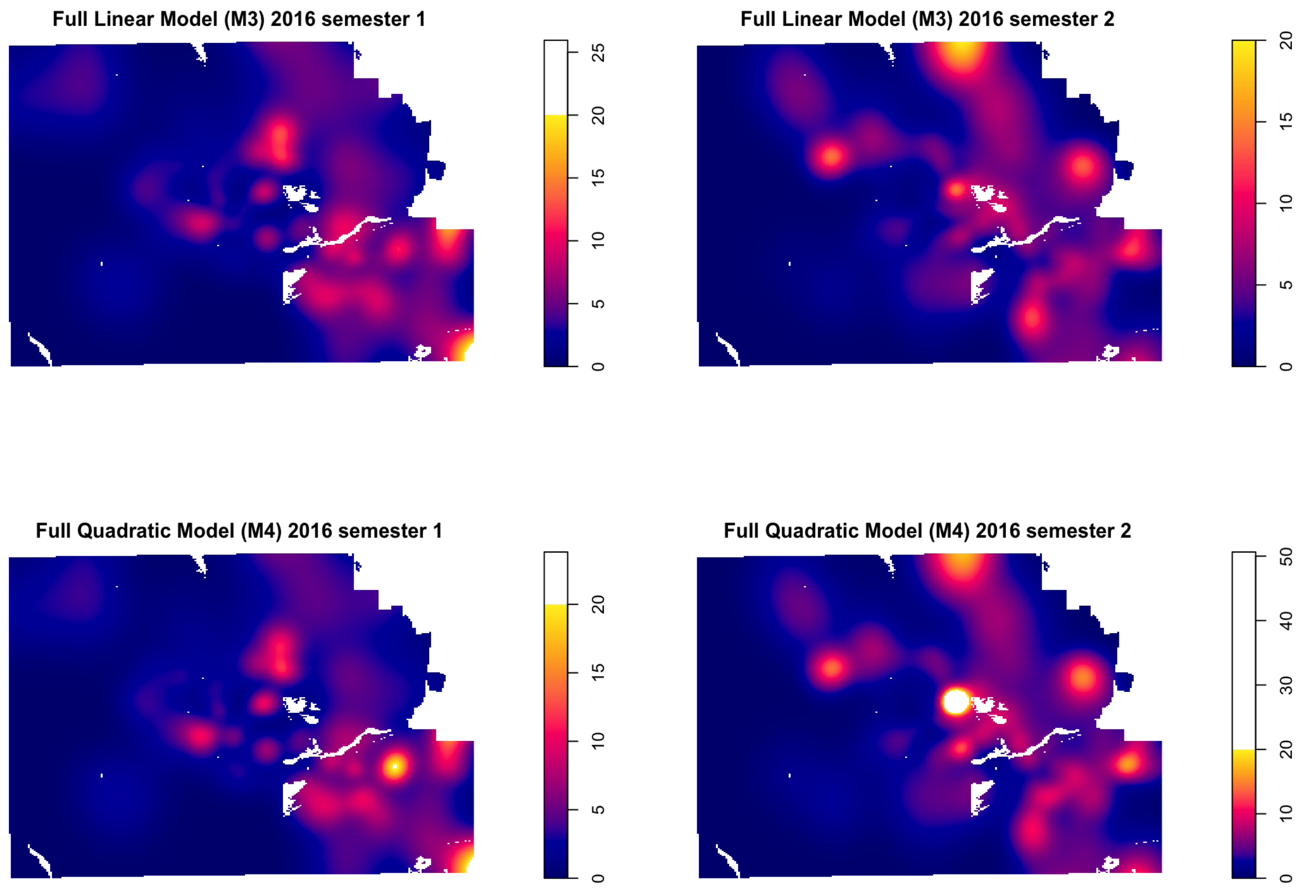
Full Linear Model (M3) 2016 semester 1
0
5
10
15
20
25
Full Linear Model (M3) 2016 semester 2
0
5
10
15
20
Full Quadratic Model (M4) 2016 semester 1
0
5
10
15
20
Full Quadratic Model (M4) 2016 semester 2
0
10
20
30
40
50

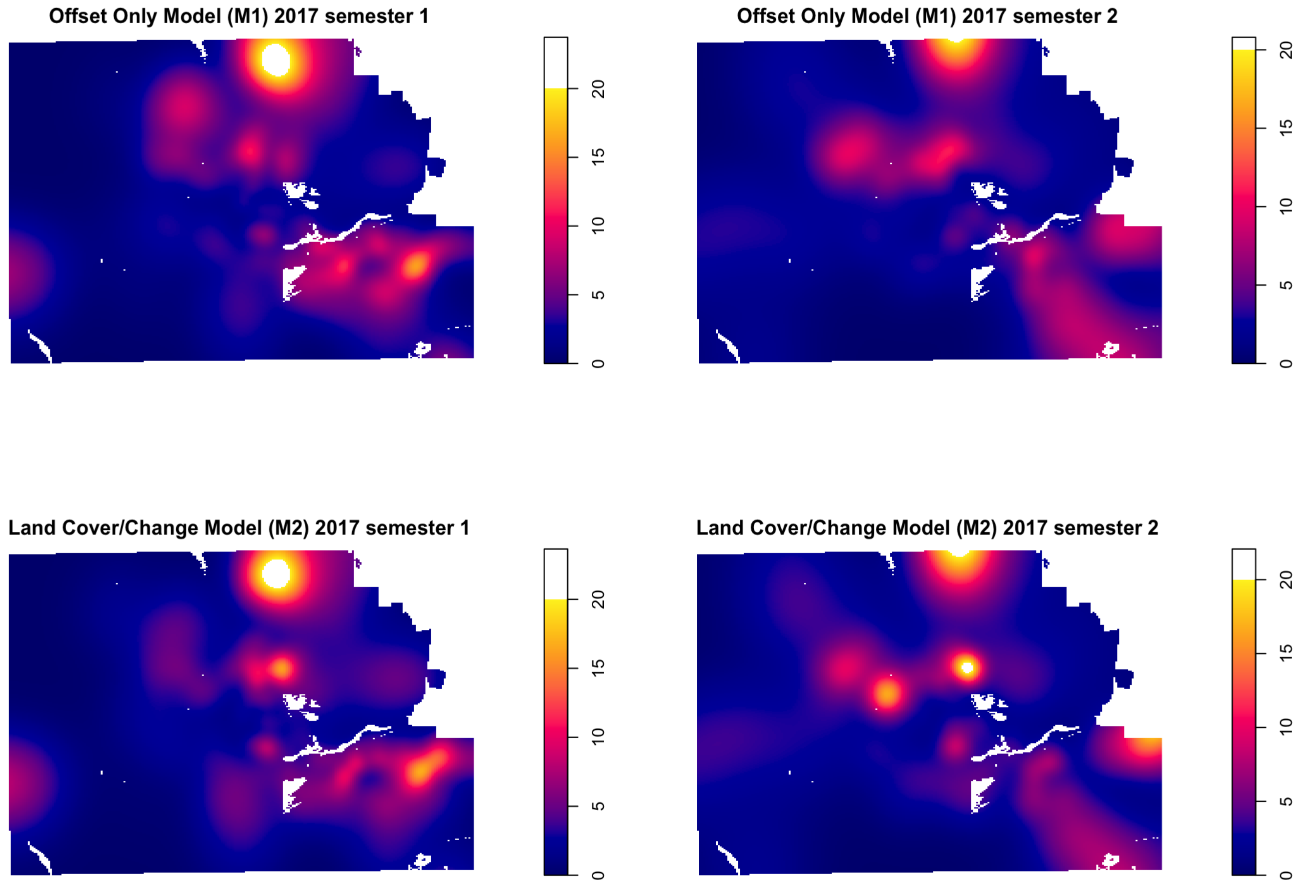

Offset Only Model (M1) 2017 semester 1
0
5
10
15
20
Offset Only Model (M1) 2017 semester 2
0
5
10
15
20
Land Cover/Change Model (M2) 2017 semester 1
0
5
10
15
20
Land Cover/Change Model (M2) 2017 semester 2
0
5
10
15
20

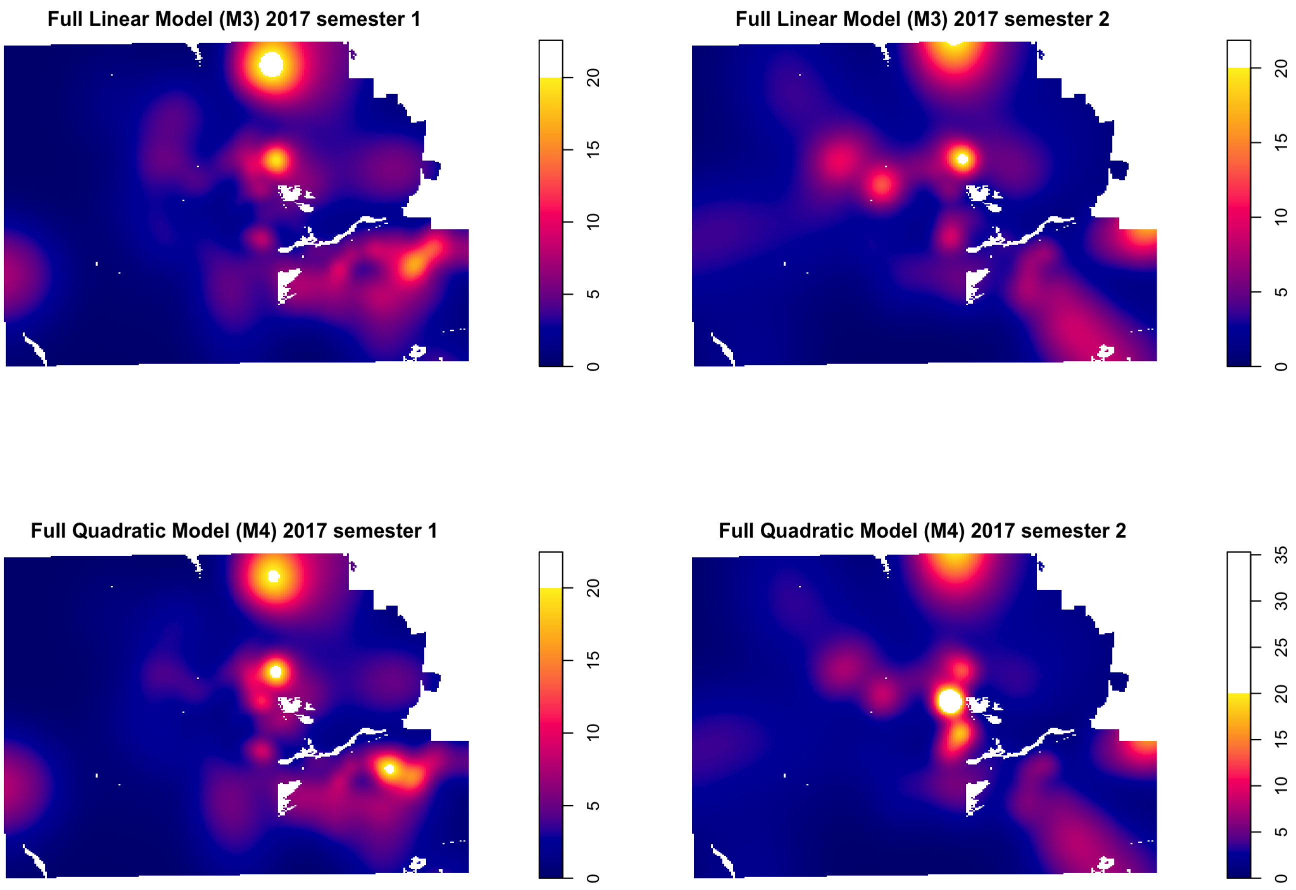
Full Linear Model (M3) 2017 semester 1
0 5 10 15 20
Full Linear Model (M3) 2017 semester 2
0 5 10 15 20
Full Quadratic Model (M4) 2017 semester 1
0 5 10 15 20
Full Quadratic Model (M4) 2017 semester 2
0 5 10 15 20 25 30 35

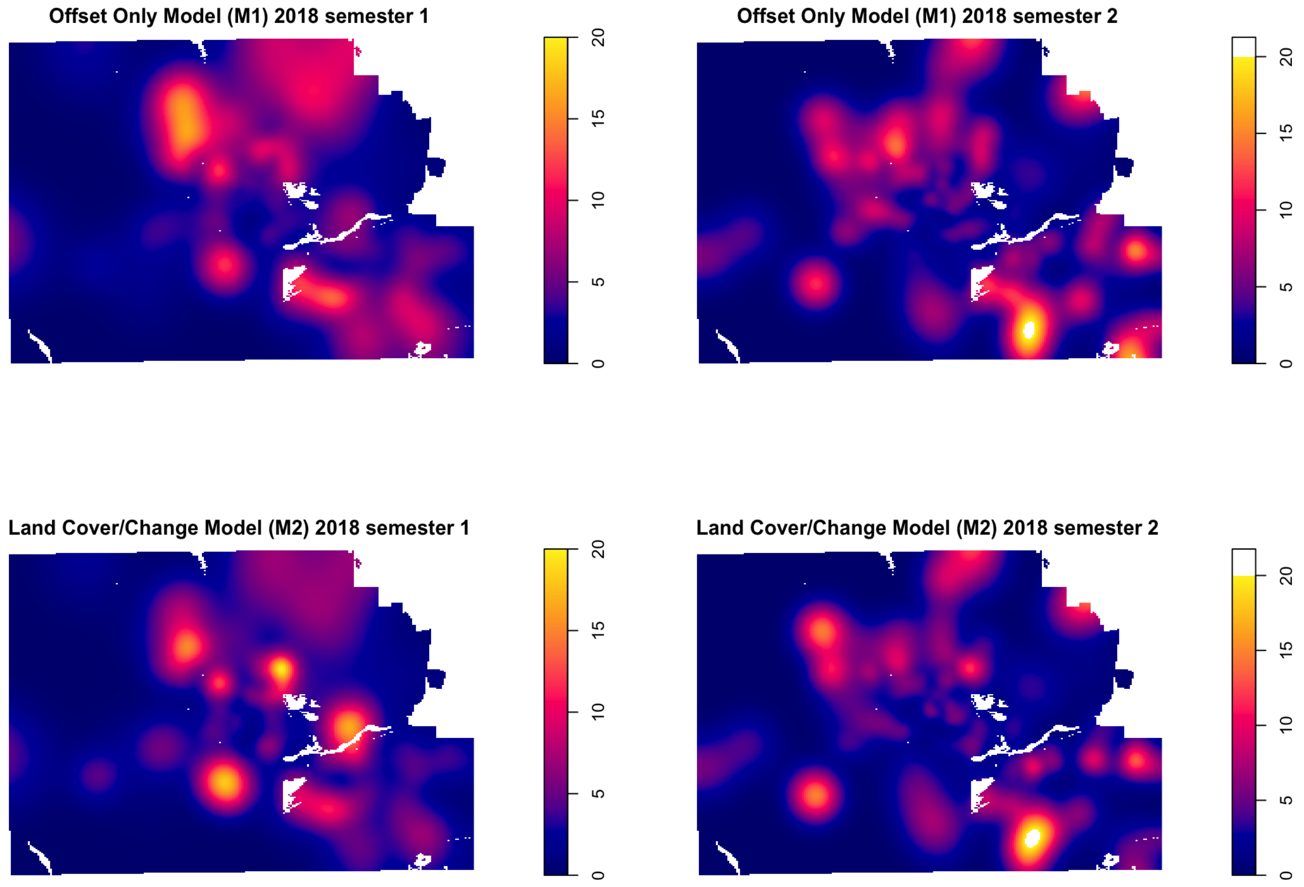

Offset Only Model (M1) 2018 semester 1
Offset Only Model (M1) 2018 semester 2
Land Cover/Change Model (M2) 2018 semester 1
Land Cover/Change Model (M2) 2018 semester 2
0
5
10
15
20

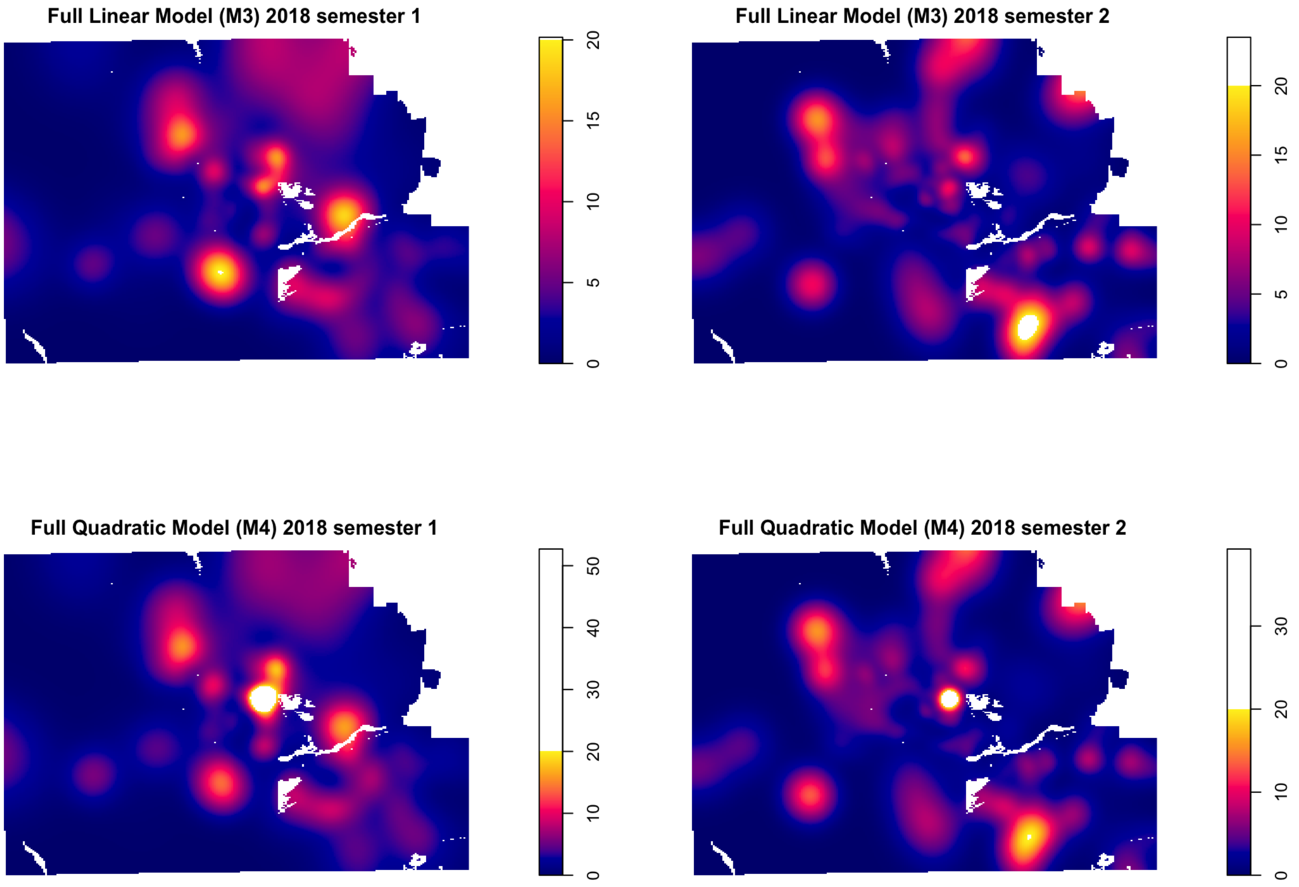
Full Linear Model (M3) 2018 semester 1
0
5
10
15
20
Full Linear Model (M3) 2018 semester 2
0
5
10
15
20
Full Quadratic Model (M4) 2018 semester 1
0
10
20
30
40
50
Full Quadratic Model (M4) 2018 semester 2
0
10
20
30

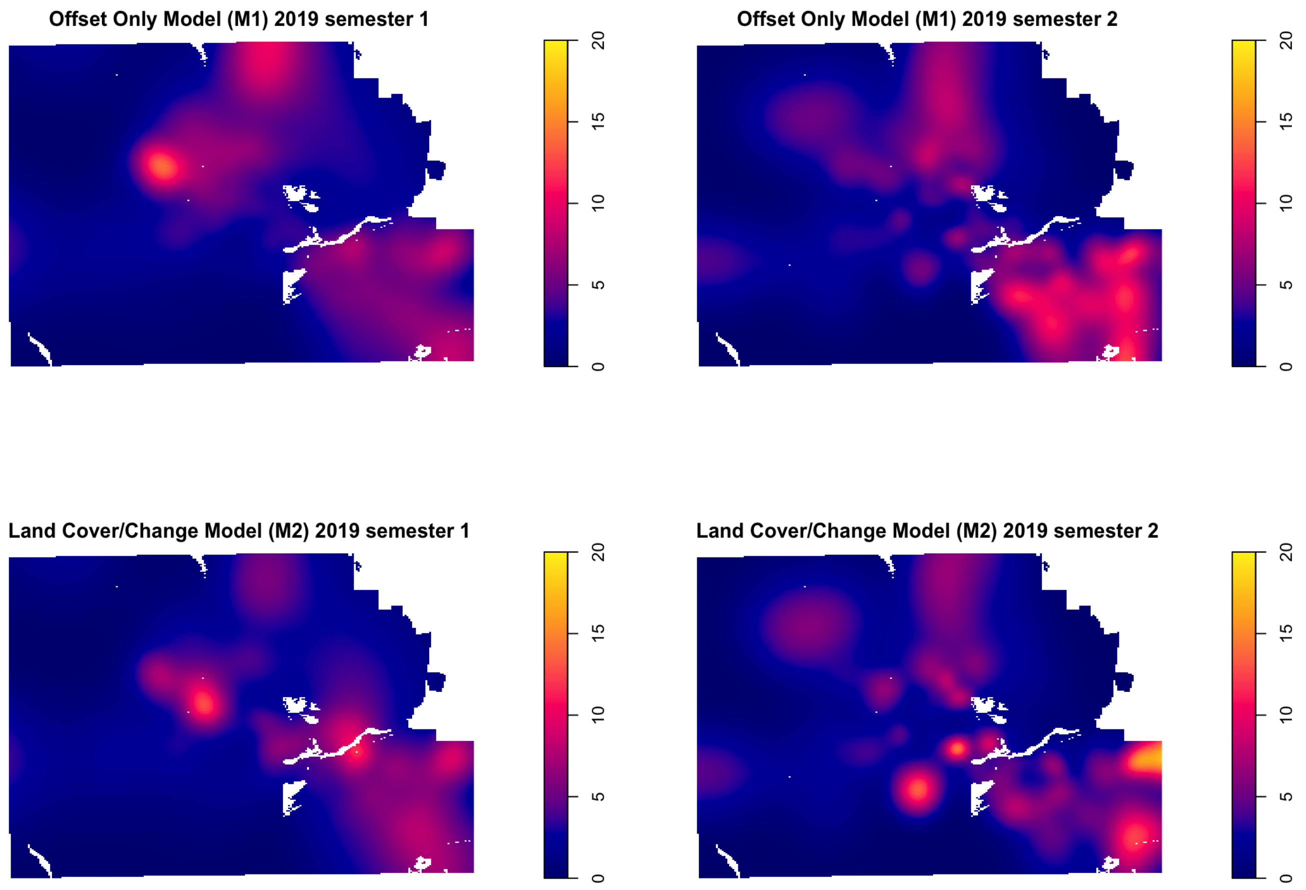

Offset Only Model (M1) 2019 semester 1
Offset Only Model (M1) 2019 semester 2
Land Cover/Change Model (M2) 2019 semester 1
Land Cover/Change Model (M2) 2019 semester 2
0
5
10
15
20

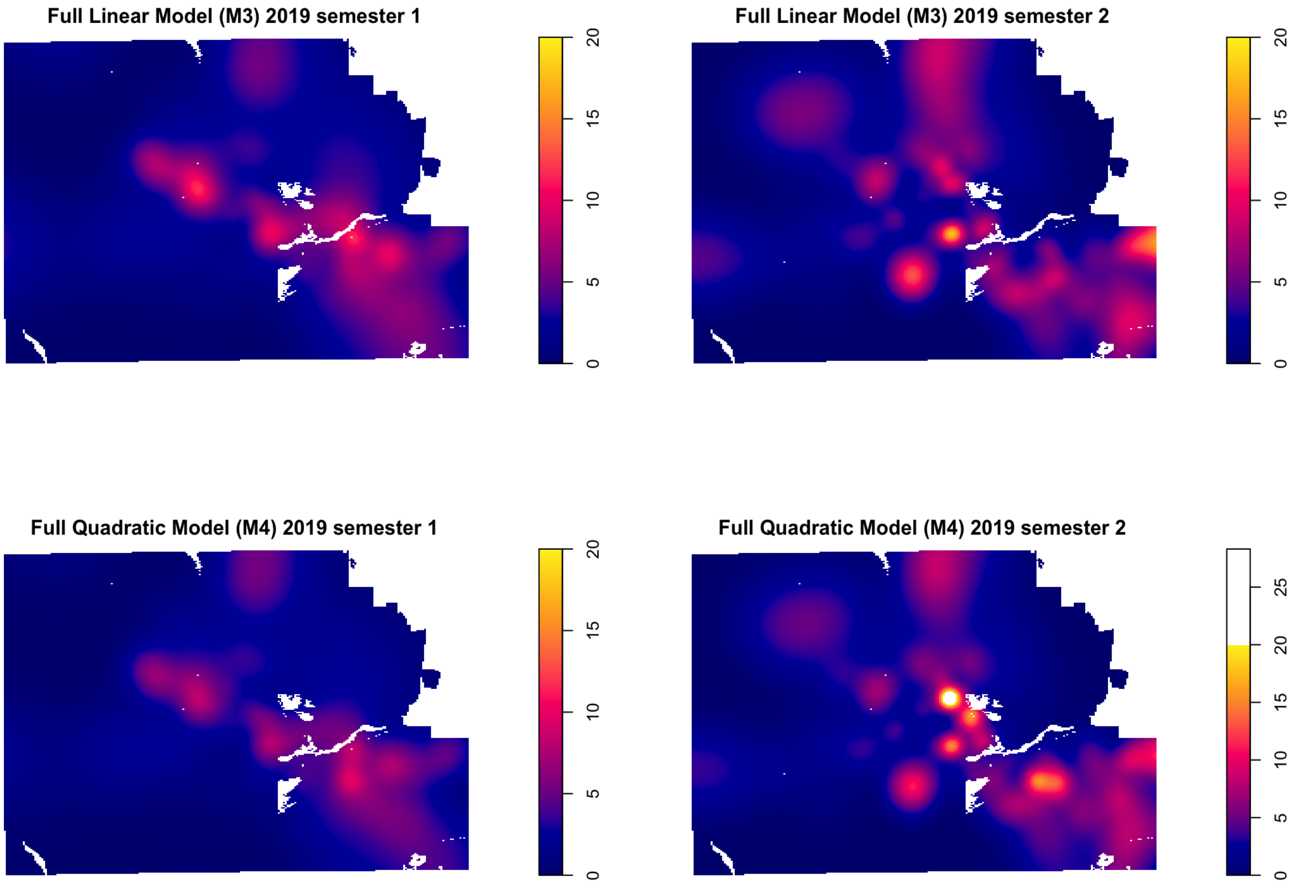
Full Linear Model (M3) 2019 semester 1
0
5
10
15
20
Full Linear Model (M3) 2019 semester 2
0
5
10
15
20
Full Quadratic Model (M4) 2019 semester 1
0
5
10
15
20
Full Quadratic Model (M4) 2019 semester 2
0
5
10
15
20
25

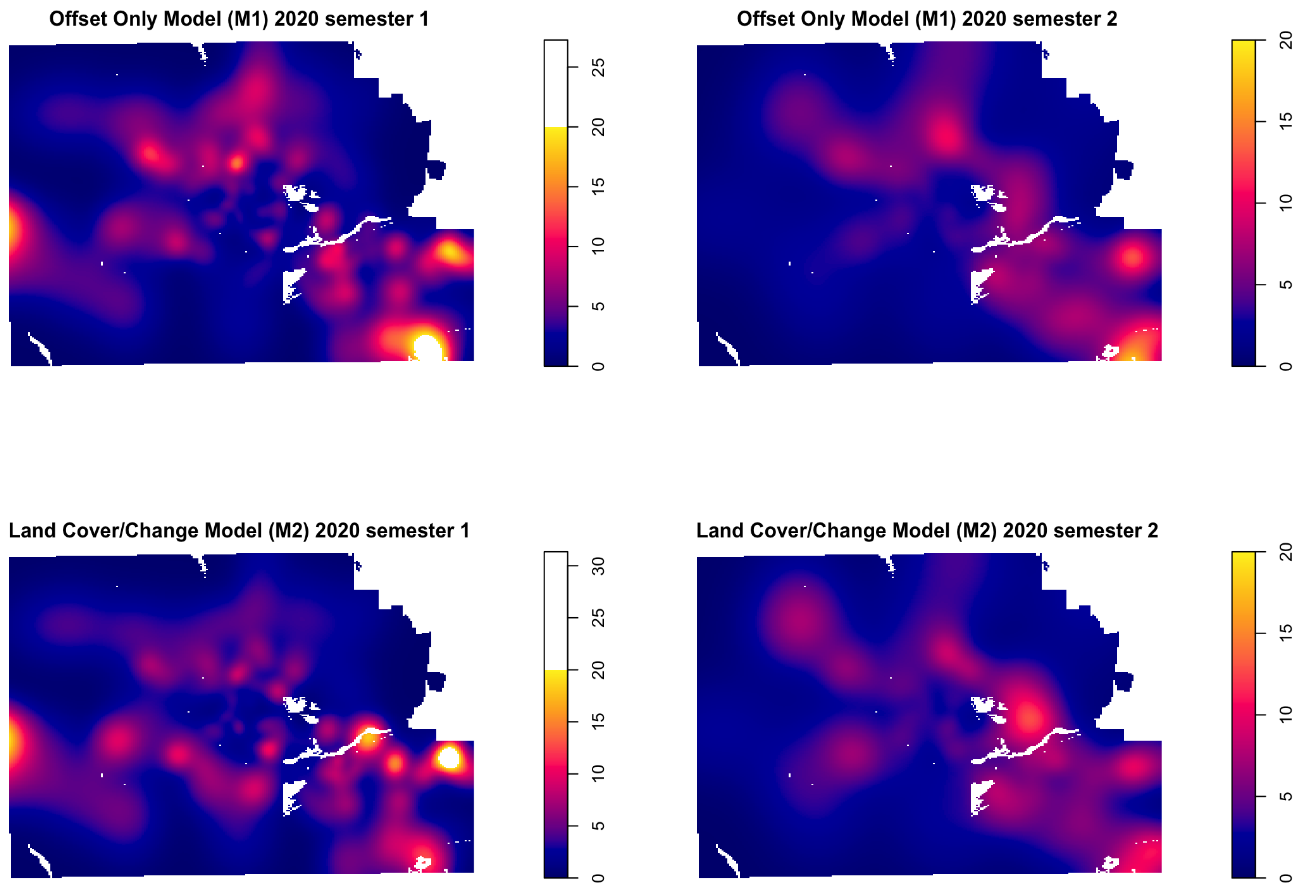
Offset Only Model (M1) 2020 semester 1
0
5
10
15
20
25
Offset Only Model (M1) 2020 semester 2
0
5
10
15
20
Land Cover/Change Model (M2) 2020 semester 1
0
5
10
15
20
25
30
Land Cover/Change Model (M2) 2020 semester 2
0
5
10
15
20

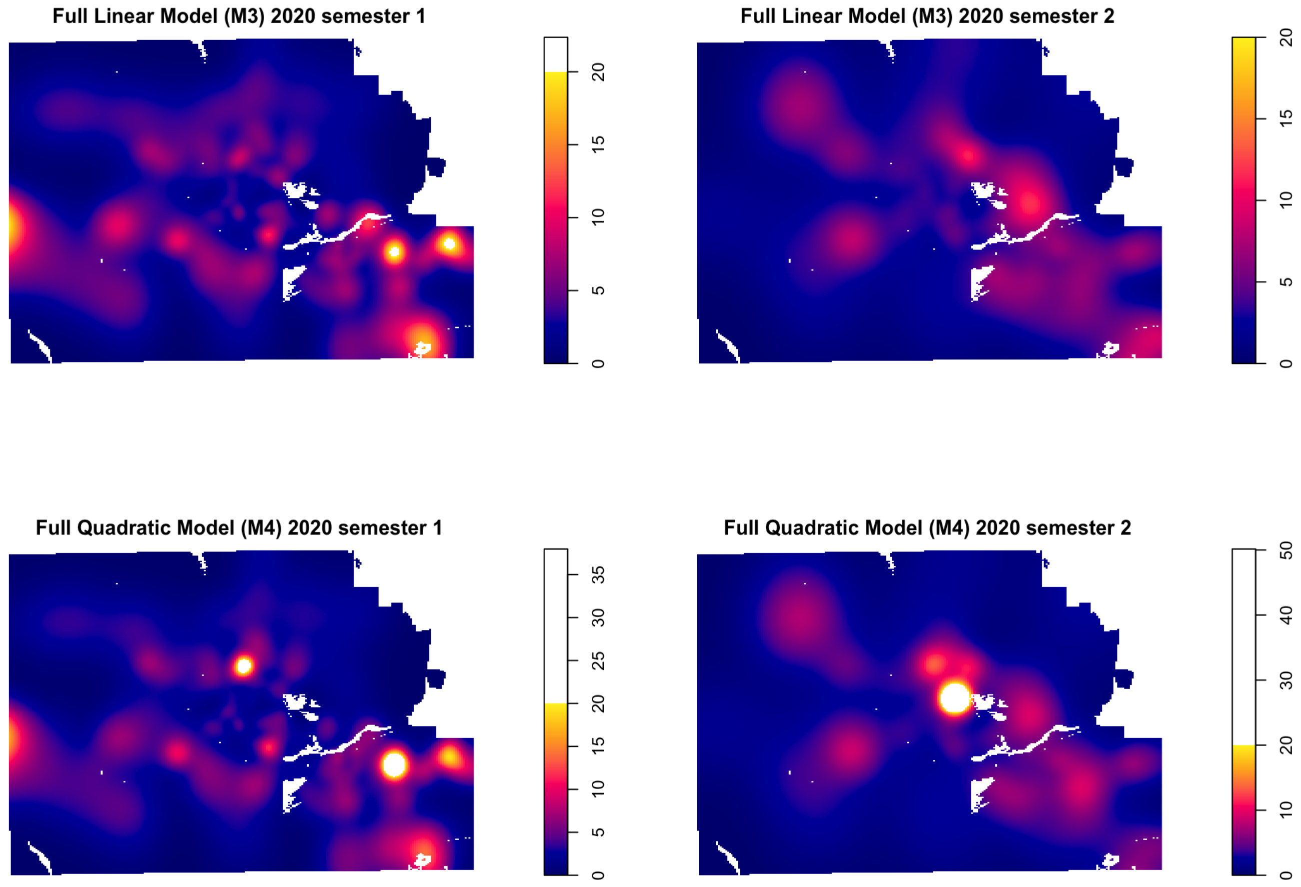
Full Linear Model (M3) 2020 semester 1
Full Linear Model (M3) 2020 semester 2
Full Quadratic Model (M4) 2020 semester 1
Full Quadratic Model (M4) 2020 semester 2

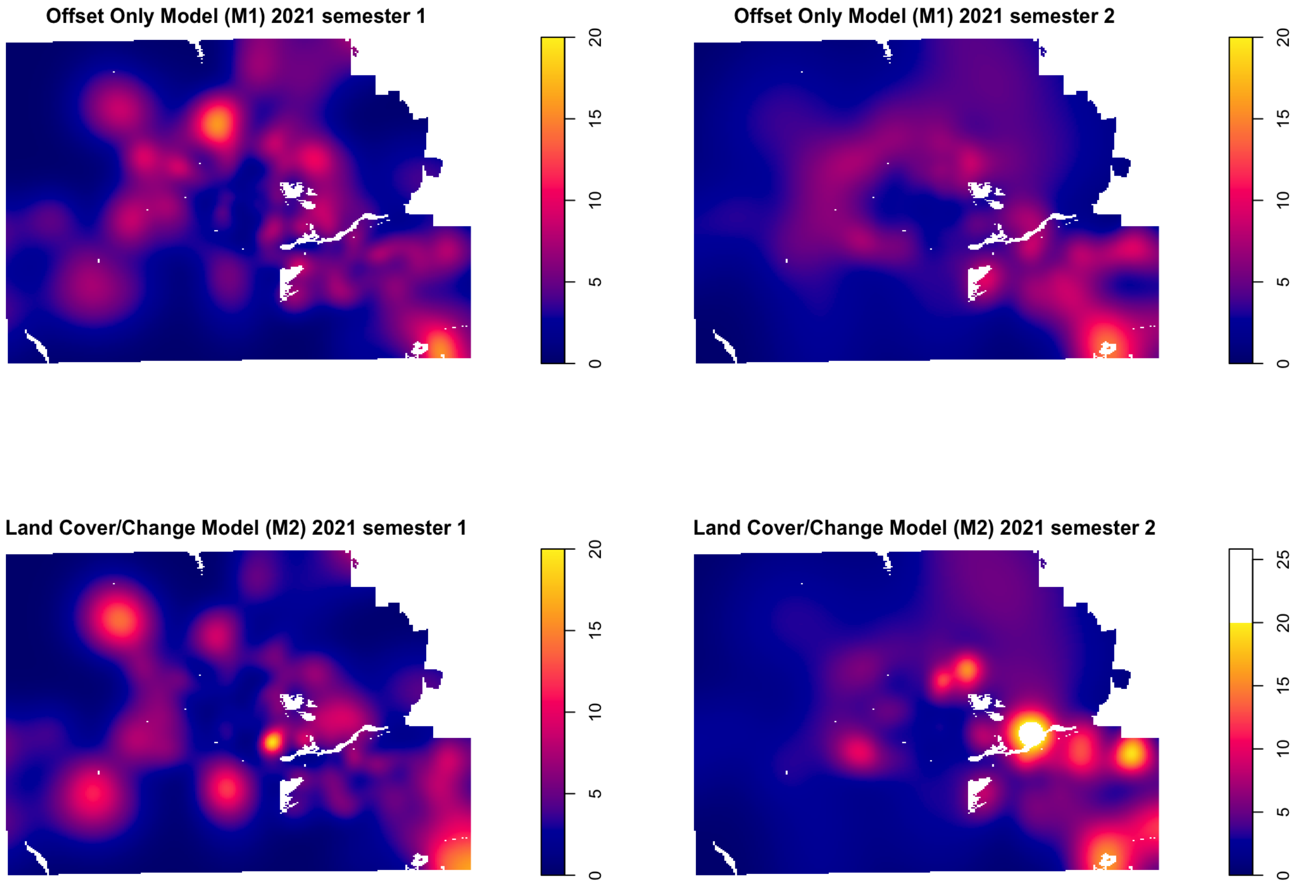
Offset Only Model (M1) 2021 semester 1
Offset Only Model (M1) 2021 semester 2
Land Cover/Change Model (M2) 2021 semester 1
Land Cover/Change Model (M2) 2021 semester 2
0
5
10
15
20
25

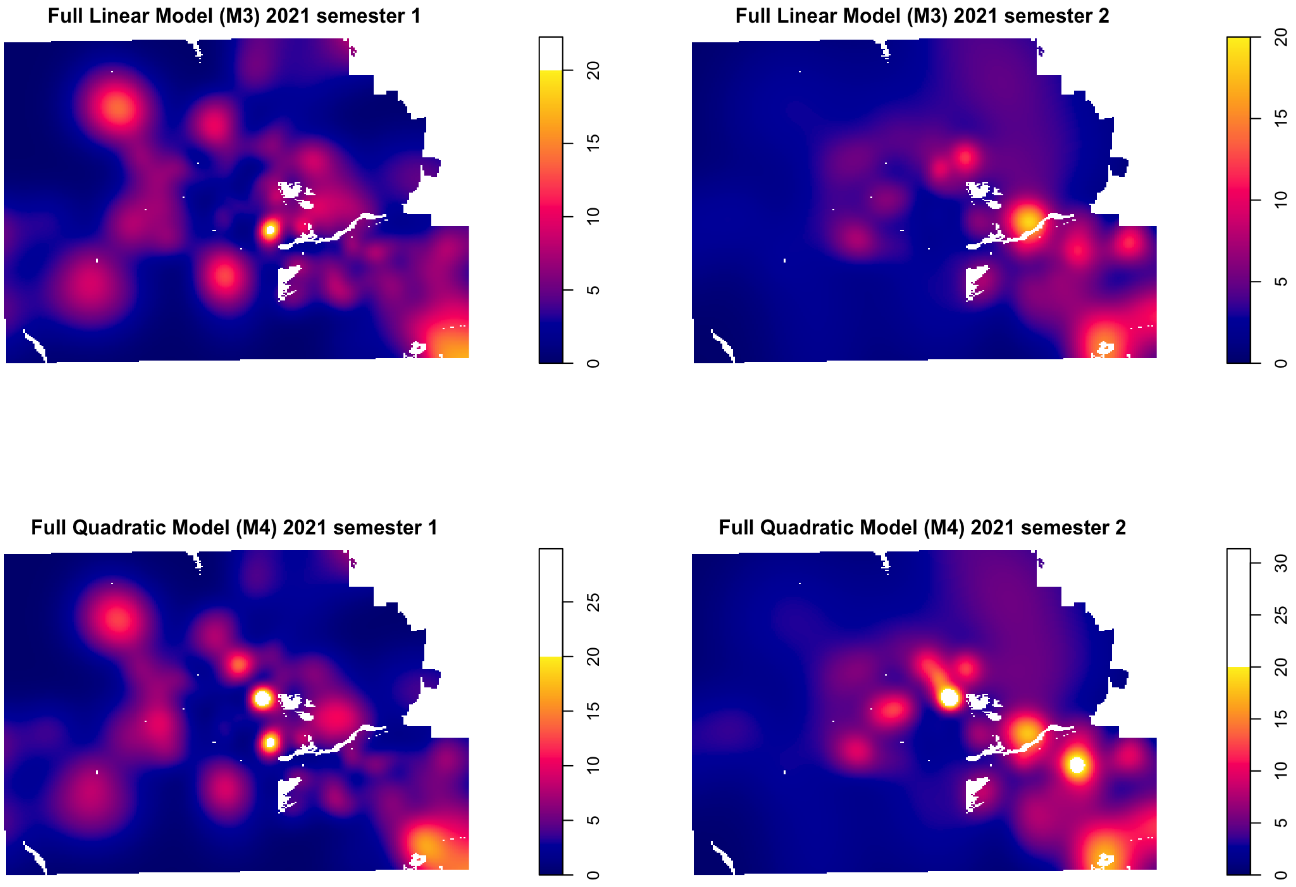
Full Linear Model (M3) 2021 semester 1
0
5
10
15
20
Full Linear Model (M3) 2021 semester 2
0
5
10
15
20
Full Quadratic Model (M4) 2021 semester 1
0
5
10
15
20
25
Full Quadratic Model (M4) 2021 semester 2
0
5
10
15
20
25
30

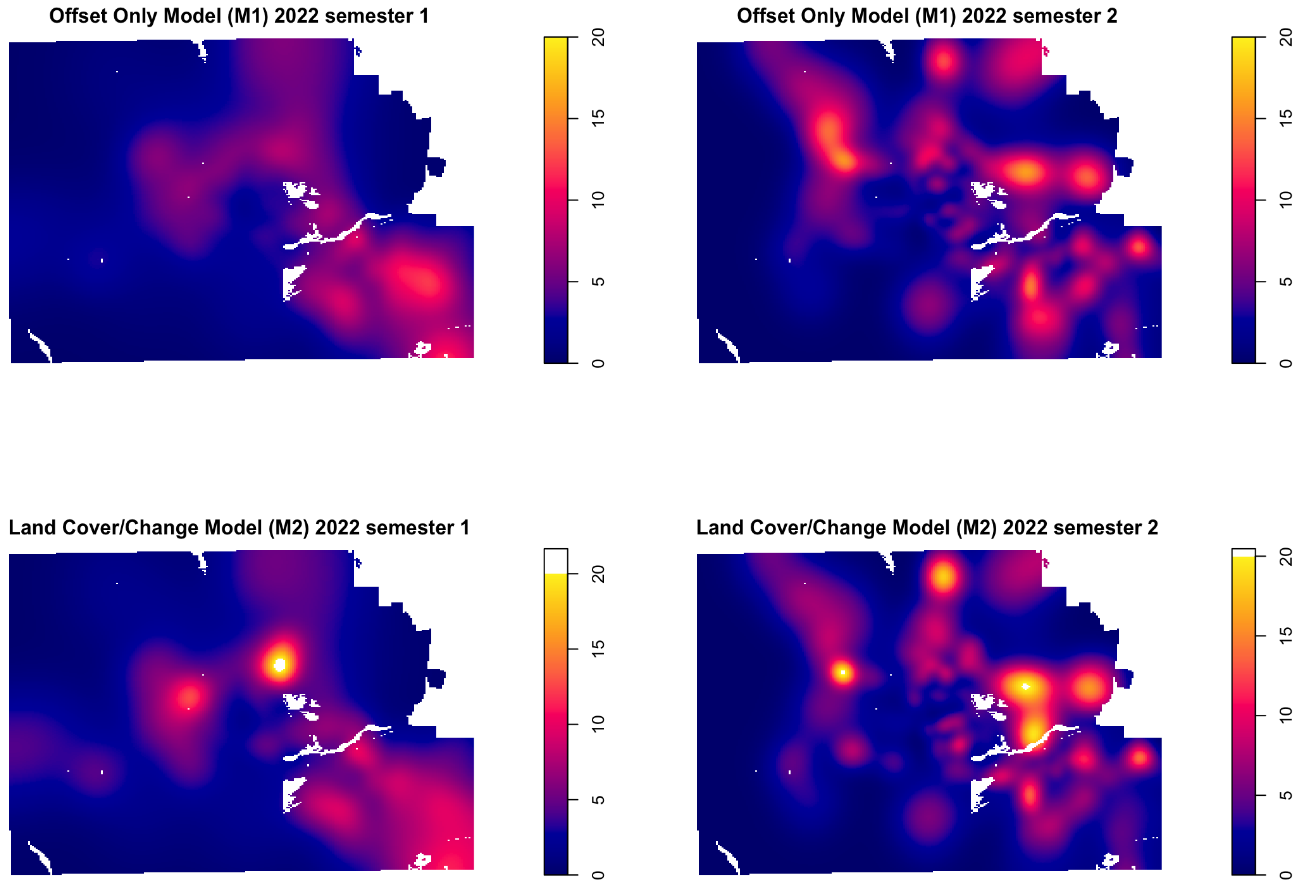

Offset Only Model (M1) 2022 semester 1
Offset Only Model (M1) 2022 semester 2
Land Cover/Change Model (M2) 2022 semester 1
Land Cover/Change Model (M2) 2022 semester 2
0
5
10
15
20

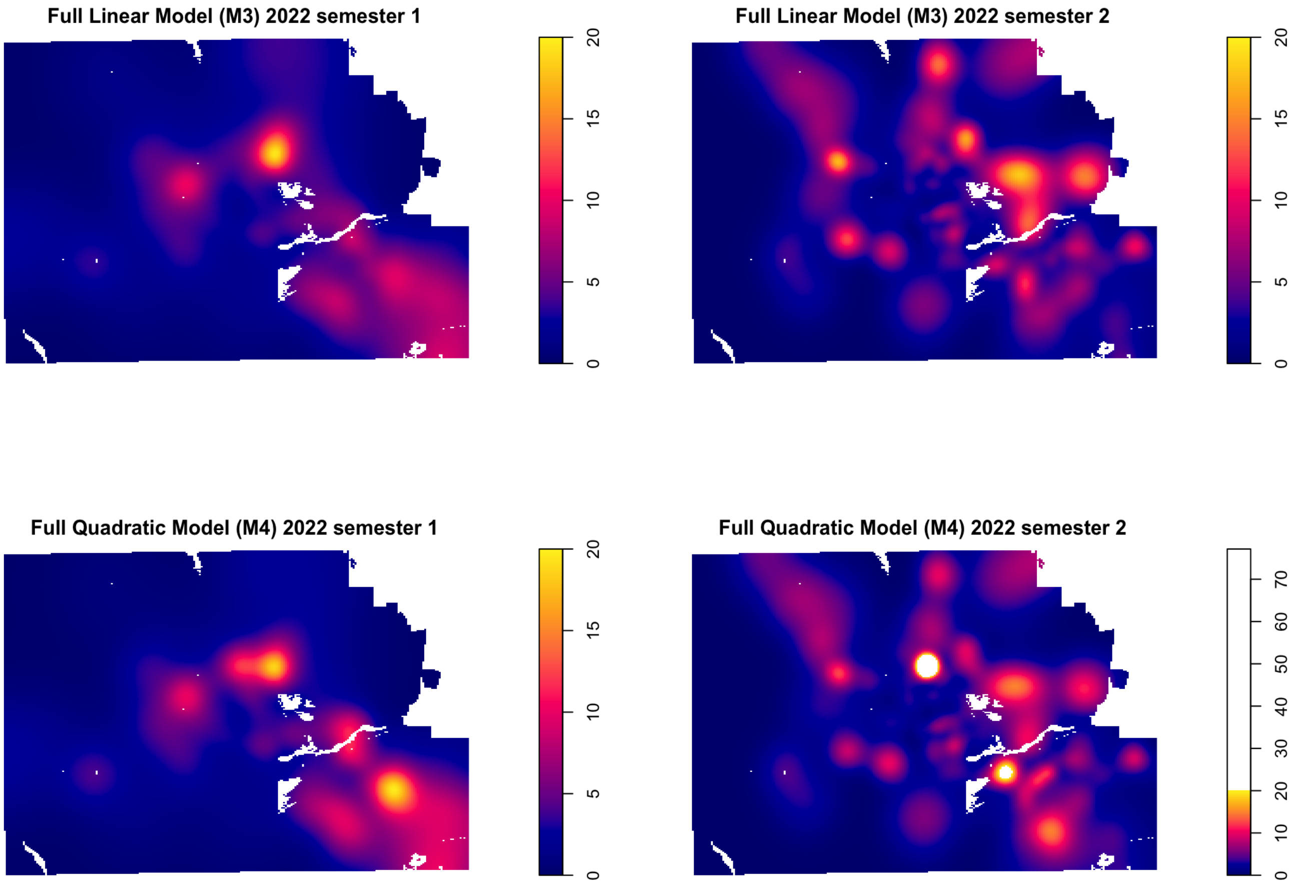
Full Linear Model (M3) 2022 semester 1
0
5
10
15
20
Full Linear Model (M3) 2022 semester 2
0
5
10
15
20
Full Quadratic Model (M4) 2022 semester 1
0
5
10
15
20
Full Quadratic Model (M4) 2022 semester 2
0
10
20
30
40
50
60
70

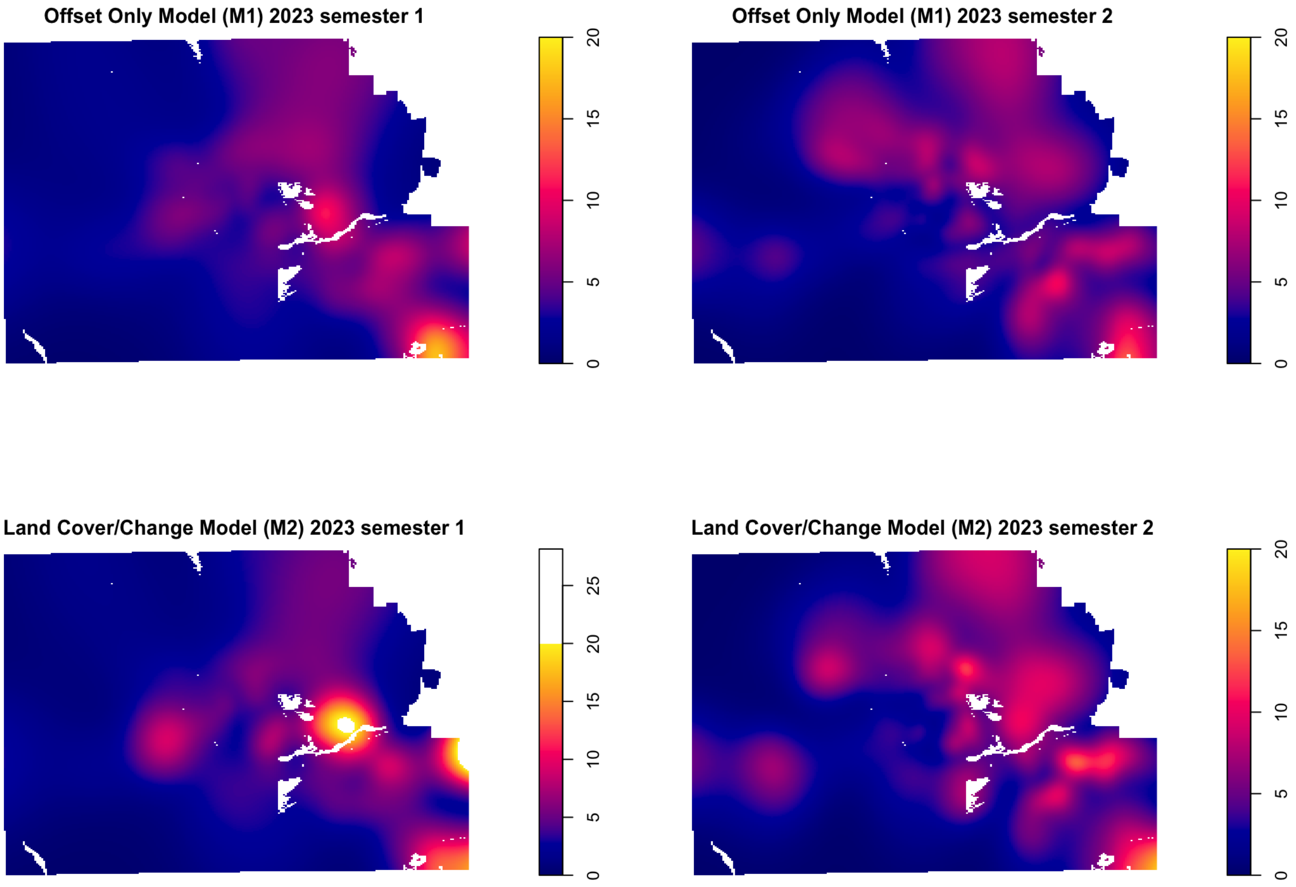
Offset Only Model (M1) 2023 semester 1
Offset Only Model (M1) 2023 semester 2
Land Cover/Change Model (M2) 2023 semester 1
Land Cover/Change Model (M2) 2023 semester 2
0
5
10
15
20
25

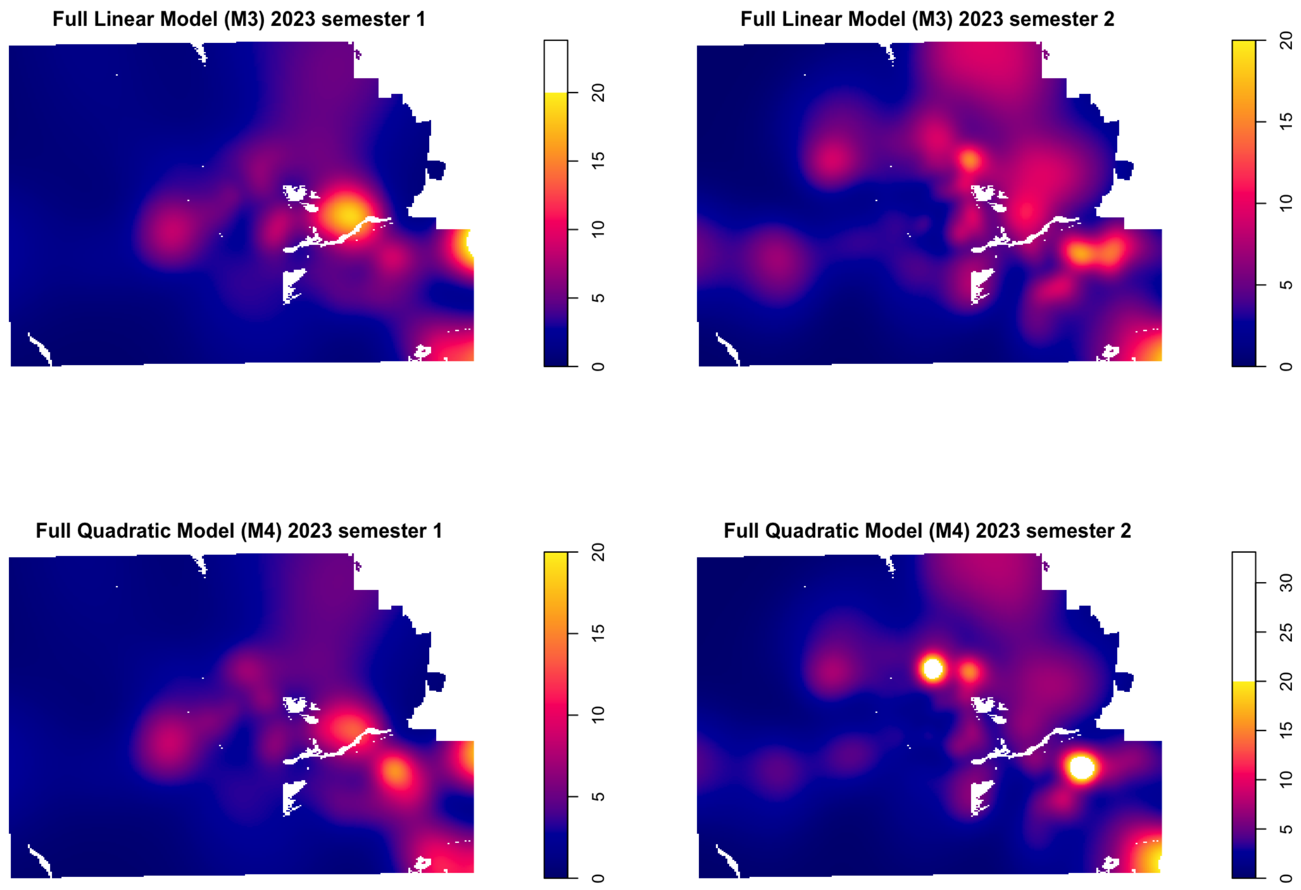
Full Linear Model (M3) 2023 semester 1
0
5
10
15
20
Full Linear Model (M3) 2023 semester 2
0
5
10
15
20
Full Quadratic Model (M4) 2023 semester 1
0
5
10
15
20
Full Quadratic Model (M4) 2023 semester 2
0
5
10
15
20
25
30

### Marginal effect curves from the full quadratic model (M4)

Below we show the marginal effect of NDVI (left column), NDVI difference (middle column) and HSI (right column) on intensity of VF cases based on the full quadratic model (M4) for all years/semesters. Effects are calculated by setting other continuous covariates to their 0.75 quantile. Land cover is fixed at the base category "Other". Shaded regions are 95% confidence bands. **Source:** AHCCCS, 2023. CHiR created patient record files from Arizona Medicaid (AHCCCS) data that were used for this analysis (including the date used for the models from which we have generated the plots below).

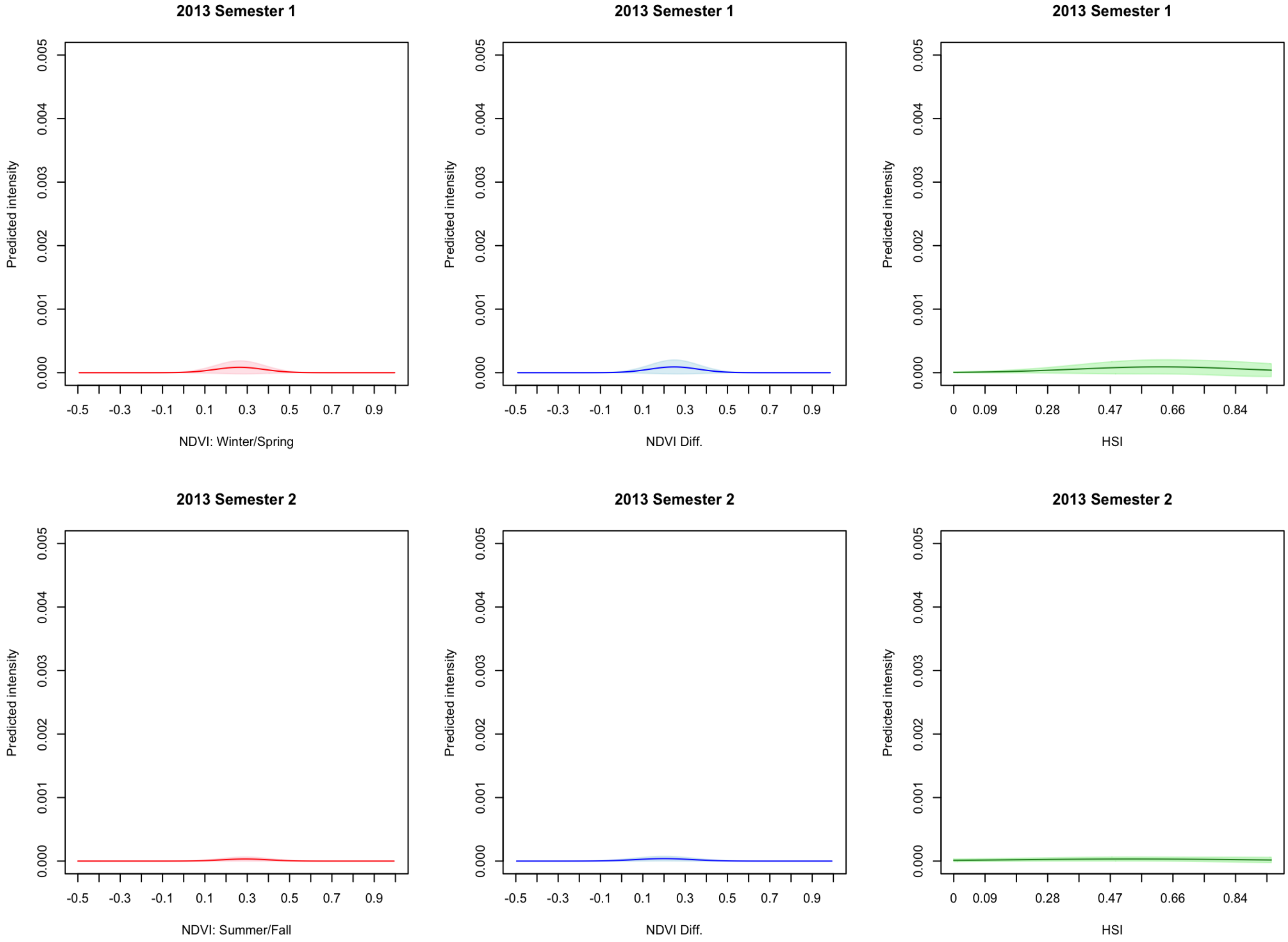
2013 Semester 1
Predicted intensity
0.000
0.001
0.002
0.003
0.004
0.005
-0.5
-0.3
-0.1
0.1
0.3
0.5
0.7
0.9
NDVI: Winter/Spring
2013 Semester 1
NDVI Diff.
2013 Semester 1
0
0.09
0.28
0.47
0.66
0.84
HSI
2013 Semester 2
NDVI: Summer/Fall
2013 Semester 2
NDVI Diff.
2013 Semester 2
HSI

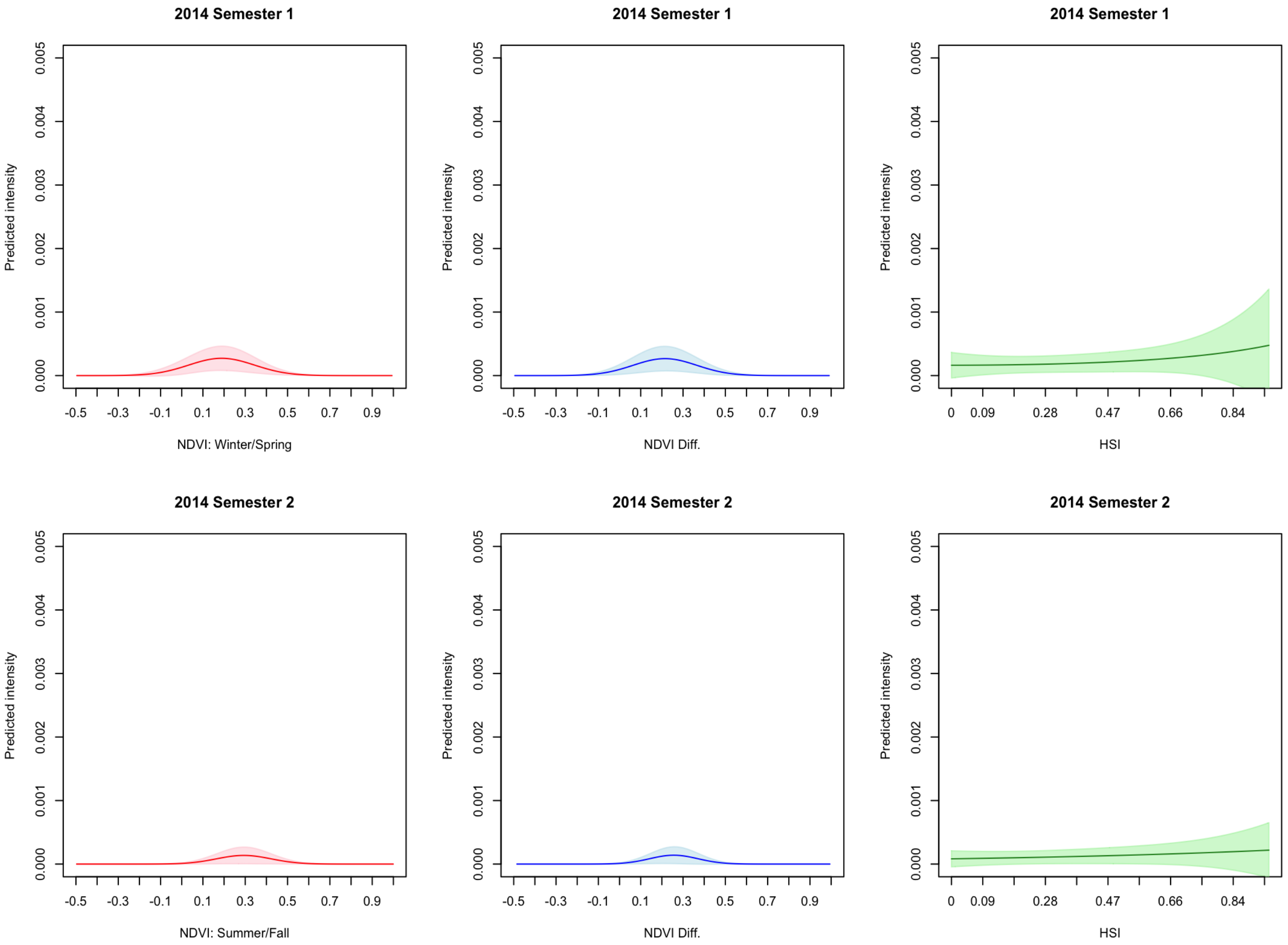
2014 Semester 1
Predicted intensity
0.000
0.001
0.002
0.003
0.004
0.005
-0.5
-0.3
-0.1
0.1
0.3
0.5
0.7
0.9
NDVI: Winter/Spring
2014 Semester 1
NDVI Diff.
2014 Semester 1
0
0.09
0.28
0.47
0.66
0.84
HSI
2014 Semester 2
NDVI: Summer/Fall
2014 Semester 2
NDVI Diff.
2014 Semester 2
HSI

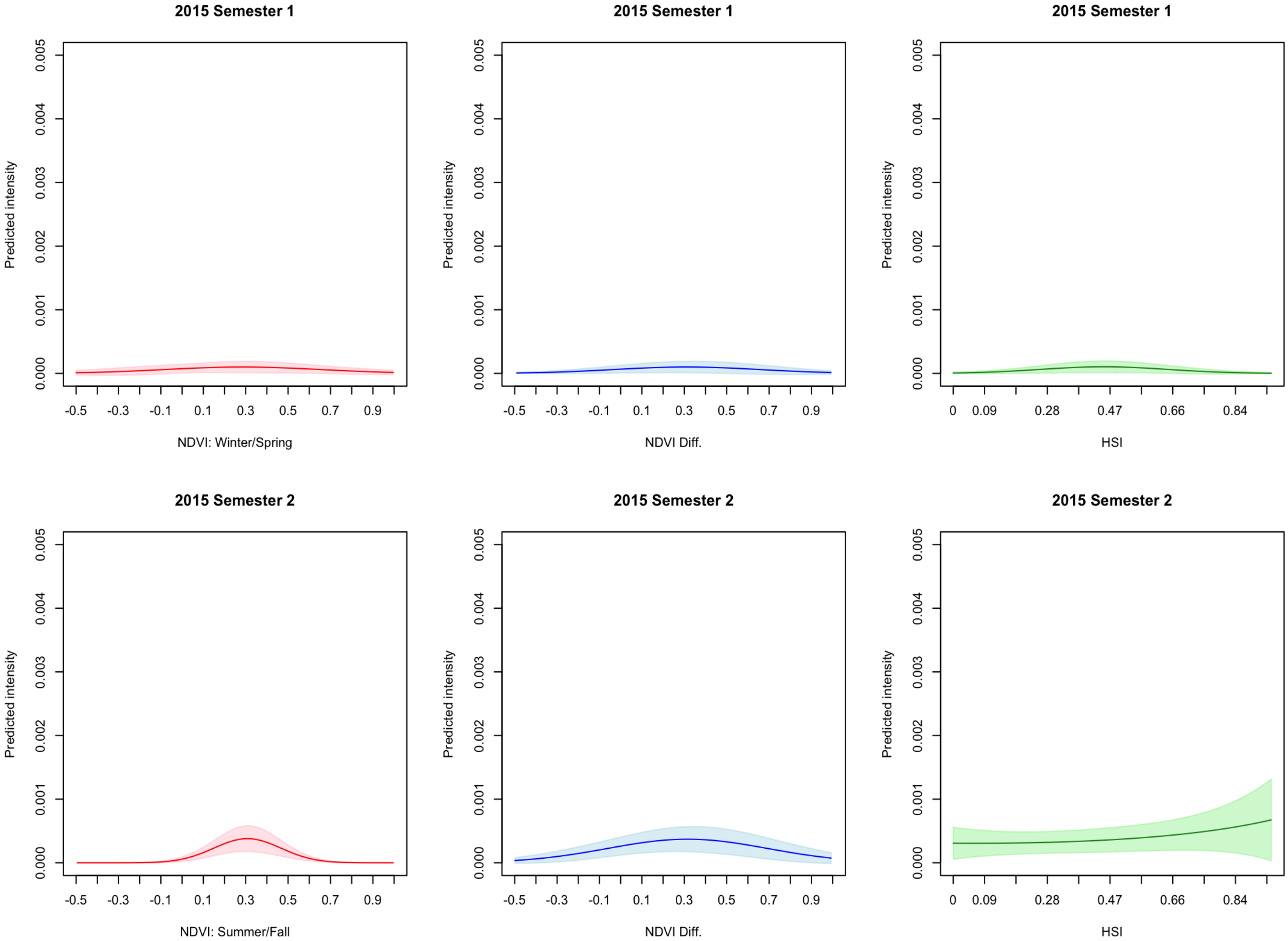
2015 Semester 1
Predicted intensity
0.000
0.001
0.002
0.003
0.004
0.005
-0.5
-0.3
-0.1
0.1
0.3
0.5
0.7
0.9
NDVI: Winter/Spring
2015 Semester 1
NDVI Diff.
2015 Semester 1
0
0.09
0.28
0.47
0.66
0.84
HSI
2015 Semester 2
NDVI: Summer/Fall
2015 Semester 2
NDVI Diff.
2015 Semester 2
HSI

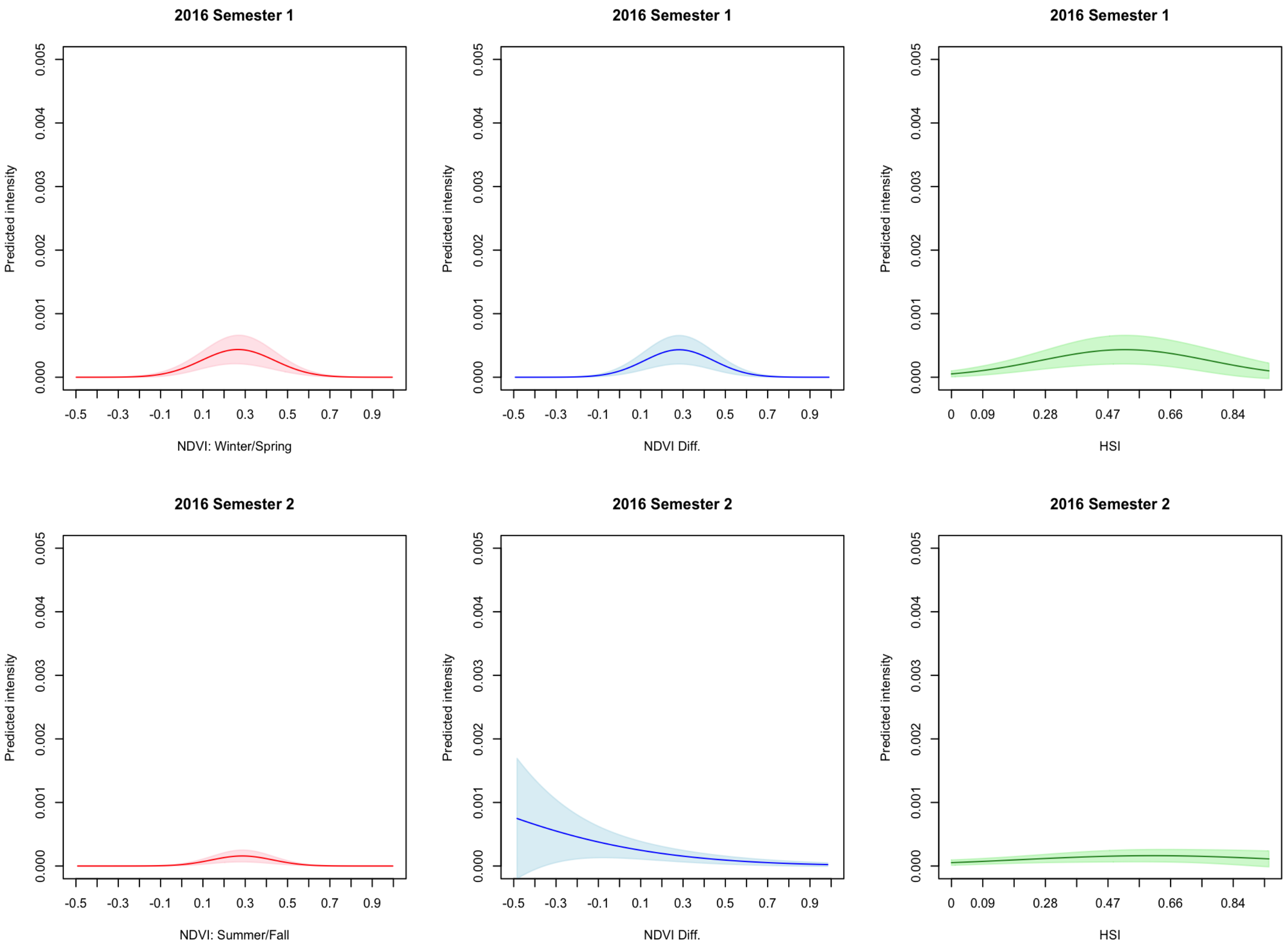
2016 Semester 1
Predicted intensity
0.000
0.001
0.002
0.003
0.004
0.005
-0.5
-0.3
-0.1
0.1
0.3
0.5
0.7
0.9
NDVI: Winter/Spring
2016 Semester 1
NDVI Diff.
2016 Semester 1
0
0.09
0.28
0.47
0.66
0.84
HSI
2016 Semester 2
NDVI: Summer/Fall
2016 Semester 2
NDVI Diff.
2016 Semester 2
HSI

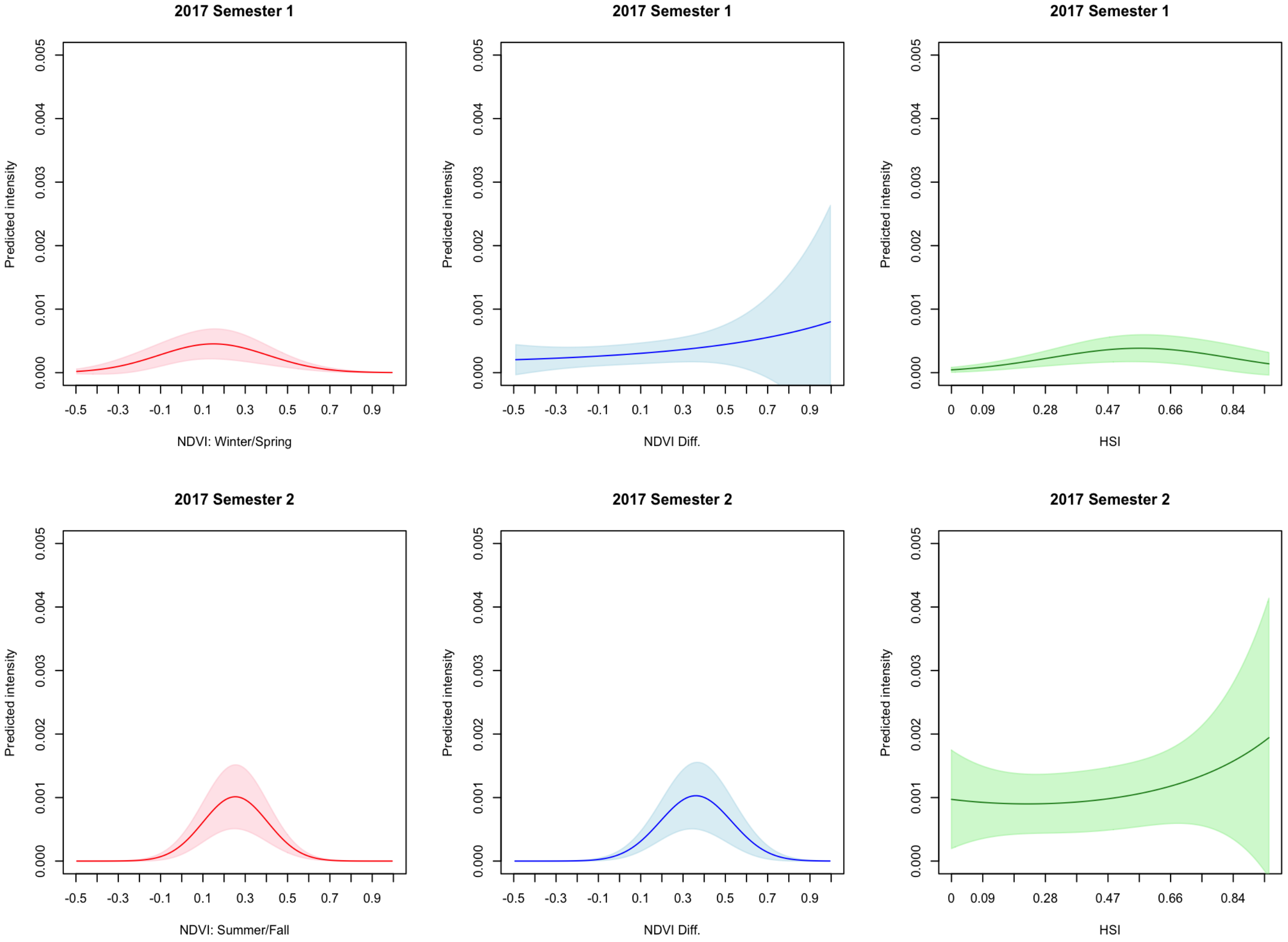
2017 Semester 1
Predicted intensity
0.000
0.001
0.002
0.003
0.004
0.005
-0.5
-0.3
-0.1
0.1
0.3
0.5
0.7
0.9
NDVI: Winter/Spring
NDVI Diff.
0
0.09
0.28
0.47
0.66
0.84
HSI
2017 Semester 2
NDVI: Summer/Fall

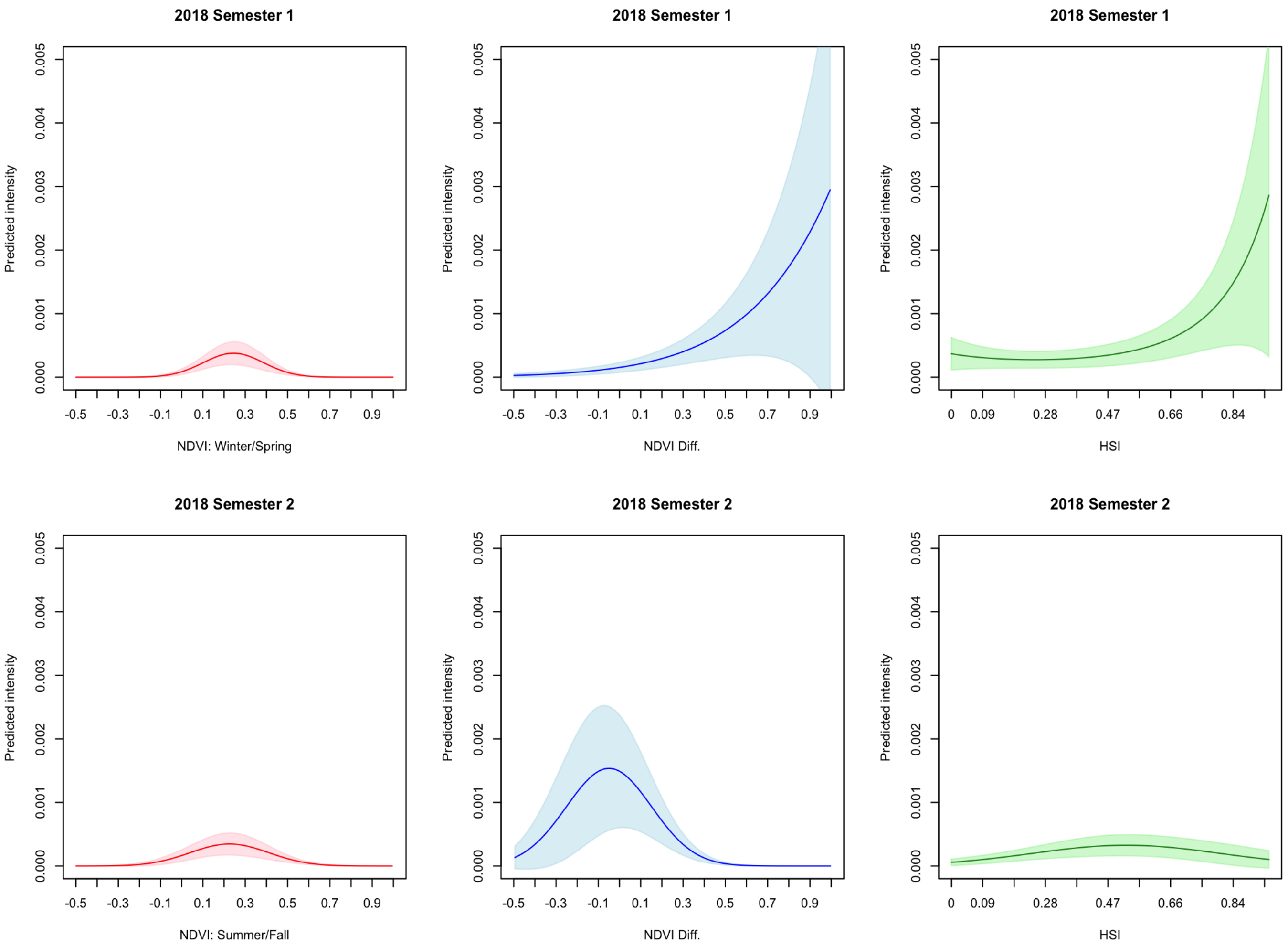
2018 Semester 1
Predicted intensity
0.000
0.001
0.002
0.003
0.004
0.005
-0.5
-0.3
-0.1
0.1
0.3
0.5
0.7
0.9
NDVI: Winter/Spring
NDVI Diff.
0
0.09
0.28
0.47
0.66
0.84
HSI
2018 Semester 2
NDVI: Summer/Fall

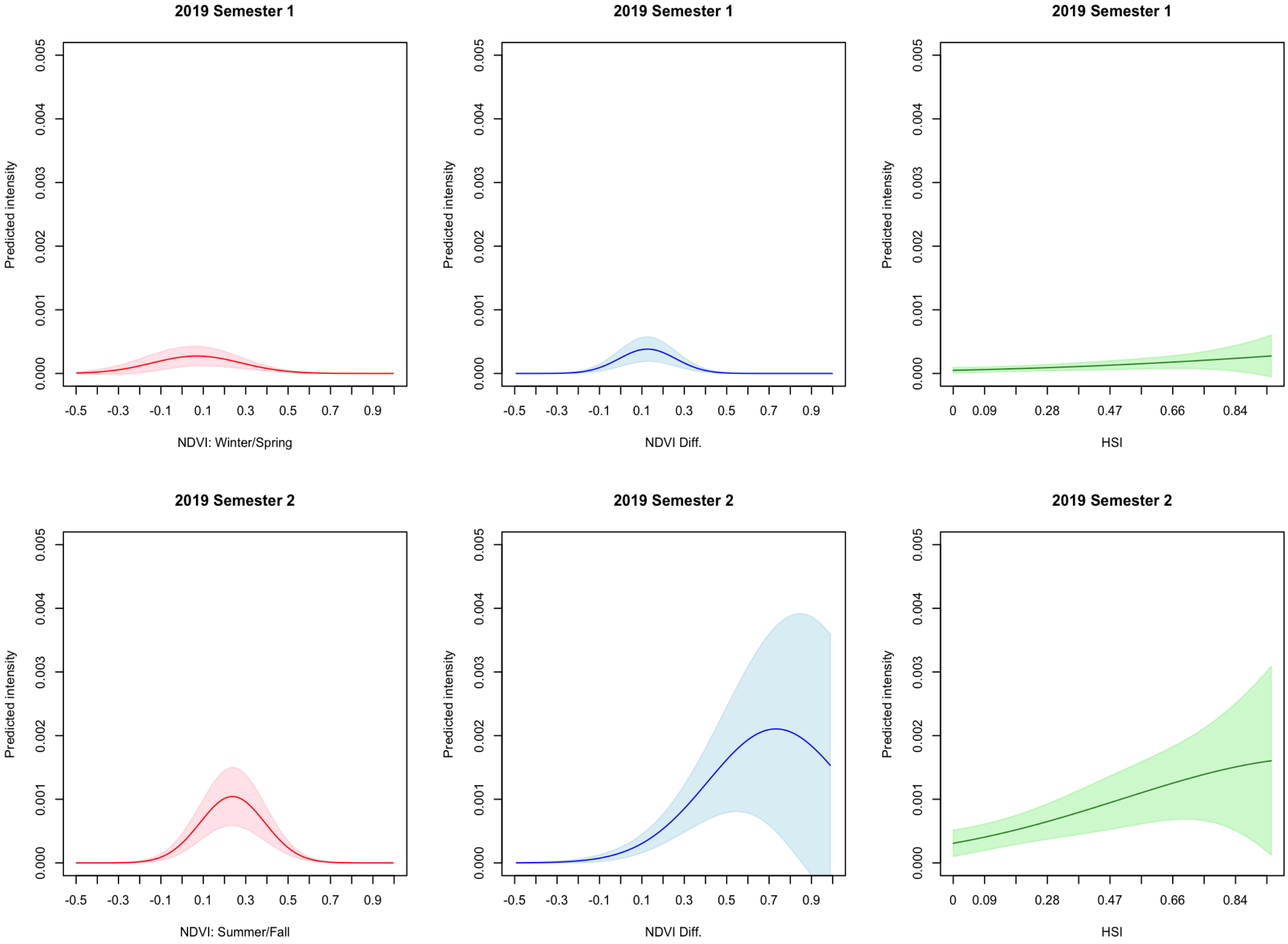

2019 Semester 1
Predicted intensity
0.000
0.001
0.002
0.003
0.004
0.005
-0.5
-0.3
-0.1
0.1
0.3
0.5
0.7
0.9
NDVI: Winter/Spring
2019 Semester 1
NDVI Diff.
2019 Semester 1
0
0.09
0.28
0.47
0.66
0.84
HSI
2019 Semester 2
NDVI: Summer/Fall
2019 Semester 2
NDVI Diff.
2019 Semester 2
HSI

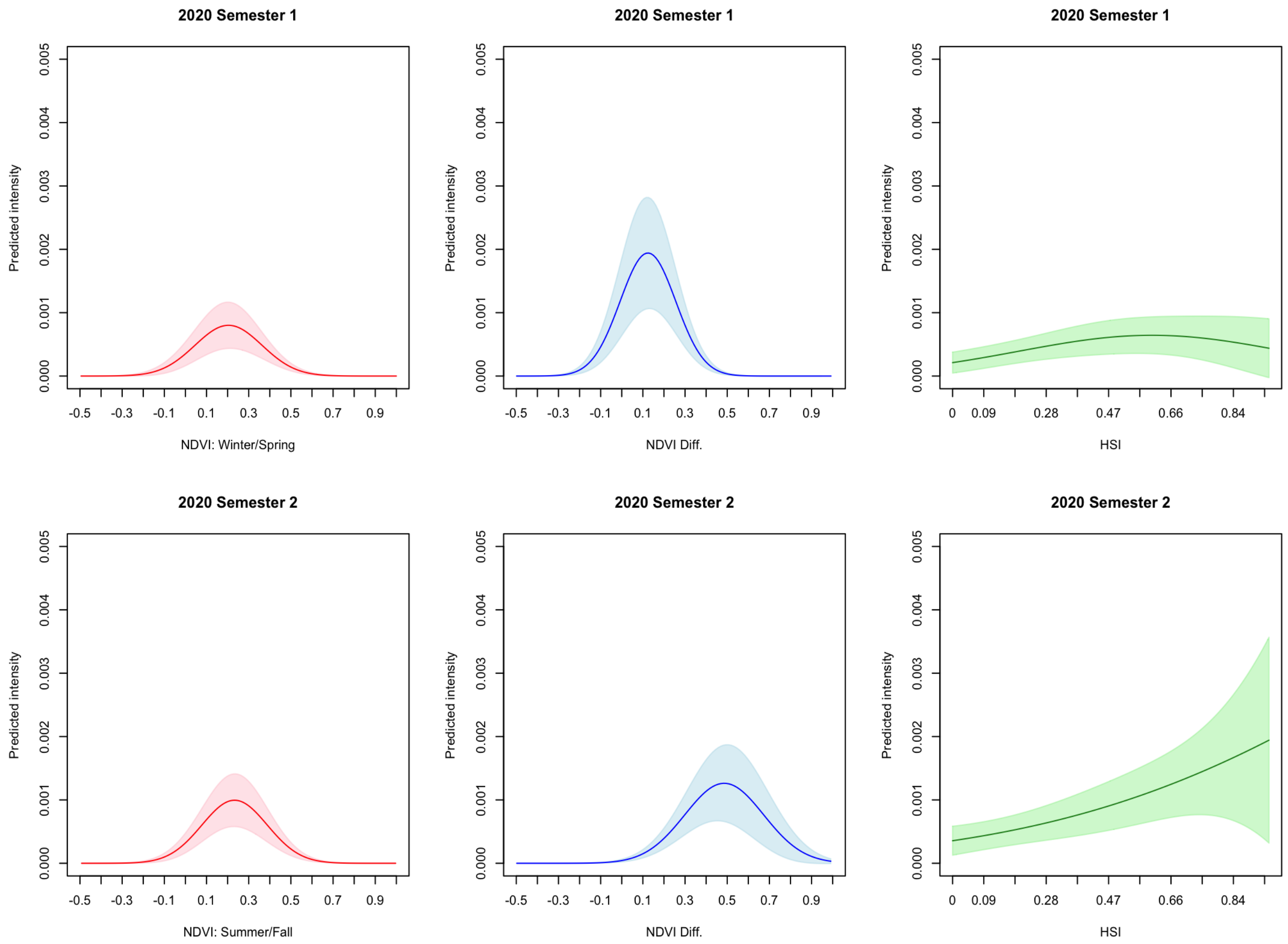

2020 Semester 1
Predicted intensity
0.000
0.001
0.002
0.003
0.004
0.005
-0.5
-0.3
-0.1
0.1
0.3
0.5
0.7
0.9
NDVI: Winter/Spring
2020 Semester 1
NDVI Diff.
2020 Semester 1
0
0.09
0.28
0.47
0.66
0.84
HSI
2020 Semester 2
NDVI: Summer/Fall
2020 Semester 2
NDVI Diff.
2020 Semester 2
HSI

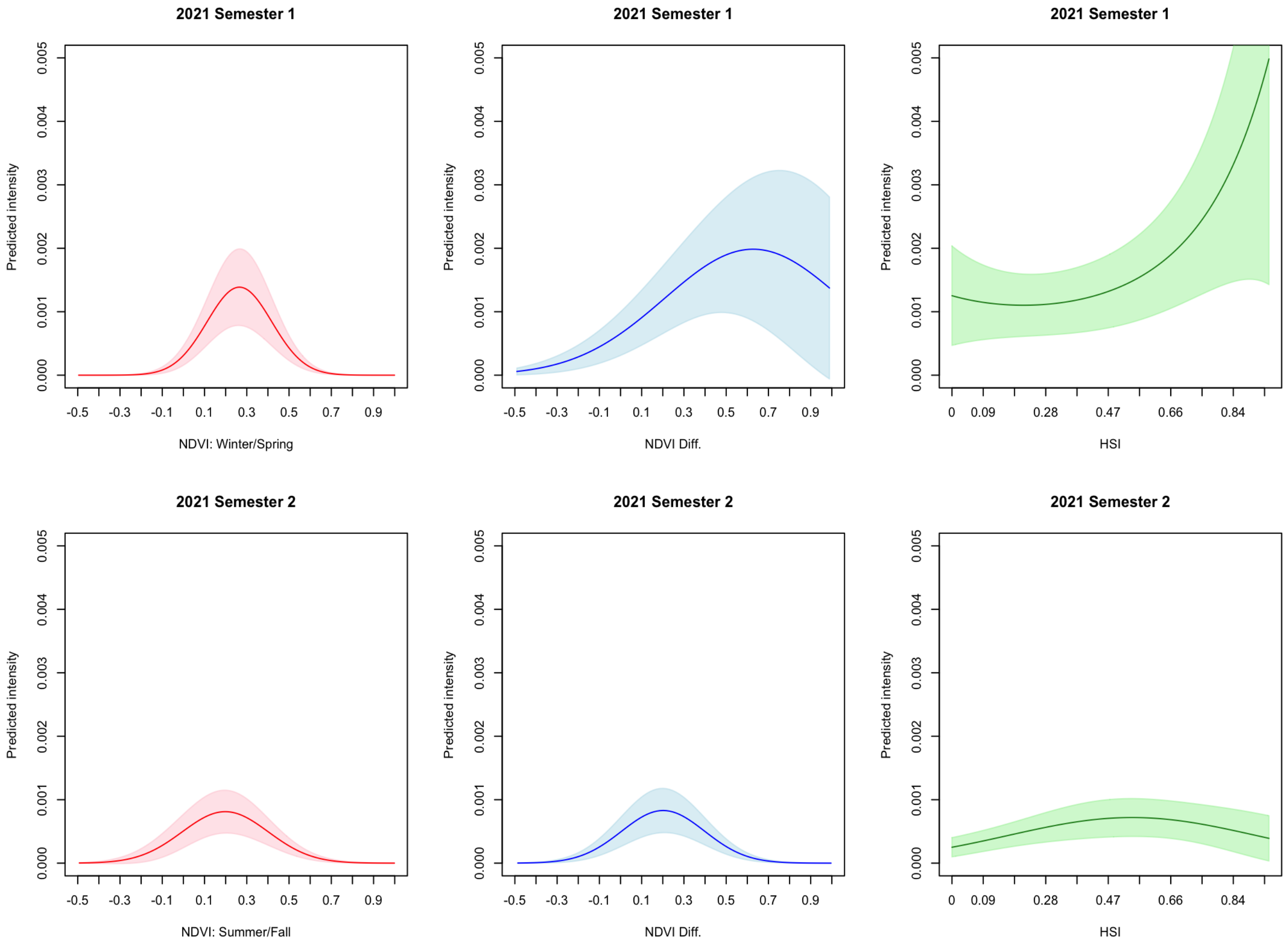
2021 Semester 1
Predicted intensity
0.000
0.001
0.002
0.003
0.004
0.005
-0.5
-0.3
-0.1
0.1
0.3
0.5
0.7
0.9
NDVI: Winter/Spring
2021 Semester 1
NDVI Diff.
2021 Semester 1
0
0.09
0.28
0.47
0.66
0.84
HSI
2021 Semester 2
NDVI: Summer/Fall
2021 Semester 2
NDVI Diff.
2021 Semester 2
HSI

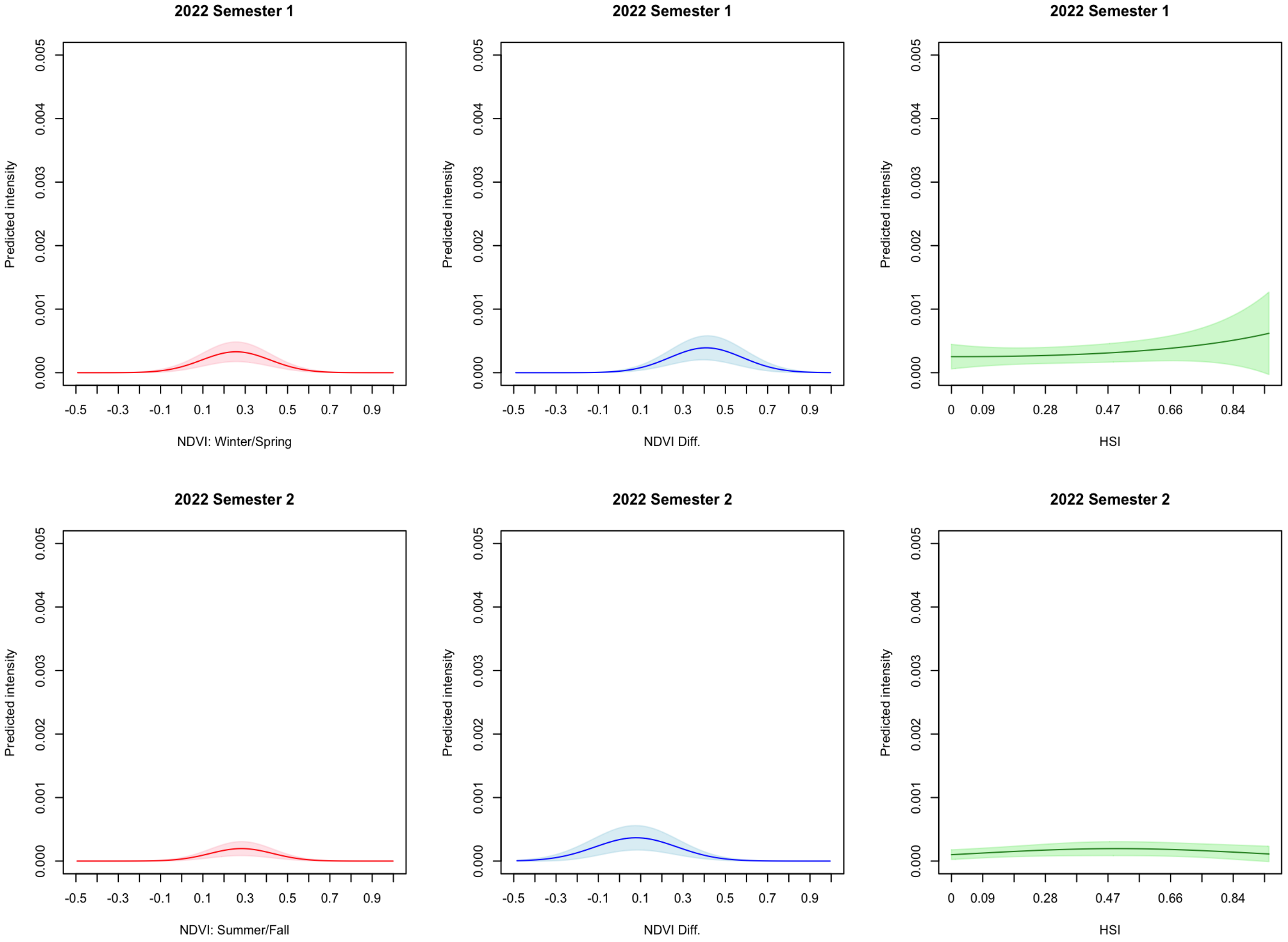
2022 Semester 1
Predicted intensity
0.000
0.001
0.002
0.003
0.004
0.005
-0.5
-0.3
-0.1
0.1
0.3
0.5
0.7
0.9
NDVI: Winter/Spring
2022 Semester 1
NDVI Diff.
2022 Semester 1
0
0.09
0.28
0.47
0.66
0.84
HSI
2022 Semester 2
NDVI: Summer/Fall
2022 Semester 2
NDVI Diff.
2022 Semester 2
HSI

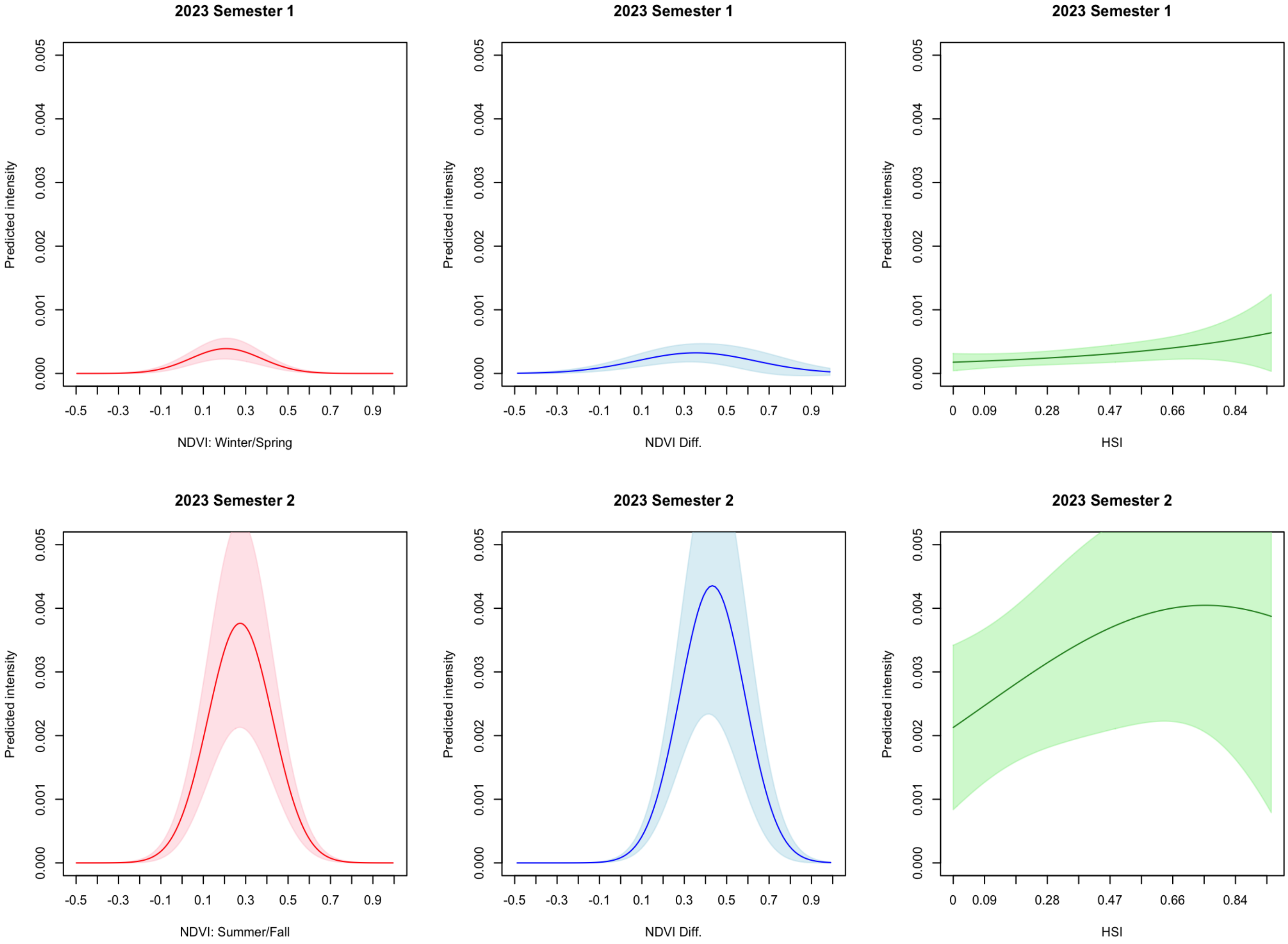
2023 Semester 1
Predicted intensity
0.000 0.001 0.002 0.003 0.004 0.005
-0.5 -0.3 -0.1 0.1 0.3 0.5 0.7 0.9
NDVI: Winter/Spring
2023 Semester 1
Predicted intensity
NDVI Diff.
2023 Semester 1
Predicted intensity
0 0.09 0.28 0.47 0.66 0.84
HSI
2023 Semester 2
Predicted intensity
NDVI: Summer/Fall
2023 Semester 2
Predicted intensity
NDVI Diff.
2023 Semester 2
Predicted intensity
HSI
